# Changes in Cardiovascular Disease Risk, Lung Function, and Other Clinical Health Outcomes When People Who Smoke Use E-cigarettes to Reduce Cigarette Smoking: An Exploratory Analysis from a Randomized Placebo-Controlled Trial

**DOI:** 10.1101/2024.12.14.24319048

**Authors:** Sitasnu Dahal, Jessica Yingst, Xi Wang, Caroline O. Cobb, Matthew Carrillo, Shari Hrabovsky, Rebecca Bascom, Alexa A Lopez, Le Kang, Sarah Maloney, Matthew Halquist, Jonathan Foulds, Susan Veldheer

## Abstract

**Introduction:** The health effects of dual use of electronic cigarettes (e-cigarettes) and combustible cigarettes are unclear. We report on differences in cardiovascular disease (CVD) risk factors, lung function, and clinical laboratory markers among people who smoke used e-cigarettes to reduce their cigarette smoking in a randomized placebo-controlled trial.

**Methods:** Participants (n=520) who regularly smoked cigarettes were randomized to 1 of 4 conditions (e-cigarette device paired with liquid containing 0, 8, or 36 mg/mL of nicotine or a cigarette-substitute [CS]) and encouraged to reduce their smoking over 6 months. Group differences were assessed between the e-cigarettes and CS conditions at baseline and 6-month using one-way ANOVA and linear mixed-effects model. Multi-testing adjustment was not applied as the analysis was exploratory in nature. Primary outcomes were: CVD risk (i.e., blood lipids, C-reactive protein, blood pressure, heart rate, waist-to-hip ratio, body mass index, and INTERHEART risk score), lung function (i.e., spirometry indices and clinical COPD questionnaire), and other clinical laboratory markers (i.e., complete blood count and complete metabolic panel).

**Results:** At 6-month, use of nicotine e-cigarettes caused no significant differences from control groups for most measures. However, participants randomized to 36 mg/mL e-cigarettes had significantly higher levels of high-density lipoprotein (HDL) (*p*=0.003 in unadjusted analysis, *p*=0.002 in adjusted analysis), and lower levels of low-density lipoprotein (LDL) (*p*=0.044 in adjusted analysis) and cholesterol/HDL ratio (*p*=0.034 in unadjusted analysis, *p*=0.026 in adjusted analysis) as compared to CS. Also, those in the 36 mg/mL e-cigarette condition had higher levels of HDL as compared to those in 0 mg/mL condition (*p*=0.016 in unadjusted analysis, *p*=0.019 in adjusted analysis).

**Conclusions:** Those randomized to the highest nicotine e-cigarettes had small improvement in some measures of blood lipids (e.g., increased HDL, and reduced LDL and cholesterol/HDL ratio) as compared to a non-aerosol CS among individuals attempting to reduce their cigarette smoking. Future studies of e-cigarettes for smoking cessation would benefit from including these measures to further explore the results found in this study.

**Trial Registration:** ClinicalTrials.gov, NCT02342795.

**Strengths and limitations of this study:**

- The use of a randomized double-blind placebo-controlled trial design with two nicotine concentrations and a relatively long follow-up period (6 months).
- Use of an additional randomized control group who did not use an electronic cigarette device, but were given a “cigarette substitute” product with no aerosol but similar behavioral requirements.
- Participants had to be daily cigarette smokers with no plans to quit smoking, but an interest in reducing, and were recruited from two U.S. sites. The results may only be generalizable to similar populations.
- These were exploratory analyses of a comprehensive group of commonly used clinical markers, rather than hypothesis-driven primary outcomes. These results therefore provide a preliminary assessment of effects that may inform future studies.

## BACKGROUND

Electronic nicotine delivery systems, popularly known as electronic cigarettes (e-cigarettes), are a diverse class of nicotine delivery products that have grown in popularity. The prevalence of current e-cigarette use among adults in the United States increased from 3.7% in 2020 to 6.6% in 2023.^1,2^ E-cigarettes vary greatly in design elements and power settings, and work by heating a liquid containing nicotine, propylene glycol, glycerin, and flavoring agents to produce an aerosol that is inhaled via a mouthpiece.^3^

The public health impact of e-cigarette use is a matter of substantial debate.^4^ On the one hand, e-cigarette aerosol contains and delivers fewer and lower concentrations of toxic chemicals than tobacco cigarettes (TCs).^5^ Further, findings from a Cochrane review that included 56 randomized controlled trials (RCTs) found that nicotine-containing e-cigarettes helped more people to achieve abstinence from TCs for six months and longer as compared to a nicotine-free e-cigarettes or nicotine replacement therapy.^6^ This evidence suggests that e-cigarettes may be a potential harm reduction tool for people who smoke TC.

On the other hand, e-cigarettes are likely not completely harmless and there are many concerns that e-cigarettes may serve as an initiation product for TCs among adolescents and young adults.^7^ Further, it has been found that e-cigarettes contain some toxic chemicals, including carcinogens, heavy metals, and ultrafine particles that could cause damage when inhaled into the lungs.^5,8^ Exposure to e-cigarette aerosol could produce a range of inflammatory reactions in different organ systems, including the pulmonary, cardiovascular, gastrointestinal, renal, neurological, ophthalmic, and immune system.^9,10^ Although e-cigarettes are not widely approved as smoking cessation devices, one of the most comprehensive reviews for the UK government concluded that, “Our findings that vaping carries only a small fraction of the health risks of smoking, suggest that smokers should be encouraged to use vaping products for stopping smoking, or as an alternative nicotine delivery device to reduce the health harms of smoking.”^11^ However, given the novelty of the e-cigarettes, the long-term health impacts remain unknown.

Part of the challenge in addressing concerns about health effects of e-cigarettes use is that the available evidence has provided conflicting results using both human and animal models.^12–15^ In addition, conclusive evidence on harm or benefit of e-cigarette use in human using standard clinical markers is either mixed or absent. For instance, recent investigations^16–18^ have found no significant changes in spirometry measures after short to mid-term use of e-cigarettes (from 8 days to 12 months) among individuals who use TC completely or partially switch. In contrast, Cibella et al, (2016) showed progressive improvements in FEF 25–75% (a sensitive marker of obstructive peripheral airflow) among people who smoke TCs switched completely to e-cigarettes over one year.^19^ Regarding cardiovascular health, several studies^18,20–22^ have reported a positive impact, including heart rate and blood pressure when switching from TC to e-cigarette, whereas others did not observe improvements in these outcomes.^13,17^ Similarly, there are few studies that have explored the effects of e-cigarette use on complete blood count (CBC), complete metabolic panel (CMP), and blood lipid levels, and these studies also have disparate findings.^11,23–25^ Furthermore, most findings are derived mainly from non-randomized clinical studies with methodological dissimilarities to reach meaningful and generalizable conclusions. In addition, most of the research on these health effects studied use of e-cigarettes with undefined nicotine delivery properties and without randomized comparison groups.

Thus, there are critical gaps in the existing literature regarding pulmonary, cardiovascular, and other health effects of e-cigarette use among those who are currently smoking. The present report is an exploratory study from a four-arm randomized placebo-controlled trial designed to examine the effects of e-cigarette use on tobacco-related toxicant exposure among individuals who were attempting to reduce their TC consumption using an e-cigarette or a cigarette substitute (CS) that does not emit aerosol or nicotine.^26^ These exploratory analyses were planned to provide a preliminary assessment of the potential effects of e-cigarette use on cardiovascular disease (CVD) risk factors, pulmonary function, and other standard clinical laboratory markers when used by individuals trying to reduce their cigarette consumption. Given that many smokers who try e-cigarettes continue to smoke (dual use), this study provides an opportunity to explore potential harmful effects on health markers of dual use.

## METHODS

### Main trial design and participants

This study was a two-site (Virginia Commonwealth University, Richmond, VA and Penn State College of Medicine, Hershey, PA), four-arm, parallel-group, randomized placebo-controlled trial, with the three e-cigarette conditions administered in a double-blind manner. Details of the trial have been published previously.^26–29^ Briefly, 520 adults aged 21-65 years who smoked at least 10 cigarettes per day (CPD), had an expired-air carbon monoxide (CO) reading of > 9 parts per million (ppm) at baseline, and were interested in reducing their TC consumption were enrolled in the 9-month trial. After an assessment visit (V1), participants returned approximately 1 week later and were randomized, at no cost, to one of the four study conditions: cigarette substitute (CS) or a 0, 8, or 36 mg/mL liquid nicotine concentration e-cigarettes. Participants selected the flavor of their e-cigarette liquid (tobacco or menthol) after briefly testing each flavor in the clinic; but they could not change it thereafter. During the randomization phase of the study, participants were encouraged to reduce their TC smoking by 50% for 2 weeks, then by 75% for 2 weeks, and then to maintain 75% reduction and/or continue to try to reduce for the remainder of the trial to week 24. At each visit after randomization, participants received brief counseling, including a discussion of their CPD reduction goals and they were encouraged to use their assigned study product as a substitute for TC. Participants were compensated for their in-person and remote visits. This trial is registered with ClinicalTrials.gov, NCT02342795. The consort diagram showing recruitment and retention can be found in online supplemental figure 1.

### Study products

The study products used included e-cigarettes and a CS. The e-cigarettes consisted of a 3.3 – 4.1 volt, 1100 mAh rechargeable battery and a 1.5 Ohm, dual-coil, 510-style cartomizer (SmokTech, Shenzhen, China). It had a button that activated the battery to heat the coil and produce an aerosol that is inhaled through the mouthpiece. The CS (QuitSmart, Inc., NC) was a plastic tube that resembled a cigarette. It had an adjustable draw to resemble the feel of a cigarette, but it did not have any electronic parts, and it did not contain nicotine or emit any aerosol. The CS serves as a true control group to ascertain the effects due to e-cigarette aerosol (nicotine containing or not). Specific details of these products and the procedures for dispensing blinded, randomized nicotine concentrations to participants have been previously published.^26–29^ Primary outcome analyses revealed that those randomized to the 36 mg/mL e-cigarettes had a significantly lower cigarette consumption, NNAL concentration, and exhaled CO at 6-month than those randomized to the CS.^30^

### Measures

#### Baseline socio-demographics and tobacco use characteristics

Demographic information included age, sex, race, education, and income. Tobacco use characteristics included number of CPD and participant’s age of smoking initiation.

#### Measured pulmonary function and self-reported pulmonary symptoms and functional state

Spirometry measurements were performed without a bronchodilator using the CareFusion SpiroUSB spirometer (Basingstoke, UK) by trained research staff at baseline, 1, 3, and 6 months. The primary focus of this report is on results from the baseline and 6-month follow-up timepoints. Participants completed 3 acceptable spirometry maneuvers at each timepoint, with assessment of repeatability. Indices measured include forced expiratory volume in 1 second in liters (FEV1), FEV1% (% predicted), forced vital capacity in liters (FVC), FVC% (% predicted), FEV1/FVC ratio, FEV1/FVC% (% predicted), peak expiratory flow in liters per second (PEF), PEF% (% predicted), forced expiratory flow in liters per second (FEF) at 25% of the FVC, 75% of the FVC, and between 25-75% of the FVC, FEF (25-75%) (% predicted), and forced expiratory time (FET) in seconds. The best FEV1 and best FVC were retained; other indices were determined from the maneuver with the largest sum of FVC and FEV1.

Self-reported pulmonary health status was measured at baseline and 6-month using the 10-item Clinical COPD Questionnaire (CCQ) which assesses three domains: symptoms (dyspnea, cough, phlegm), functional state (limitations in activities of daily life due to breathing) and mental state (feelings of depression or concerns about breathing).^30^ Scores for each of the three domains were calculated using standard analytical guidelines. The overall scores range between 0 (best possible clinical control) to 6 (worst possible clinical control). The CCQ is a validated patient-reported outcome measure of symptoms and functional state, applicable not only to individuals diagnosed with COPD but also to those “at risk” of developing the condition (e.g., current smoking), thereby broadening its applicability in both clinical and research settings.^30^

#### Blood measurements

Blood samples were collected at baseline and 6-month by trained research staff and analyzed by the clinical lab at the Penn State Hershey Medical Center and Virginia Commonwealth University Medical Center Clinical Laboratory. Whole blood was used to assess white blood cells (WBC), red blood cells (RBC), platelets, hemoglobin, hematocrit, mean corpuscular volume (MCV), mean corpuscular hemoglobin (MCH), mean corpuscular hemoglobin concentration (MCHC), red cell distribution width (RDW), and mean platelet volume (MPV). The whole blood was centrifuged to obtain plasma for analyzing C-reactive protein (CRP), CMP (sodium, potassium, chloride, carbon dioxide, anion gap, glucose, blood urea nitrogen [BUN], aspartate aminotransferase [AST], alkaline transferase [ALT], alkaline phosphatase [ALP], bilirubin, protein, albumin, calcium, and creatinine). Lipoprotein measures included total cholesterol, high-density lipoprotein (HDL), non-HDL cholesterol, cholesterol/HDL ratio, low-density lipoprotein (LDL), and triglycerides. All of these laboratory outcomes are non-fasting measurements.

#### Other physiological measures

Blood pressure (BP) and heart rate were measured using an electronic sphygmomanometer after participants were seated quietly for ≥ 5 minutes. Weight was collected on digital scales that were calibrated according to the manufacturer’s specifications. Height was measured using a standard stadiometer. Body mass index (BMI) was calculated as the weight in kg divided by the square of height in meter. Participants’ hip and waist circumferences were measured by trained research staff and the waist-to-hip ratio was calculated. Cardiovascular disease (CVD) risk was assessed using the INTERHEART risk score, a validated tool for quantifying risk-factor burden without laboratory testing.^31^ The non-laboratory risk calculator includes self-reported items relating to smoking, diabetes, blood pressure, psychosocial factors, diet, physical activity, family history of CVD, biological sex, age, and measured waist-to-hip ratio. Scores range from 0 to 48; higher scores indicate a greater risk-factor burden.

### Statistical analysis

We first examined the overall characteristics of the study population by comparing baseline demographics, tobacco use, and psychosocial/health characteristics among the four randomized conditions. We calculated frequencies (percentages) and *p*-values using chi-square tests for categorical variables, as well as means, standard deviations (SD), and *p*-values using ANOVA test for continuous variables. We used one-way ANOVA tests to assess between group differences (i.e., CS, 0 mg/mL, 8 mg/mL and 36 mg/mL nicotine e-cigarettes) for clinical outcomes at baseline.

To compare the changes in clinical outcome measures across the randomized conditions at each time point (i.e., baseline, 1-month, 3-month and 6-month follow-up) and within conditions relative to week 0, we conducted linear mixed-effects models for both unadjusted and adjusted analyses. The model included main effect terms of categorical time (e.g., baseline, 1-month follow-up, etc.), and condition (e.g., CS, 0, 8, and 36 mg/mL nicotine e-cigarettes), and their interaction as fixed effects, assuming a different mean for each treatment at each time point. A random-effect term with variance–covariance matrix (in the unstructured, autoregressive structure of order 1, or the compound symmetry structure) determined by the fit statistics (e.g., Akaike information criterion, AIC) was included to capture the within-subject correlation across repeated measures analyses. Multi-testing adjustment was not applied in the pairwise comparisons as this analysis was exploratory.

The primary analyses used intention-to-treat (ITT) methodology, utilizing all available data at each visit without imputing missing values in the clinical outcome measures. In addition, we conducted three sensitivity analyses to investigate the robustness of the statistical inference to the untestable missing mechanism assumption for both unadjusted and adjusted analyses: (1) the per-protocol analyses included only completers who have the respective clinical outcome measure at baseline and the three follow-up visits; (2) the last-observation-carried-forward analyses filled missing values with the last observed clinical outcome measure of the participant; (3) the multiple imputation procedure invoked a parametric model for missing values imputation and generated multiple datasets to adjust for the variability in the imputation process; specifically, we generated 50 imputed datasets based on the fully conditional regression model.

The baseline covariates which included demographic characteristics, cigarettes per day, exhaled CO level, psychosocial/health characteristics etc. for adjusted analyses were selected using the stepwise model selection procedure via AIC. We used a fully conditional regression model (logistic regression for categorical variables and linear regression for continuous variables) as the imputation model to impute missing values in baseline participants’ characteristics. It assumed a separate conditional distribution for each imputed variable as implemented by the SAS procedure via the FCS statement and demonstrated comparable performance as another popular multiple imputation method with a multivariate normal distribution assumption.^32^ Detailed description of covariates included in fully conditional regression model and in the adjusted models are presented in Supplemental Table 1 and Supplemental Table 2 respectively.

## RESULTS

### Baseline socio-demographics and tobacco use characteristics

An overview of baseline study participant demographics and tobacco use characteristics across the four randomized groups is presented in Table 1. The sample was 58.8% female, 67.3% White, and had a mean age of 46.2 ± 11.6 years. Nearly 60% of the participants had some college education or more and 59% had an income of less than $39,999. Participants smoked an average of 18.6 ± 7.7 CPD and had a mean age of 18.1 ± 5.1 years when they initiated smoking. There were no significant differences in participant demographics or tobacco use characteristics across randomized conditions. The number of participants in each outcome measurements across randomized arms at 6-month has been presented in Supplemental Table 3.

**Table 1.**
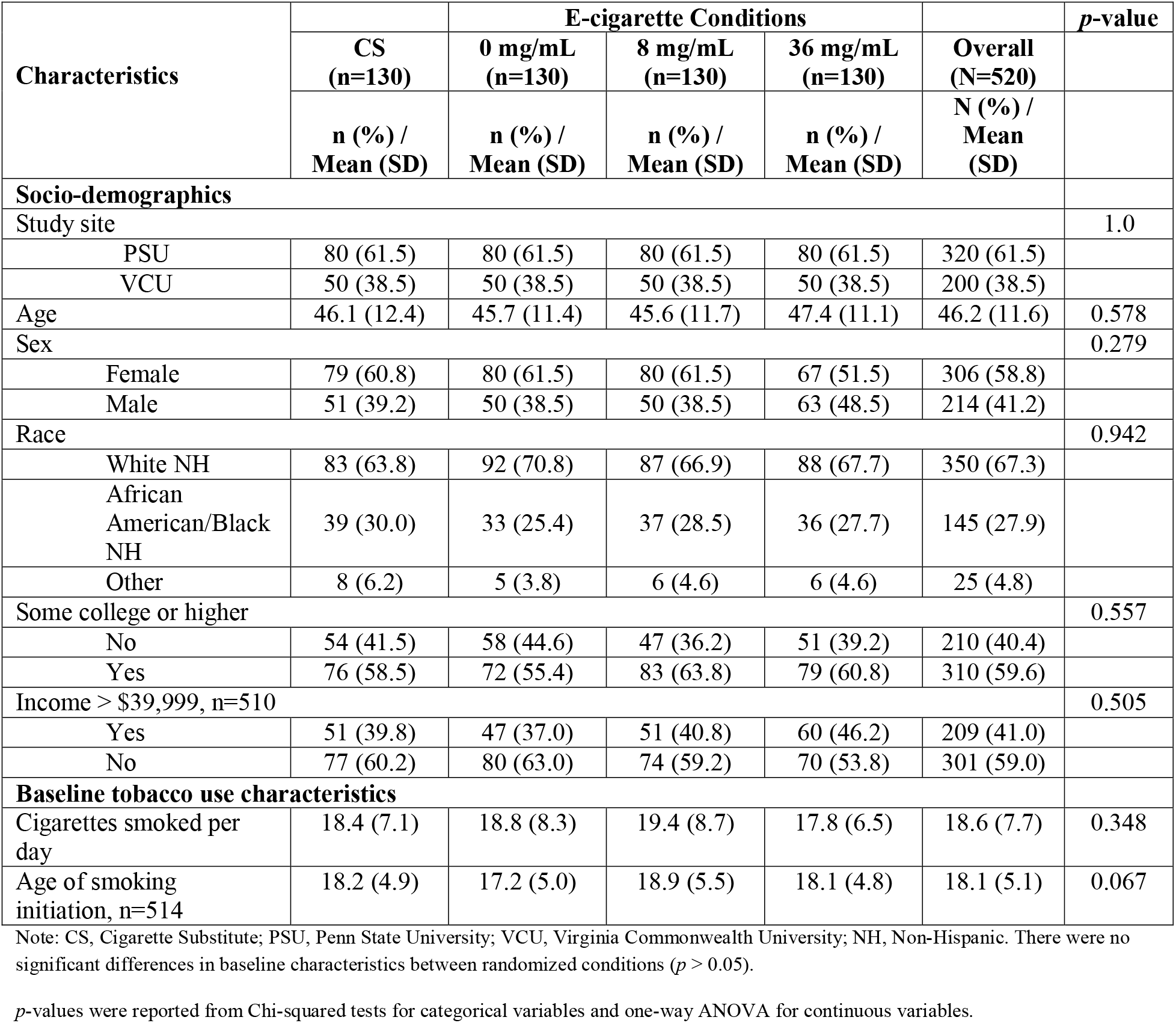
Baseline Characteristics of Study Participants by Overall and Conditions.

### Measured pulmonary function and self-reported pulmonary symptoms and functional state

At baseline, there were no significant differences between the randomized conditions for any of the spirometry indices (Table 2). At 6-month, there were also no differences across randomized conditions in FEV1 (Table 3; Figure 1A) and other PFT measures (Table 3) with the exception of FET which was significantly higher among those using 36 mg/mL e-cigarettes as compared to CS group in both unadjusted (*p* = 0.033; Table 3) and adjusted analysis (*p* = 0.02; Supplemental Figure 14).

**Figure 1.**
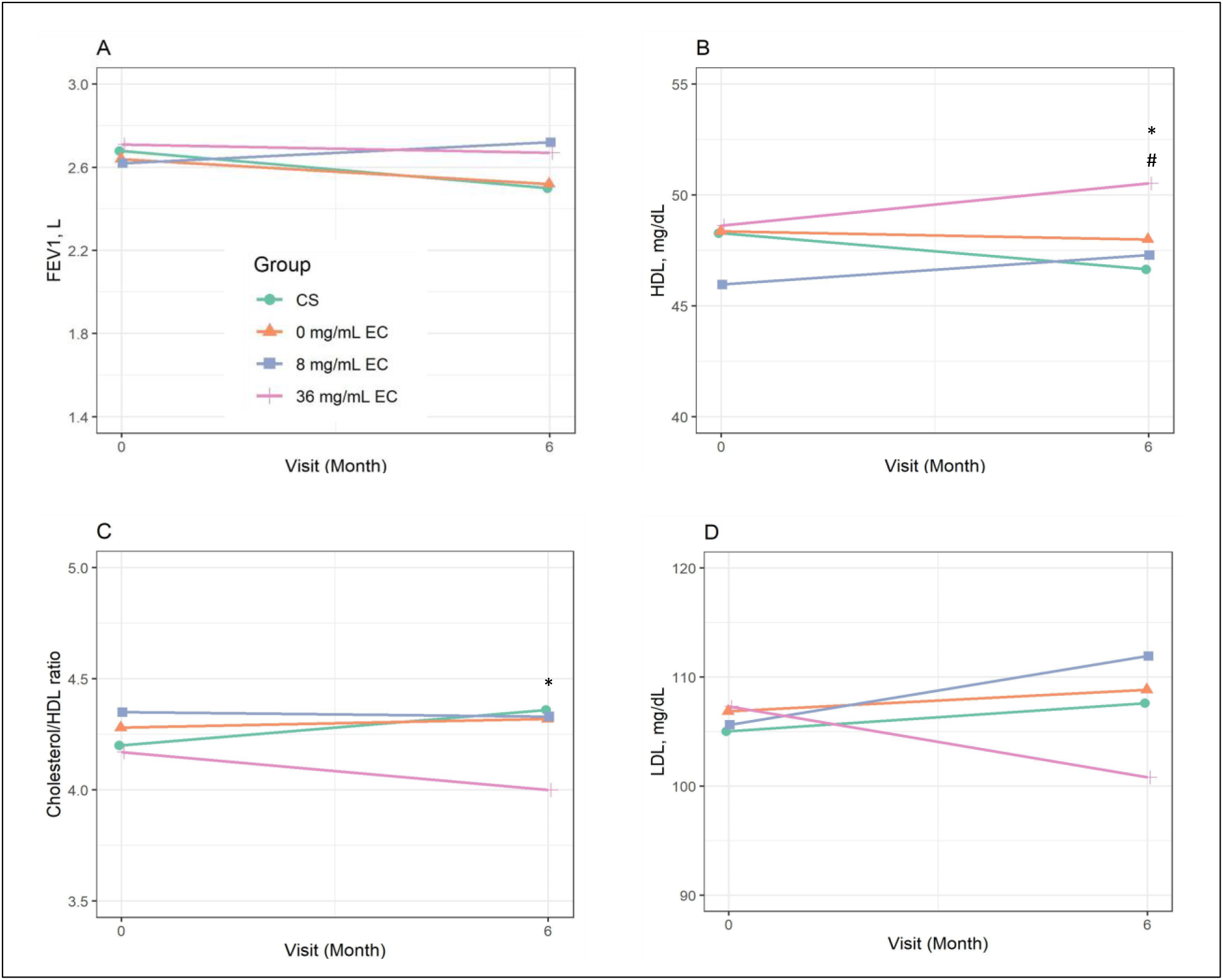
(A – D). Unadjusted mean of FEV1 (A), HDL (B), Cholesterol/HDL ratio (C), and LDL (D) at baseline and at 6 months post intervention across randomized groups; * indicate a significant difference between the 36 mg/mL group relative to the cigarette substitute group in unadjusted intention-to-treat analysis at 6 months (*p* < 0.05). # indicate a significant difference between the 36 mg/mL group relative to the 0 mg/mL EC group in unadjusted intention-to-treat analysis at 6 months (*p* < 0.05).

**Table 2.** Results from One-Way ANOVAs comparing outcome measurements at baseline by overall and conditions.

| Characteristics | E-cigarette Conditions |  |  |  | Overall<br>(N=520) | *p*-value |
| --- | --- | --- | --- | --- | --- | --- |
|  | CS<br>(n=130) | 0 mg/mL<br>(n=130) | 8 mg/mL<br>(n=130) | 36 mg/mL<br>(n=130) |  |  |
|  | Mean<br>(SD) | Mean (SD) | Mean (SD) | Mean (SD) |  |  |
| **Pulmonary Function Tests, N=518** |  |  |  |  |  |  |
| FEV1, L | 2.7 (0.9) | 2.6 (0.9) | 2.6 (0.8) | 2.7 (0.9) | 2.7 (0.9) | 0.833 |
| FEV1 % pred | 80.5 (18.5) | 79.6 (19.8) | 77.4 (17.6) | 81.2 (19.0) | 79.7 (18.7) | 0.389 |
| FVC, L | 3.4 (1.1) | 3.4 (1.0) | 3.4 (1.0) | 3.4 (1.0) | 3.4 (1.0) | 0.916 |
| FVC % pred | 82.0 (19.3) | 82.2 (18.6) | 79.1 (16.1) | 82.8 (18.5) | 81.5 (18.2) | 0.372 |
| FEV1/FVC,<br>N=517 | 79.1 (9.6) | 77.7 (10.3) | 77.9 (9.5) | 78.9 (9.8) | 78.4 (9.8) | 0.573 |
| FEV1/FVC % pred | 91.1 (12.7) | 90.5 (11.9) | 91.2 (11.9) | 91.0 (12.3) | 91.0 (12.2) | 0.972 |
| PEF L/s | 6.1 (1.9) | 6.0 (1.9) | 6.1 (1.7) | 6.1 (1.9) | 6.1 (1.9) | 0.963 |
| PEF % pred | 76.9 (21.6) | 76.4 (20.5) | 75.9 (19.1) | 77.8 (22.5) | 76.7 (20.9) | 0.893 |
| FEF 25 L/s | 5.2 (2.0) | 5.0 (1.9) | 5.1 (1.9) | 5.2 (1.9) | 5.1 (1.9) | 0.763 |
| FEF 75 L/s | 1.2 (0.7) | 1.1 (0.7) | 1.1 (0.6) | 1.2 (0.6) | 1.2 (0.7) | 0.349 |
| FEF 25-75 L/s | 2.6 (1.2) | 2.5 (1.2) | 2.5 (1.2) | 2.6 (1.1) | 2.6 (1.2) | 0.789 |
| FEF 25-75 % pred | 80.0 (29.1) | 77.0 (31.6) | 75.0 (31.5) | 79.2 (29.1) | 77.8 (30.4) | 0.546 |
| FET, sec N=504 | 4.8 (1.7) | 4.7 (1.7) | 4.5 (1.5) | 4.5 (1.7) | 4.6 (1.7) | 0.522 |
| **Clinical COPD Questionnaire (CCQ)** |  |  |  |  |  |  |
| COPD Symptoms,<br>N=514 | 1.7 (1.0) | 1.8 (1.1) | 2.0 (1.1) | 1.8 (1.0) | 1.8 (1.1) | 0.185 |
| COPD Functional<br>State, N=512 | 0.8 (1.1) | 0.7 (0.9) | 0.7 (1.0) | 0.6 (0.8) | 0.7 (1.0) | 0.221 |
| COPD Mental<br>State, N=508 | 1.1 (1.3) | 1.0 (1.1) | 1.2 (1.2) | 1.1 (1.3) | 1.1 (1.2) | 0.792 |
| COPD Total Score,<br>N=506 | 1.2 (0.9) | 1.2 (0.9) | 1.3 (1.0) | 1.2 (0.8) | 1.2 (0.9) | 0.739 |
| **Complete Metabolic Panel, N=496** |  |  |  |  |  |  |
| Sodium, mmol/L | 139.2 (2.9) | 139.7 (3.1) | 139.6 (2.8) | 139.1 (3.1) | 139.4 (3.0) | 0.346 |
| Potassium, mmol/L | 4.0 (0.3) | 4.1 (0.4) | 4.0 (0.3) | 4.1 (0.5) | 4.1 (0.4) | 0.194 |
| Chloride, mmol/L | 103.2 (3.1) | 103.5 (3.0) | 103.8 (3.0) | 103.1 (3.7) | 103.4 (3.2) | 0.302 |
| Carbon dioxide,<br>mmol/L | 26.8 (2.8) | 26.6 (2.5) | 26.6 (2.3) | 26.1 (2.4) | 26.5 (2.5) | 0.186 |
| Anion gap,<br>mmol/L | 9.2 (3.5) | 9.6 (3.8) | 9.2 (3.4) | 9.9 (3.4) | 9.5 (3.5) | 0.299 |
| Glucose, mg/dL | 108.3<br>(47.4) | 105.1 (43.2) | 106.0 (37.7) | 111.8 (67.5) | 107.8 (50.2) | 0.721 |
| BUN, mg/dL | 12.7 (4.0) | 12.3 (4.1) | 13.1 (4.4) | 12.7 (4.3) | 12.7 (4.2) | 0.528 |
| AST, units/L | 32.4 (22.0) | 33.3 (23.1) | 33.4 (21.4) | 33.8 (19.2) | 33.2 (21.4) | 0.964 |
| ALT, units/L | 30.7 (22.4) | 29.8 (26.3) | 33.2 (27.8) | 32.4 (20.1) | 31.5 (24.3) | 0.691 |
| ALP, units/L | 79.9 (24.7) | 80.1 (28.3) | 81.5 (24.0) | 76.3 (23.6) | 79.4 (25.2) | 0.408 |
| Bilirubin, mg/dL | 0.3 (0.2) | 0.4 (0.3) | 0.4 (0.2) | 0.3 (0.2) | 0.3 (0.2) | 0.898 |
| Protein, mg/dL | 7.4 (0.5) | 7.4 (0.5) | 7.4 (0.6) | 7.4 (0.5) | 7.4 (0.5) | 0.649 |
| Albumin, mg/dL | 4.3 (0.3) | 4.2 (0.3) | 4.2 (0.3) | 4.3 (0.3) | 4.2 (0.3) | 0.446 |
| Calcium, mg/dL | 9.4 (0.4) | 9.3 (0.4) | 9.3 (0.5) | 9.3 (0.4) | 9.3 (0.4) | 0.192 |
| CRP, mg/dL, | 0.8 (0.8) | 0.7 (0.5) | 0.7 (0.5) | 0.7 (0.5) | 0.7 (0.6) | 0.451 |
| N=495 |  |  |  |  |  |  |
| Creatinine, mg/dL,<br>N=520 | 122.4<br>(98.6) | 118.2 (83.7) | 96.9 (71.0) | 108.9 (91.6) | 111.6 (87.1) | 0.392 |
| **Complete Blood Count, N=494** |  |  |  |  |  |  |
| WBC, 10<sup>9</sup>/L | 7.7 (2.4) | 8.1 (2.4) | 7.9 (2.3) | 7.8 (2.1) | 7.9 (2.3) | 0.642 |
| RBC, 10<sup>12</sup>/L | 4.6 (0.5) | 4.7 (0.4) | 4.8 (0.5) | 4.6 (0.5) | 4.7 (0.5) | 0.018* |
| Hemoglobin, g/dL | 14.0 (1.5) | 14.2 (1.2) | 14.4 (1.5) | 14.1 (1.4) | 14.2 (1.4) | 0.256 |
| Hematocrit, % | 41.6 (3.9) | 42.5 (3.4) | 42.9 (3.9) | 42.0 (3.6) | 42.2 (3.7) | 0.033* |
| MCV, fL | 89.8 (5.4) | 90.3 (5.9) | 90.0 (5.7) | 91.5 (5.7) | 90.4 (5.7) | 0.077 |
| MCH, pg | 30.3 (2.3) | 30.2 (2.5) | 30.1 (2.4) | 30.7 (2.3) | 30.3 (2.4) | 0.208 |
| MCHC, g/dL | 33.7 (1.2) | 33.4 (1.1) | 33.4 (1.2) | 33.5 (1.1) | 33.5 (1.2) | 0.273 |
| RDW, % | 13.3 (1.2) | 13.4 (1.4) | 13.4 (1.5) | 13.2 (1.3) | 13.3 (1.3) | 0.556 |
| Platelets, 10<sup>9</sup>/L | 252.5<br>(64.8) | 252.6 (61.8) | 254.4 (58.9) | 257.9 (59.8) | 254.3 (61.2) | 0.889 |
| MPV, fL | 10.2 (1.5) | 10.2 (1.2) | 10.2 (1.2) | 10.2 (1.2) | 10.2 (1.3) | 0.944 |
| **Lipoproteins, N=496** |  |  |  |  |  |  |
| Cholesterol, mg/dL | 188.3<br>(38.6) | 189.0 (44.9) | 186.1 (42.1) | 187.3 (37.9) | 187.7 (40.8) | 0.952 |
| HDL, mg/dL | 48.3 (17.1) | 48.4 (16.1) | 46.0 (13.4) | 48.6 (14.7) | 47.8 (15.4) | 0.495 |
| Non-HDL<br>Cholesterol, mg/dL | 140.0<br>(38.3) | 140.6 (47.8) | 140.2 (43.5) | 138.7 (39.2) | 139.9 (42.2) | 0.986 |
| Cholesterol/HDL<br>Ratio | 4.2 (1.3) | 4.3 (1.8) | 4.4 (1.5) | 4.2 (1.5) | 4.3 (1.5) | 0.804 |
| LDL Calculated,<br>mg/dL (N=476) | 105.0<br>(33.7) | 106.9 (36.1) | 105.6 (33.6) | 107.3 (37.3) | 106.2 (35.1) | 0.955 |
| Triglycerides,<br>mg/dL | 177.2<br>(117.5) | 171.6<br>(148.2) | 186.1<br>(160.3) | 169.9<br>(125.6) | 176.2<br>(138.6) | 0.794 |
| **Other Physiological Measures** |  |  |  |  |  |  |
| Weight, pound,<br>N=520 | 193.5<br>(50.7) | 188.8 (58.0) | 194.0 (50.8) | 188.4 (44.3) | 191.2 (51.1) | 0.722 |
| BMI, kg/m<sup>2</sup>,<br>N=519 | 30.3 (7.2) | 29.7 (8.1) | 30.2 (7.4) | 29.5 (6.6) | 29.9 (7.3) | 0.801 |
| Heart rate, bpm,<br>N=519 | 81.4 (13.0) | 83.0 (14.1) | 81.9 (12.8) | 80.9 (12.4) | 81.8 (13.1) | 0.579 |
| Systolic blood<br>pressure, mmHg,<br>N=519 | 125.3<br>(14.0) | 124.3 (13.8) | 127.1 (15.5) | 124.6 (14.5) | 125.3 (14.5) | 0.428 |
| Diastolic blood<br>pressure, mmHg,<br>N=519 | 78.6 (9.9) | 79.6 (9.7) | 80.3 (9.8) | 78.3 (10.1) | 79.2 (9.9) | 0.316 |
| Waist-to-hip ratio,<br>N=518 | 0.9 (0.1) | 0.9 (0.1) | 0.9 (0.1) | 0.9 (0.1) | 0.9 (0.1) | 0.898 |
| INTERHEART<br>risk score, N=519 | 20.6 (5.1) | 20.2 (5.6) | 21.0 (6.2) | 20.6 (5.8) | 20.6 (5.7) | 0.766 |

Note: CS, cigarette substitute; FEV1, forced expiratory volume in 1 second; FVC, forced vital capacity; PEF, peak expiratory flow; FEF, forced expiratory flow; FET, forced expiratory time; BUN, blood urea nitrogen; AST, aspartate aminotransferase; ALT, alkaline transferase; ALP, alkaline phosphatase; CRP, C-reactive Protein; WBC, white blood cells; RBC, red blood cells; MCV, mean corpuscular volume; MCH, mean corpuscular hemoglobin; MCHC, mean corpuscular hemoglobin concentration; RDW, red cell distribution width; MPV, mean platelet volume; HDL, high-density lipoprotein; LDL, low-density lipoprotein; COPD, chronic obstructive pulmonary disorder; BMI, body mass index.
*p*-values were reported from one-way ANOVA.
\*: *p*-value < 0.05, Reference group: CS

**Table 3.**
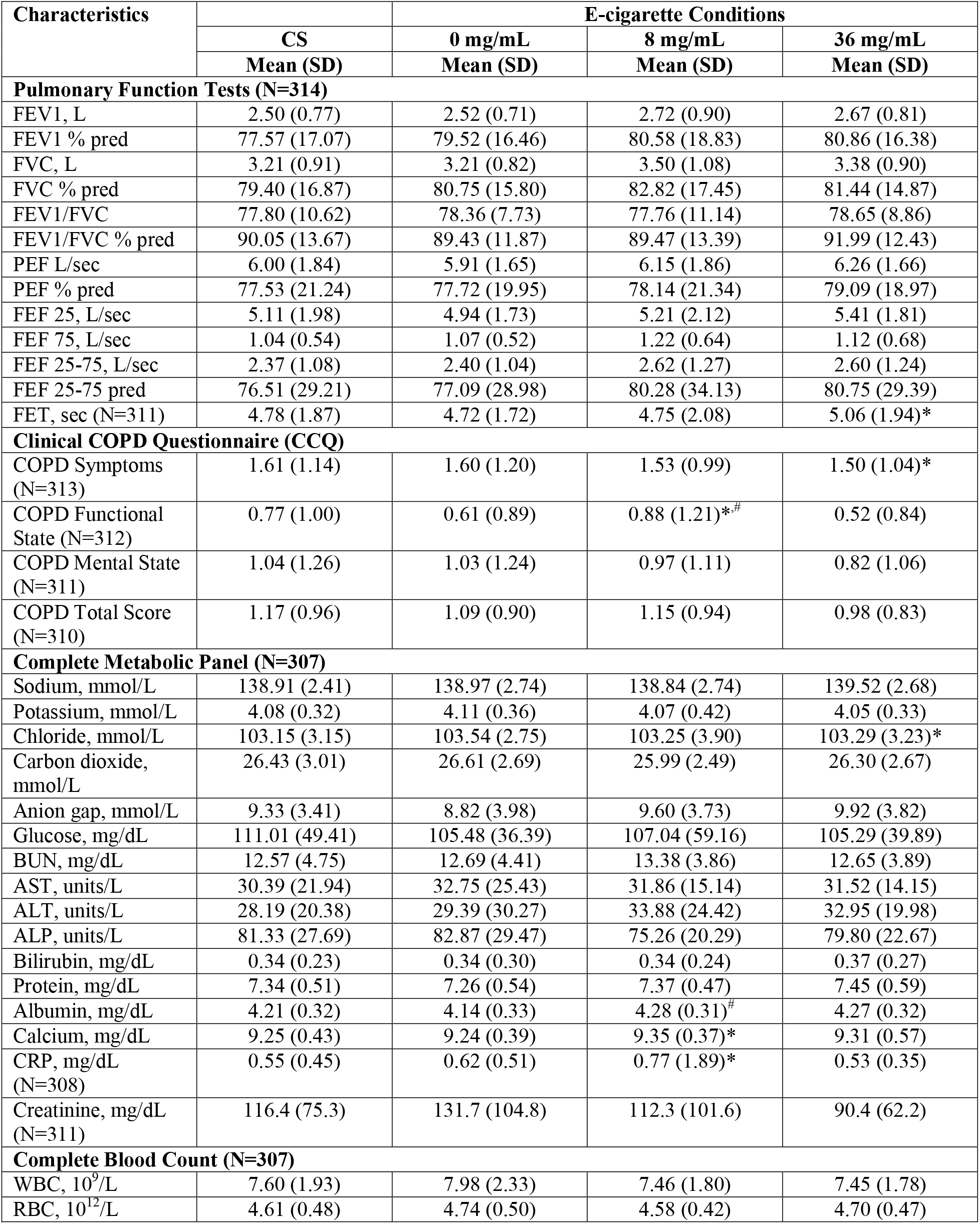
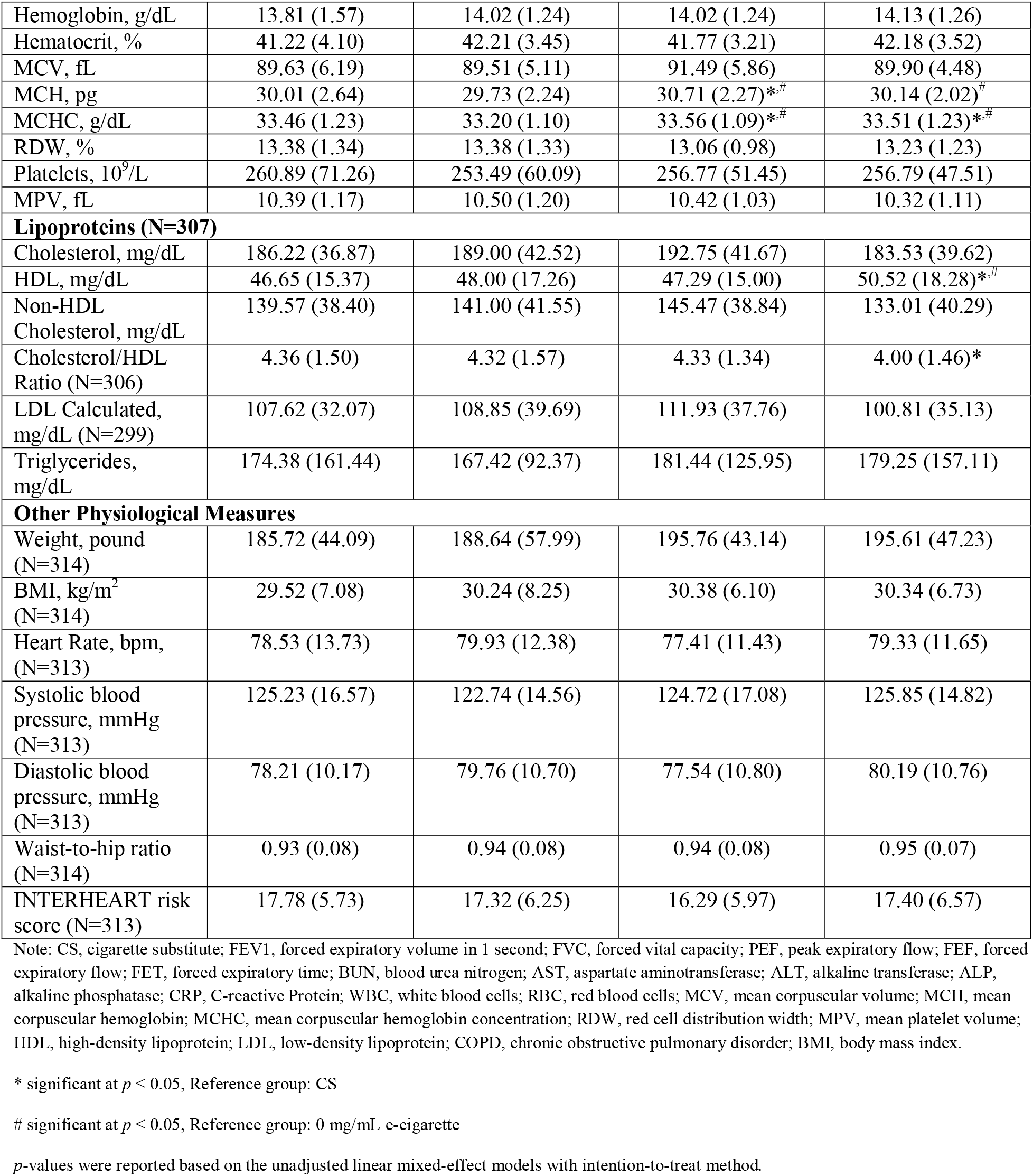
Results from linear mixed-effects models comparing outcome measurements at the end of intervention by conditions (6-month)

There were no significant differences across the randomized conditions for total CCQ score or any individual CCQ domain (symptoms, functional and mental state) at baseline (Table 2). At 6-month, participants using 36 mg/mL nicotine e-cigarettes had significantly lower COPD symptoms as compared to CS group in the unadjusted analysis (*p*=0.021; Table 3; Supplemental Figure 15 – A, C). There were no significant differences across the randomized conditions in COPD mental state domain and COPD total score at the 6-month follow-up assessment (Table 3).

### Blood measurements

#### Complete blood count

At baseline, there were significant differences across the randomized groups in RBC count (*p*=0.018) and hematocrit levels (*p*=0.033; Table 2). At 6-month, participants randomized to the 8 mg/mL e-cigarettes had higher levels of MCH as compared to CS (*p*=0.019) or 0 mg/mL e-cigarettes (*p*=0.004) in unadjusted analysis (Table 3), and this effect remained significant after controlling for other factors (Supplemental Figure 40). Also, MCHC levels were higher among participants randomized to the 8 or 36 mg/mL e-cigarettes as compared to either CS or 0 mg/mL e-cigarettes in both unadjusted and adjusted analysis (Table 3; Supplemental Figure 41).

#### Complete metabolic panel, CRP and lipoproteins

At baseline, there were no significant differences between conditions for complete metabolic panel, CRP, or lipoprotein measures (Table 2). At 6-month, participants randomized to the 36 mg/mL e-cigarettes had significantly higher levels of HDL as compared to participants randomized to the CS or 0 mg/mL e-cigarettes in both the unadjusted (Figure 1B; Table 3) and adjusted analysis (Supplementary Figure 46 – B, D). Cholesterol/HDL ratio was significantly lower among participants randomized to 36 mg/mL e-cigarettes as compared to participants in CS condition (*p*=0.034; Table 3; Figure 1C). The direction and magnitude of the differences were similar in the unadjusted and adjusted analysis (Supplementary Figure 48). Similarly, LDL levels were significantly lower among participants randomized to 36 mg/mL e-cigarettes as compared to CS condition in adjusted analysis (*p*=0.044; Supplemental Figure 49 – B, D), although it was not significant in unadjusted analysis (Figure 1D; Supplemental Figure 49 – A, C).

#### Other physiological measures

At baseline and 6-month, no significant group differences were seen between e-cigarette and CS conditions for all physiological measures (weight, BMI, heart rate, systolic and diastolic BP, waist-to-hip ratio, and INTERHEART risk score) in either the unadjusted or adjusted analysis (Table 2, Table 3, and Supplemental Figures 51 – 57). Similarly, no differences were observed when comparing 0 mg/mL condition to 8 or 36 mg/mL nicotine-containing e-cigarette conditions on these physiological measures (Table 3; Supplemental Figures 51 – 57).

## DISCUSSION

In this four-arm, prospective, parallel-group, RCT, we examined differences in CVD risk factors, pulmonary function, and other standard clinical laboratory markers among participants who smoked cigarettes regularly and were instructed to reduce their cigarette smoking with one of four study products (e-cigarettes with 0, 8 or 36 mg/mL nicotine or a CS). Compared with participants in the CS or non-nicotine e-cigarette group, we found no consistent patterns of harm or benefit from nicotine e-cigarette use on most of the measured outcomes. We observed a statistically significant improvement in HDL levels, cholesterol/HDL ratio, LDL levels and COPD symptoms among participants using the 36 mg/mL nicotine e-cigarettes compared to CS. These results were consistent in both the unadjusted and adjusted analyses for HDL levels, cholesterol/HDL ratio and COPD symptoms though between group differences were small with limited clinical significance. Further, as this study is exploratory in nature, these significant findings must be interpreted cautiously and verified in follow-up studies.

We found no clinically significant between group differences in PFT measures at 6-month. These results are similar to other RCTs that explored the acute (<7 days)^33^, short to mid-term (from 8 days to 12 months)^17,18,25^ or long-term (more than 12 months)^19^ effects of e-cigarette use on spirometry indices among people who partially or completely switched from TCs to e-cigarettes. At 6-month follow-up, the forced expiratory time (FET) was significantly longer among participants who were using the 36 mg/mL nicotine e-cigarettes as compared to CS indicating longer time in lung emptying. FET has long been proposed as an early marker of small airway dysfunction.^34^ More recent investigators emphasize the interindividual variation in FET determined by two competing effects: airway narrowing that increases the duration of expiration and airway closure that terminates it.^35^ FET becomes longer with age and among those who smoke, and this effect may be differentially impacted by obstructive and restrictive processes.^35^ In our study, other spirometry indices such as FVC, FEV1/FVC, and FEF 25–75 were descriptively higher in the 36 mg/mL nicotine e-cigarette group as compared to CS group, suggesting that the prolonged FET was not an indicator of physiologic dysfunction.

The purpose of the present study was to explore the impact of nicotine liquid concentration in e-cigarettes on clinical health outcomes across a 6-month period, which has been understudied in the current literature. This detailed analysis could help to identify harm related to e-cigarette use among participants trying to reduce their TC consumption. In effect, we found no statistically or clinically significant changes in spirometry measures and most of the CVD risk factors (i.e., CRP, SBP, DBP, heart rate, BMI, waist-to-hip ratio, triglycerides) across the randomized conditions despite evidence from other outcomes from this same trial that support the utility of the 36 mg/mL nicotine e-cigarettes in reducing TC use and associated toxicant exposure as compared to other conditions.^26^ The methodological differences in study designs, control groups (i.e., variations in smoking/e-cigarette use history), and follow-up time periods (i.e., acute or long-term effects) in the related evidence from prior reports presents challenges for the comparison of the study findings.^11,13,17,18,21,36,37^

Of interest, we observed in the current study that participants using the 36 mg/mL nicotine e-cigarettes had significantly lower self-reported COPD symptoms (i.e., shortness of breath at rest or during physical activities, cough, phlegm production), higher HDL levels (“good cholesterol”), lower LDL levels (“bad cholesterol”), and lower total cholesterol/HDL ratio (indicator of risk of getting cardiovascular disease) as compared to the CS group. Findings from our primary outcome paper indicate that the 36 mg/mL e-cigarette group reduced their cigarette consumption by 58% and the CS group reduced by 39% at 6-month.^26^ The abstinence rate was also significantly higher in the 36 mg/mL group as compared to the CS group at 6-month (10.8% vs. 3.1%).^27^ Therefore, a plausible explanation for the positive changes may be the sustained reduction in cigarette smoking and carbon monoxide exposure, and greater abstinence rate among those in the 36 mg/mL nicotine e-cigarette group as compared to the CS group.

Similar to our study findings, a one-year RCT demonstrated substantial improvement in respiratory symptoms (shortness of breath and cough/phlegm production) among individuals who quit or reduced their TC consumption by switching to e-cigarettes as compared to those who failed to reduce TC use (<50% reduction in the CPD from baseline).^19^ Studies have shown that low levels of HDL, and high levels of LDL and total cholesterol/HDL ratio have been associated with atherosclerosis and elevated risk of developing CVD.^38,39^ In addition, smoking cessation leads to an increase in cardio-protective HDL levels.^40–43^ People who smoke and are more addicted, as indicated by time to first cigarettes have less favorable lipid indices.^44^ The available evidence and the current study findings suggest that high nicotine e-cigarettes may better facilitate reductions in TC smoking and beneficial changes in lipid parameters, which may mediate reduction in CVD risk among participants. However, the results must be interpreted cautiously due to small effect size and limited literature on changes in lipid profile among dual-users trying to reduce TC use. Although statistically significant, the mean of HDL, LDL and cholesterol/HDL ratio across four randomized arms falls in the normal reference range for clinical interpretation. Further investigation is recommended to determine whether this might be a transient phenomenon or translates to a more sustained health benefit.

Participants randomized to nicotine e-cigarettes had significantly higher levels of MCHC (amount of hemoglobin per unit volume) and MCH (amount of hemoglobin per red blood cell) (only for 8 mg/mL e-cigarette group) as compared to the two non-nicotine groups. Also, the 36 mg/mL group had significantly higher MCH than the placebo group. Elisia et al. (2020) found that both MCH and MCHC were elevated for those who smoked versus those who did not,^45^ which is contrary to our finding as these measures were elevated in nicotine e-cigarette groups that smoked less and had significantly lower CO measurements.^26^ As the differences across groups were very small and the values fall in normal reference range across each group, the mechanism behind these differences remains unclear, but decreased CO exposure after reducing TC leading to decrease in carboxyhemoglobin may be involved.^46^ There were no clinically meaningful between group differences in most of the other laboratory measurements included in standard blood work panels.

Overall, our findings did not show additional harm among individuals using 8 and 36 mg/mL nicotine e-cigarettes to reduce TC smoking compared to CS or 0 mg/mL e-cigarette use. Contrary to our findings, other studies have found adverse changes at the cellular level using e-cigarettes, such as, airway hyperresponsiveness, sympathetic activation, vascular stiffening, endothelial dysfunction, and atherosclerotic plaque formation in human and animal models.^12,14,47^ The variability in the findings might be due to different factors such as the e-cigarette device and liquid used, the composition of e-cigarette aerosol, and individual use patterns, as they may influence the effects on cells/tissues and blood and urinary markers. Future studies with long-term follow-up (> 1 year) and methodological similarities are required.

The major strengths of this study are the use of a randomized, double-blind design which included both a placebo and a non-e-cigarette control group, the use of an e-cigarette with a well characterized nicotine delivery profile,^48^ a large and diverse sample, and a 6 month intervention period. Also, the study encompassed a comprehensive battery of clinically relevant biological measures (e.g., pulmonary function, CVD risk, and clinical laboratory markers) to examine changes due to e-cigarette use among people trying to reduce their TC consumption. The study’s limitations included participant drop-out (36%) and use of one type of e-cigarette. In addition, the findings are only generalizable to individuals who are interested in reducing their TC smoking. Multiple comparison adjustments were not done as this is an exploratory analysis aiming to identify potential changes in clinical health outcomes and generate hypotheses for future research.

## CONCLUSION

The current RCT found that nicotine-containing e-cigarette use during 6 months of TC smoking reduction did not contribute to additional harms as compared to a plastic CS or a non-nicotine e-cigarette group. This study examined 56 physiological and related measures of CVD risk, pulmonary function, and standard clinical laboratory tests. A novel finding that warrants further exploration was the improvement in blood lipids (e.g., increased HDL, reduced LDL and cholesterol/HDL ratio) in participants assigned to use the 36 mg/mL nicotine e-cigarettes. While the magnitude of these changes was small, future studies should further examine the impact of e-cigarette use on lipid profiles and CVD risk reduction.

## Contributors

SD: formal analysis, writing-original draft, writing-review and editing, guarantor. JY: conceptualisation, methodology, investigation, data curation, writing-review and editing, supervision. XW: formal analysis, writing-review and editing. COC: conceptualisation, methodology, investigation, data curation, writing-review and editing. MC: writing-review and editing. SH: writing-review and editing. RB: writing-review and editing. AAL: writing-review and editing. LK: writing-review and editing. SM: writing-review and editing. MH: writing-review and editing. JF: conceptualisation, methodology, writing-review and editing, funding acquisition, supervision. SV: conceptualisation, methodology, investigation, data curation, writing-review and editing, supervision.

## Disclaimer

The content is solely the responsibility of the authors and does not necessarily represent the official views of the National Institutes of Health or the Food and Drug Administration.

## Data availability statement

Data are available on reasonable request. On publication, requests for deidentified individual participant data or study documents (eg, data dictionary; protocol; statistical analysis plan; measures, manuals or informed consent documentation) will be considered. The requestor must submit a one-page abstract of their proposed research, including the purpose, analytical plan and dissemination plans. The Executive Leadership Committee (Virginia Commonwealth University and Penn State University) will review the abstract and decide on the basis of the individual merits. Review criteria and prioritization of projects include potential of the proposed work to advance public health, qualifications of the applicant, the potential for publication, the potential for future funding, and enhancing the scientific, geographic and demographic diversity of the research portfolio. Following abstract approval, requestors must receive institutional ethics approval or confirmation of exempt status for the proposed research. An executed data use agreement must be completed before data distribution. Requests should be made to the corresponding author.

## REFERENCES

1. Adjaye-Gbewonyo D. National Health Interview Survey Early Release Program. 2021

2. Norris T. Early Release of Selected Estimates Based on Data From the 2023 National Health Interview Survey. Published online 2023.

3. Breland A, Soule E, Lopez A, et al. Electronic cigarettes: what are they and what do they do? Ann N Y Acad Sci. 2017;1394(1):5–30. doi:10.1111/nyas.12977

4. Samet JM, Barrington-Trimis J. E-Cigarettes and Harm Reduction: An Artificial Controversy Instead of Evidence and a Well-Framed Decision Context. Am J Public Health. 2021;111(9):1572–1574. doi:10.2105/AJPH.2021.306457

5. Centers for Disease Control and Prevention. About electronic cigarettes (e-cigarettes). 2023 https://www.cdc.gov/tobacco/basic_information/e-cigarettes/about-e-cigarettes.html

6. Hartmann-Boyce J, McRobbie H, Butler AR, et al. Electronic cigarettes for smoking cessation. Cochrane Database Syst Rev. 2021;2021(4):CD010216. doi:10.1002/14651858.CD010216.pub5

7. Soneji S, Barrington-Trimis JL, Wills TA, et al. Association Between Initial Use of e-Cigarettes and Subsequent Cigarette Smoking Among Adolescents and Young Adults: A Systematic Review and Meta-analysis. JAMA Pediatr. 2017;171(8):788–797. doi:10.1001/jamapediatrics.2017.1488

8. U.S. Department of Health and Human Services. E-Cigarette Use Among Youth and Young Adults. A Report of the Surgeon General. Atlanta, GA: U.S. Department of Health and Human Services, Centers for Disease Control and Prevention, National Center for Chronic Disease Prevention and Health Promotion, Office on Smoking and Health, 2016.

9. Seiler-Ramadas R, Sandner I, Haider S, et al. Health effects of electronic cigarette (e cigarette) use on organ systems and its implications for public health. Wien Klin Wochenschr. 2021;133(19-20):1020–1027. doi:10.1007/s00508-020-01711-z

10. Ali N, Xavier J, Engur M, et al. The impact of e-cigarette exposure on different organ systems: A review of recent evidence and future perspectives. J Hazard Mater. 2023;457:131828. doi:10.1016/j.jhazmat.2023.131828

11. McNeill A, Brose L, Robson D, et al. Nicotine vaping in England: An evidence update including health risks and perceptions, 2022. Published online September 29, 2022. Accessed March 23, 2023. https://kclpure.kcl.ac.uk/portal/en/publications/nicotine-vaping-in-england(58869843-3d0c-40e7-a9e9-d92881a2ed22).html

12. Tsai M, Byun MK, Shin J, et al. Effects of e cigarettes and vaping devices on cardiac and pulmonary physiology. J Physiol. 2020;598(22):5039–5062. doi:10.1113/JP279754

13. Dimitriadis K, Narkiewicz K, Leontsinis I, et al. Acute Effects of Electronic and Tobacco Cigarette Smoking on Sympathetic Nerve Activity and Blood Pressure in Humans. Int J Environ Res Public Health. 2022;19(6):3237. doi:10.3390/ijerph19063237

14. Marczylo T. How bad are e-cigarettes? What can we learn from animal exposure models? J Physiol. 2020;598(22):5073–5089. doi:10.1113/JP278366

15. Keith R, Bhatnagar A. Cardiorespiratory and Immunologic Effects of Electronic Cigarettes. Curr Addict Rep. 2021;8(2):336–346. doi:10.1007/s40429-021-00359-7

16. Qureshi MA, Vernooij RWM, La Rosa GRM, et al. Respiratory health effects of e-cigarette substitution for tobacco cigarettes: a systematic review. Harm Reduct J. 2023;20(1):1–14. doi:10.1186/s12954-023-00877-9

17. Pulvers K, Nollen NL, Rice M, et al. Effect of Pod e-Cigarettes vs Cigarettes on Carcinogen Exposure Among African American and Latinx Smokers: A Randomized Clinical Trial. JAMA Netw Open. 2020;3(11):e2026324. doi:10.1001/jamanetworkopen.2020.26324

18. Veldheer S, Yingst J, Midya V, et al. Pulmonary and other health effects of electronic cigarette use among adult smokers participating in a randomized controlled smoking reduction trial. Addict Behav. 2019;91:95–101. doi:10.1016/j.addbeh.2018.10.041

19. Cibella F, Campagna D, Caponnetto P, et al. Lung function and respiratory symptoms in a randomized smoking cessation trial of electronic cigarettes. Clin Sci. 2016;130(21):1929–1937. doi:10.1042/CS20160268

20. Klonizakis M, Gumber A, McIntosh E, et al. Short-Term Cardiovascular Effects of E-Cigarettes in Adults Making a Stop-Smoking Attempt: A Randomized Controlled Trial. Biology. 2021;10(11):1208. doi:10.3390/biology10111208

21. George J, Hussain M, Vadiveloo T, et al. Cardiovascular Effects of Switching From Tobacco Cigarettes to Electronic Cigarettes. J Am Coll Cardiol. 2019;74(25):3112–3120. doi:10.1016/j.jacc.2019.09.067

22. Skotsimara G, Antonopoulos AS, Oikonomou E, et al. Cardiovascular effects of electronic cigarettes: A systematic review and meta-analysis. Eur J Prev Cardiol. 2019;26(11):1219–1228. doi:10.1177/2047487319832975

23. Wang Y, Zhu Y, Chen Z, et al. Association between electronic cigarettes use and whole blood cell among adults in the USA—a cross-sectional study of National Health and Nutrition Examination Survey analysis. Environ Sci Pollut Res. 2022;29(59):88531–88539. doi:10.1007/s11356-022-21973-6

24. Flouris AD, Poulianiti KP, Chorti MS, et al. Acute effects of electronic and tobacco cigarette smoking on complete blood count. Food Chem Toxicol. 2012;50(10):3600–3603. doi:10.1016/j.fct.2012.07.025

25. Cravo AS, Bush J, Sharma G, et al. A randomised, parallel group study to evaluate the safety profile of an electronic vapour product over 12 weeks. Regul Toxicol Pharmacol. 2016;81:S1–S14. doi:10.1016/j.yrtph.2016.10.003

26. Cobb CO, Foulds J, Yen MS, et al. Effect of an electronic nicotine delivery system with 0, 8, or 36 mg/mL liquid nicotine versus a cigarette substitute on tobacco-related toxicant exposure: a four-arm, parallel-group, randomised, controlled trial. Lancet Respir Med. 2021;9(8):840–850. doi:10.1016/S2213-2600(21)00022-9

27. Foulds J, Cobb CO, Yen MS, et al. Effect of Electronic Nicotine Delivery Systems on Cigarette Abstinence in Smokers With No Plans to Quit: Exploratory Analysis of a Randomized Placebo-Controlled Trial. Nicotine Tob Res Off J Soc Res Nicotine Tob. 2022;24(7):955–961. doi:10.1093/ntr/ntab247

28. Yingst J, Wang X, Lopez AA, et al. Changes in Nicotine Dependence Among Smokers Using Electronic Cigarettes to Reduce Cigarette Smoking in a Randomized Controlled Trial. Nicotine Tob Res. 2023;25(3):372–378. doi:10.1093/ntr/ntac153

29. Lopez AA, Cobb CO, Yingst JM, et al. A transdisciplinary model to inform randomized clinical trial methods for electronic cigarette evaluation. BMC Public Health. 2016;16(1):217. doi:10.1186/s12889-016-2792-8

30. van der Molen T, Willemse BW, Schokker S, et al. Development, validity and responsiveness of the Clinical COPD Questionnaire. Health Qual Life Outcomes. 2003;1:13. doi:10.1186/1477-7525-1-13

31. Yusuf S, Hawken S, Ounpuu S, et al. Effect of potentially modifiable risk factors associated with myocardial infarction in 52 countries (the INTERHEART study): case-control study. Lancet Lond Engl. 2004;364(9438):937–952. doi:10.1016/S0140-6736(04)17018-9

32. van Buuren S. Multiple imputation of discrete and continuous data by fully conditional specification. Stat Methods Med Res. 2007;16(3):219–242. doi:10.1177/0962280206074463

33. D’Ruiz CD, O’Connell G, Graff DW, et al. Measurement of cardiovascular and pulmonary function endpoints and other physiological effects following partial or complete substitution of cigarettes with electronic cigarettes in adult smokers. Regul Toxicol Pharmacol. 2017;87:36–53. doi:10.1016/j.yrtph.2017.05.002

34. Cochrane GM, Benatar SR, Davis J, et al. Correlation between tests of small airway function. Thorax. 1974;29(2):172–178. doi:10.1136/thx.29.2.172

35. Skloot GS, O’Connor-Chapman KL, Schechter CB, et al. Forced expiratory time: a composite of airway narrowing and airway closure. J Appl Physiol. 2021;130(1):80–86. doi:10.1152/japplphysiol.00556.2020

36. Polosa R, Cibella F, Caponnetto P, et al. Health impact of E-cigarettes: a prospective 3.5-year study of regular daily users who have never smoked. Sci Rep. 2017;7(1):13825. doi:10.1038/s41598-017-14043-2

37. Tattersall MC, Hughey CM, Piasecki TM, et al. Cardiovascular and Pulmonary Responses to Acute Use of Electronic Nicotine Delivery Systems and Combustible Cigarettes in Long-term Users. Chest. Published online April 2023:S0012369223004944. doi:10.1016/j.chest.2023.03.047

38. Zhou Z, Ong KL, Whelton SP, et al. Impact of Blood Lipids on 10-Year Cardiovascular Risk in Individuals Without Dyslipidemia and With Low Risk Factor Burden. Mayo Clin Proc. 2022;97(10):1883–1893. doi:10.1016/j.mayocp.2022.03.025

39. Borén J, Chapman MJ, Krauss RM, et al. Low-density lipoproteins cause atherosclerotic cardiovascular disease: pathophysiological, genetic, and therapeutic insights: a consensus statement from the European Atherosclerosis Society Consensus Panel. Eur Heart J. 2020;41(24):2313–2330. doi:10.1093/eurheartj/ehz962

40. Gepner AD, Piper ME, Johnson HM, et al. Effects of smoking and smoking cessation on lipids and lipoproteins: Outcomes from a randomized clinical trial. Am Heart J. 2011;161(1):145–151. doi:10.1016/j.ahj.2010.09.023

41. Forey BA, Fry JS, Lee PN, et al. The effect of quitting smoking on HDL-cholesterol - a review based on within-subject changes. Biomark Res. 2013;1:26. doi:10.1186/2050-7771-1-26

42. Maeda K, Noguchi Y, Fukui T. The effects of cessation from cigarette smoking on the lipid and lipoprotein profiles: a meta-analysis. Prev Med. 2003;37(4):283–290. doi:10.1016/S0091-7435(03)00110-5

43. Colsoul ML, Goderniaux N, Onorati S, et al. Changes in biomarkers of endothelial function, oxidative stress, inflammation and lipids after smoking cessation: A cohort study. Eur J Clin Invest. 2023;53(8):e13996. doi:10.1111/eci.13996

44. Selya AS, Hesse ND. Time to first cigarette and serum cholesterol levels. Soc Sci Med 1982. 2017;174:213–219. doi:10.1016/j.socscimed.2016.12.014

45. Elisia I, Lam V, Cho B, et al. The effect of smoking on chronic inflammation, immune function and blood cell composition. Sci Rep. 2020;10:19480. doi:10.1038/s41598-020-76556-7

46. Fernández CS, Matute WIG, Zichen J, et al. Correlation between blood carboxyhemoglobin levels and smoking. Eur Respir J. 2022;60(suppl 66). doi:10.1183/13993003.congress-2022.3618

47. Neczypor EW, Mears MJ, Ghosh A, et al. E-Cigarettes and Cardiopulmonary Health: Review for Clinicians. Circulation. 2022;145(3):219–232. doi:10.1161/CIRCULATIONAHA.121.056777

48. Hiler M, Breland A, Spindle T, et al. Electronic cigarette user plasma nicotine concentration, puff topography, heart rate, and subjective effects: Influence of liquid nicotine concentration and user experience. Exp Clin Psychopharmacol. 2017;25(5):380–392. doi:10.1037/pha0000140

