## Supplemental Table 1-3, Supplemental Figure 1-57 for "Changes in Cardiovascular Disease Risk, Lung Function, and Other Clinical Health Outcomes When People Who Smoke Use E-cigarettes to Reduce Cigarette Smoking: An Exploratory Analysis from a Randomized Placebo-Controlled Trial"

### Supplementary Materials

**Overview of Supplementary Files**

- In this file, we summarize results of the unadjusted and adjusted analysis for outcomes of interest. For exploratory analysis, we do not adjust for multi-testing due to lack of power.

- In **unadjusted line plot**, we show means of outcome at baseline and 6 months.

- In **adjusted line plot**, we show means of outcome at baseline and 6 months **adjusting for baseline covariates**.

- The heatmap shows p-values by treatment group (y-axis) and analysis method (x-axis) to evaluate

  1) change in outcome from baseline to follow-up timepoint (6 months) **between groups** (column panel “Cig Sub”, “8 E-cig”, “36 E-cig”) and
  2) change from baseline to 1,3,6 months **within each group** (column panel “Within Group”).

  We report p-values at $\alpha = 0.05$ level.

#### Supplemental Table 1: Covariates included in fully conditional regression model

| Covariates | |
|---|---|
| **Fully conditional regression model for all outcomes** | Study site, gender, age, race/ethnicity, education, total household income, age of smoking initiation, week 0 environmental smoke score, week 0 urinary total NNAL (transformed, pg/mg creatinine), week 0 urinary cotinine (ng/mg creatinine), week 0 exhaled CO, week 0 cigarettes smoked per day, week 0 body mass index, week 0 height, week 0 weight, week 0 hip circumference, week 0 waist circumference, week 0 diastolic blood pressure, week 0 Center for Epidemiologic Studies Depression Scale score, week 0 Clinical Chronic Obstructive Pulmonary Disease score, week 0 partial INTERHEART non-laboratory score, week 0 Kessler K6 score, week 0 Perceived Stress Scale score, week 0 forced expiratory volume in one second, and week 0 forced vital capacity, week 0 Penn State Cigarette Dependence Index score |

#### Supplemental Table 2: Covariates included in adjusted analyses

| Covariates included in adjusted analyses | |
|---|---|
| **PFTs (FEV1/FVC, FEV1/FVC%, FEF 25, FEF 75, FEF 25-75, FEF 25-75%, Forced expiratory time in seconds)** | Study site, gender, age, race/ethnicity, education, age of smoking initiation, week 0 urinary total NNAL (transformed, pg/mg creatinine), week 0 urinary cotinine (transformed, ng/mg creatinine), week 0 exhaled CO, week 0 body mass index, week 0 height, week 0 weight, week 0 hip circumference, week 0 waist circumference, week 0 Center for Epidemiologic Studies depression scale score, week 0 Clinical Chronic Obstructive Pulmonary Disease score, week 0 partial INTERHEART non-laboratory score, week 0 Kessler K6 score, week 0 Penn State Cigarette Dependence Index score, week 0 Perceived Stress Scale score, week 0 forced expiratory volume in one second, week 0 forced vital capacity |
| **COPD** | Age, education, total household income, age of smoking initiation, week 0 urinary total NNAL (transformed, pg/mg creatinine), week 0 urinary cotinine (transformed, ng/mg creatinine), week 0 exhaled CO, week 0 cigarettes smoked per day, week 0 body mass index, week 0 diastolic blood pressure, week 0 center for Epidemiologic Studies Depression Scale score, week 0 Clinical Chronic Obstructive Pulmonary Disease score, week 0 partial INTERHEART non-laboratory score, week 0 Kessler K6 score, week 0 Penn State Cigarette Dependence Index score, week 0 Perceived Stress Scale score, week 0 forced expiratory volume in one second, week 0 forced vital capacity |
| **Complete metabolic panel, complete blood count, Lipoproteins** | Study site, gender, age, race/ethnicity, education, total household income, age of smoking initiation, week 0 environmental smoke score, week 0 urinary total NNAL (transformed, pg/mg creatinine), week 0 urinary cotinine (transformed, ng/mg creatinine), week 0 exhaled CO, week 0 cigarettes smoked per day, week 0 body mass index, week 0 height, week 0 weight, week 0 hip circumference, week 0 waist circumference, week 0 diastolic blood pressure, week 0 Center for Epidemiologic Studies Depression Scale score, week 0 Clinical Chronic Obstructive Pulmonary Disease score, week 0 partial INTERHEART non-laboratory score, week 0 Kessler K6 score, week 0 Penn State Cigarette Dependence Index score, week 0 Perceived Stress Scale score, week 0 forced expiratory volume in one second, week 0 forced vital capacity |
| **Other Physiologic Measures (Weight, BMI, pulse, Systolic BP, Diastolic BP)** | Study site, gender, age, race/ethnicity, education, age of smoking initiation, week 0 Environmental Smoke score, week 0 urinary total NNAL (transformed, pg/mg creatinine), week 0 urinary cotinine (transformed, ng/mg creatinine), week 0 exhaled CO, week 0 cigarettes smoked per day, week 0 body mass index, week 0 height, week 0 weight, week 0 hip circumference, week 0 waist circumference, week 0 diastolic blood pressure, week 0 Clinical Chronic Obstructive Pulmonary Disease score, week 0 partial INTERHEART non-laboratory score, week 0 Kessler K6 score, week 0 Penn State Cigarette Dependence Index score, week 0 Perceived Stress Scale score, week 0 forced expiratory volume in one second, week 0 forced vital capacity |

**Supplemental Table 3: Sample size for each of the outcome measurements at 6 months by overall and conditions**

| Characteristics | | EC Conditions | | | |
|---|---|---|---|---|---|
| | CS | 0 mg/mL | 8 mg/mL | 36 mg/mL | Overall |
| | n | n | n | n | N |
| **Pulmonary Function Tests** | | | | | |
| FEV1, L | 91 | 69 | 74 | 80 | 314 |
| FEV1% | 91 | 69 | 74 | 80 | 314 |
| FVC, L | 91 | 69 | 74 | 80 | 314 |
| FVC% | 91 | 69 | 74 | 80 | 314 |
| FEV1/FVC | 91 | 69 | 74 | 80 | 314 |
| FEV1/FVC% | 91 | 69 | 74 | 80 | 314 |
| PEF | 91 | 69 | 74 | 80 | 314 |
| PEF% | 91 | 69 | 74 | 80 | 314 |
| FEF 25 | 91 | 69 | 74 | 80 | 314 |
| FEF 75 | 91 | 69 | 74 | 80 | 314 |
| FEF 25-75 | 91 | 69 | 74 | 80 | 314 |
| FEF 25-75% | 91 | 69 | 74 | 80 | 314 |
| FET | 91 | 67 | 73 | 80 | 311 |
| COPD Symptoms | 91 | 69 | 73 | 80 | 313 |
| COPD Functional State | 91 | 68 | 73 | 80 | 312 |
| COPD Mental State | 89 | 69 | 73 | 80 | 311 |
| COPD Total Score | 89 | 68 | 73 | 80 | 310 |
| **Complete Metabolic Panel** | | | | | |
| Sodium, mmol/L | 88 | 67 | 73 | 79 | 307 |
| Potassium, mmol/L | 88 | 67 | 73 | 79 | 307 |
| Chloride, mmol/L | 88 | 67 | 73 | 79 | 307 |
| Carbon dioxide, mmol/L | 88 | 67 | 73 | 79 | 307 |
| Anion gap, mmol/L | 88 | 67 | 73 | 79 | 307 |
| Glucose, mg/dL | 88 | 67 | 73 | 79 | 307 |
| BUN, mg/dL | 88 | 67 | 73 | 79 | 307 |
| AST, units/L | 88 | 67 | 73 | 79 | 307 |
| ALT, units/L | 88 | 67 | 73 | 79 | 307 |
| ALP, units/L | 88 | 67 | 73 | 79 | 307 |
| Bilirubin, mg/dL | 88 | 67 | 73 | 79 | 307 |
| Protein, mg/dL | 88 | 67 | 73 | 79 | 307 |
| Albumin, mg/dL | 88 | 67 | 73 | 79 | 307 |
| Calcium, mg/dL | 88 | 67 | 73 | 79 | 307 |
| CRP, mg/dL | 88 | 67 | 74 | 79 | 308 |
| Creatinine, mg/dL | 90 | 69 | 73 | 79 | 311 |
| **Complete Blood Count** | | | | | |
| WBC, $10^9$/L | 87 | 67 | 73 | 80 | 307 |
| RBC, $10^{12}$/L | 87 | 67 | 73 | 80 | 307 |
| Hemoglobin, g/dL | 87 | 67 | 73 | 80 | 307 |
| Hematocrit, % | 87 | 67 | 73 | 80 | 307 |
| MCV, fL | 87 | 67 | 73 | 80 | 307 |
| MCH, pg | 87 | 67 | 73 | 80 | 307 |
| MCHC, g/dL | 87 | 67 | 73 | 80 | 307 |
| RDW, % | 87 | 67 | 73 | 80 | 307 |
| Platelets, $10^9$/L | 87 | 67 | 73 | 80 | 307 |
| MPV, fL | 87 | 67 | 73 | 80 | 307 |

| | | | | | |
|---|---|---|---|---|---|
| **Lipoproteins** | | | | | |
| Cholesterol, mg/dL | 88 | 67 | 73 | 79 | 307 |
| HDL, mg/dL | 88 | 67 | 73 | 79 | 307 |
| Non-HDL Cholesterol, mg/dL | 88 | 67 | 73 | 79 | 307 |
| Cholesterol/HDL Ratio | 88 | 66 | 73 | 79 | 306 |
| LDL Calculated, mg/dL | 86 | 66 | 70 | 77 | 299 |
| Triglycerides, mg/dL | 88 | 67 | 73 | 79 | 307 |
| **Other Physiologic Measures** | | | | | |
| Weight, pound | 91 | 69 | 74 | 80 | 314 |
| BMI, $kg/m^2$ | 91 | 69 | 74 | 80 | 314 |
| Heart rate, bpm | 91 | 68 | 74 | 80 | 313 |
| Systolic blood pressure, mmHg | 91 | 68 | 74 | 80 | 313 |
| Diastolic blood pressure, mmHg | 91 | 68 | 74 | 80 | 313 |
| Waist-to-hip ratio | 91 | 69 | 74 | 80 | 314 |
| INTERHEART risk score | 91 | 69 | 73 | 80 | 313 |

Note: CS, cigarette substitute; FEV1, forced expiratory volume in 1 second; FVC, forced vital capacity; PEF, peak expiratory flow; FEF, forced expiratory flow; FET, forced expiratory time; BUN, blood urea nitrogen; AST, aspartate aminotransferase; ALT, alkaline transferase; ALP, alkaline phosphatase; CRP, C-reactive Protein; WBC, white blood cells; RBC, red blood cells; MCV, mean corpuscular volume; MCH, mean corpuscular hemoglobin; MCHC, mean corpuscular hemoglobin concentration; RDW, red cell distribution width; MPV, mean platelet volume; HDL, high-density lipoprotein; LDL, low-density lipoprotein; COPD, chronic obstructive pulmonary disorder; BMI, body mass index.

**Supplemental Figure 1: Consort Flow Diagram**

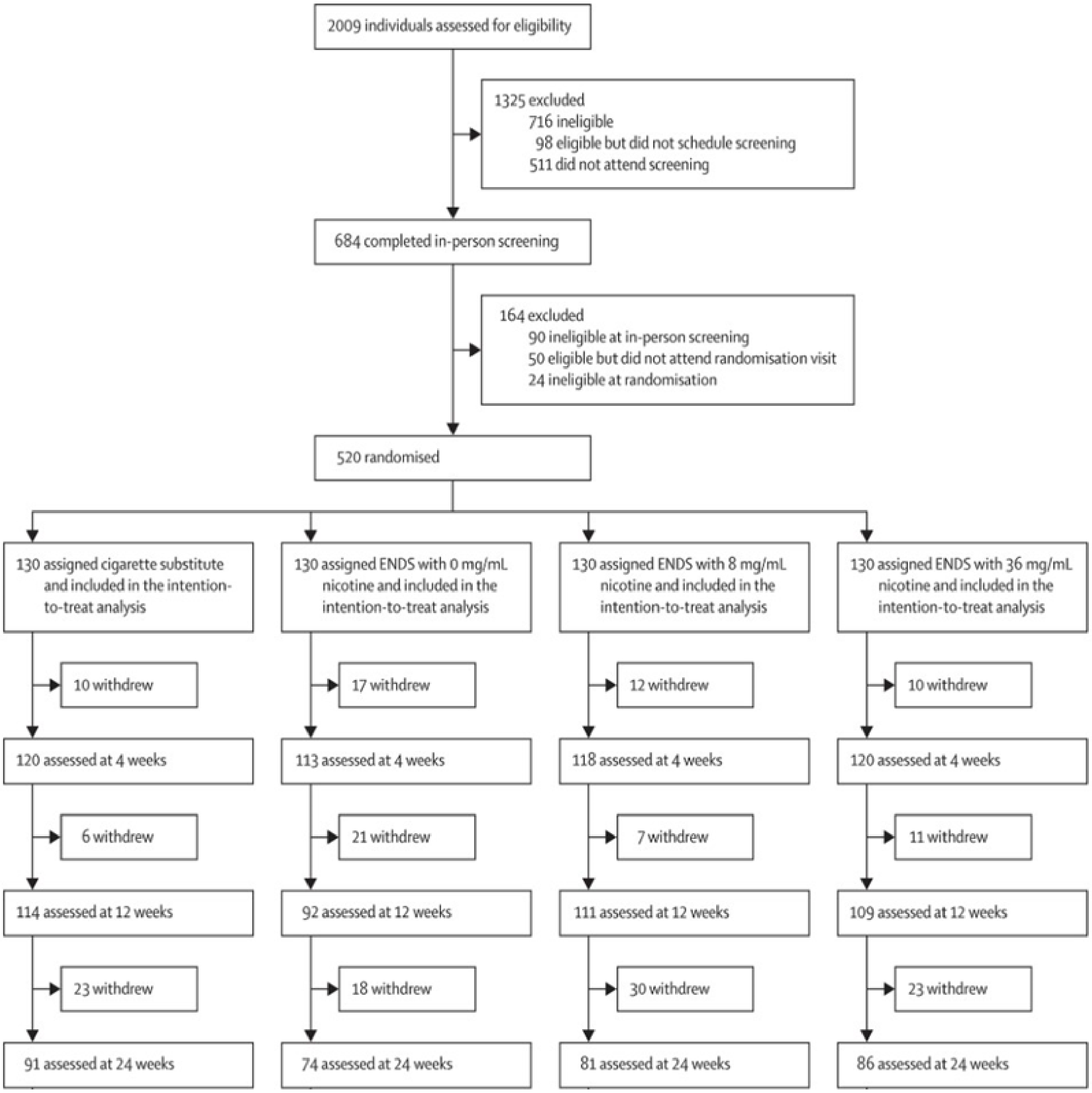

#### Supplemental Figure 2: Unadjusted and adjusted analyses of FEV1

**A**

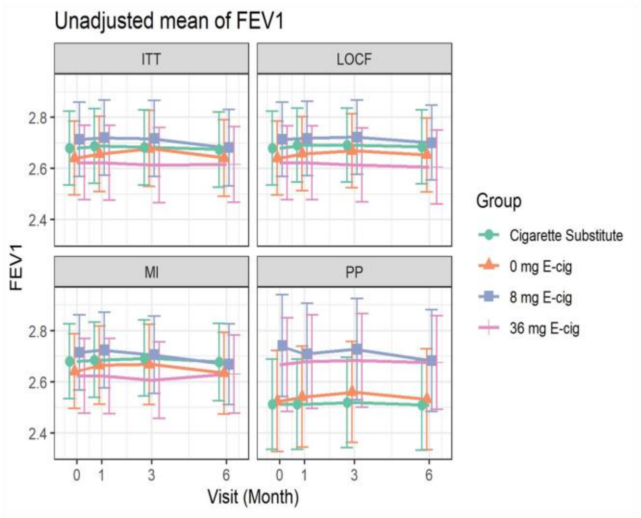

**B**

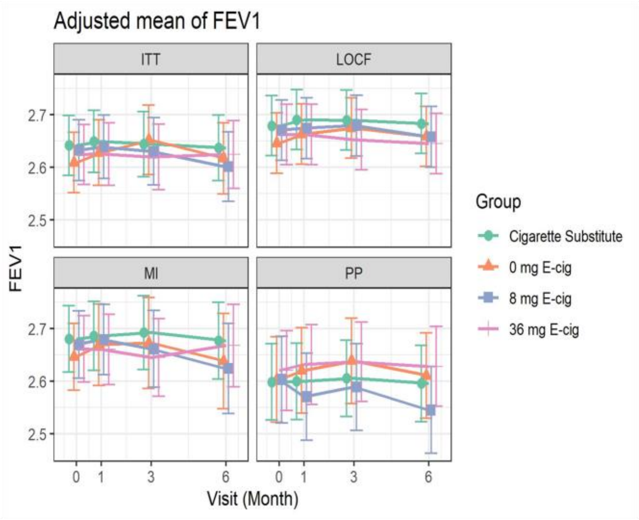

**C.** Unadjusted Mean

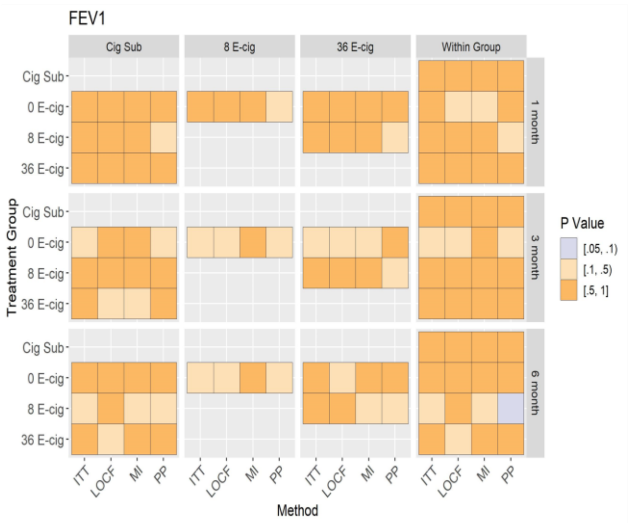

**D.** Adjusted Mean

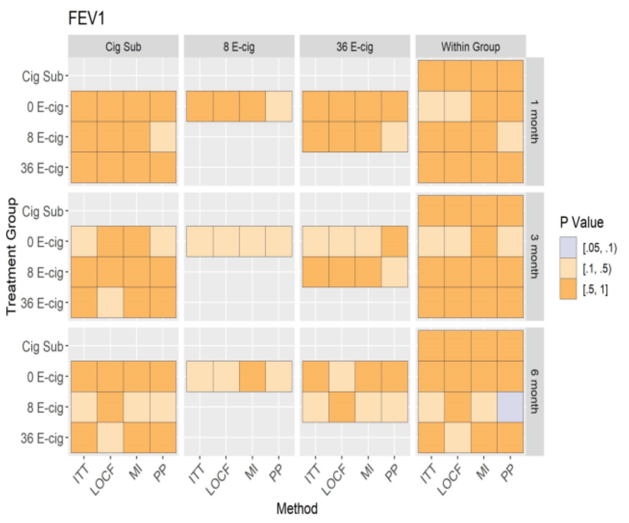

Figure A, B: Mean of FEV1 (95% CI) at baseline, 1, 3, and 6 months (unadjusted and adjusted models) across randomized groups. Figure C, D: Between and within group differences in FEV1 at baseline, 1, 3 and 6 months (unadjusted and adjusted models) among all randomized participants.

**Supplemental Figure 3: Unadjusted and adjusted analyses of FEV1%**

**A**

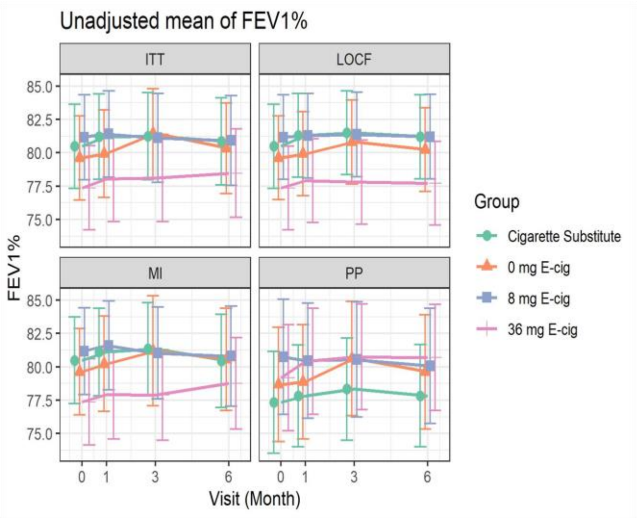

**B**

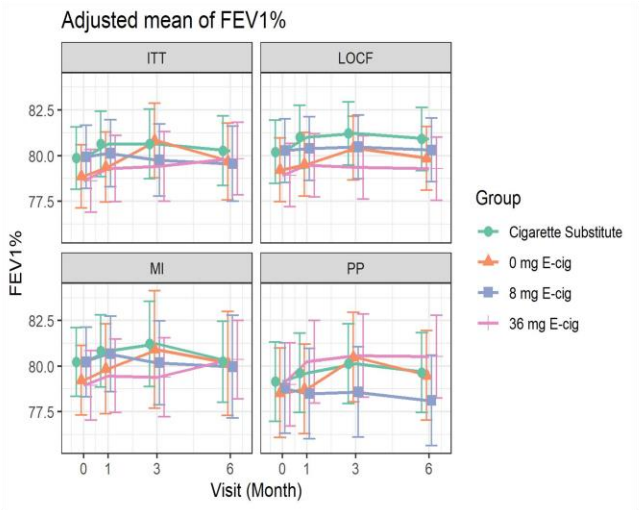

**C.** Unadjusted Mean

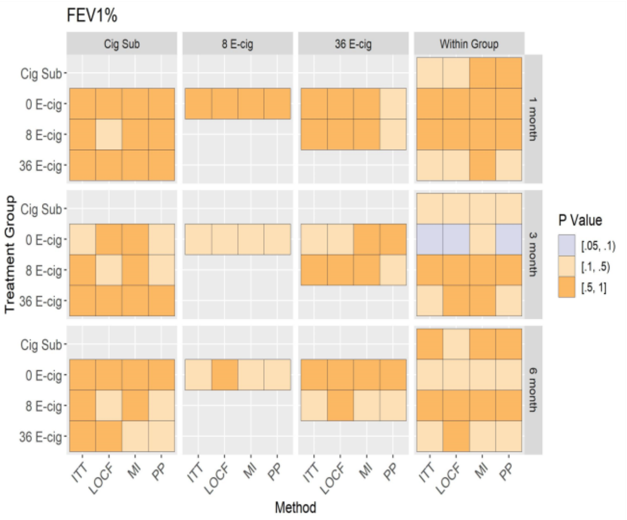

**D.** Adjusted Mean

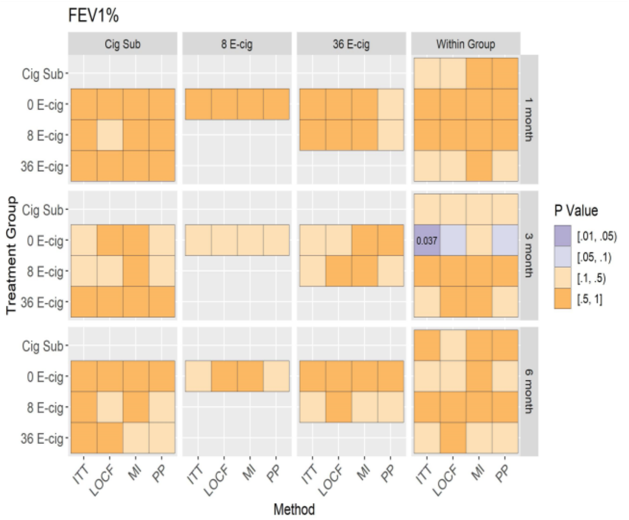

Figure A, B: Mean of FEV1% (95% CI) at baseline, 1, 3, and 6 months (unadjusted and adjusted models) across randomized groups. Figure C, D: Between and within group differences in FEV1% at baseline, 1, 3 and 6 months (unadjusted and adjusted models) among all randomized participants.

#### Supplemental Figure 4: Unadjusted and adjusted analyses of FVC

**A**

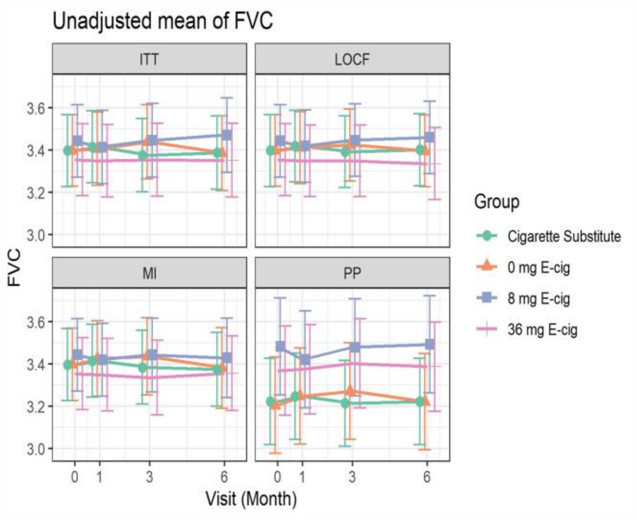

**B**

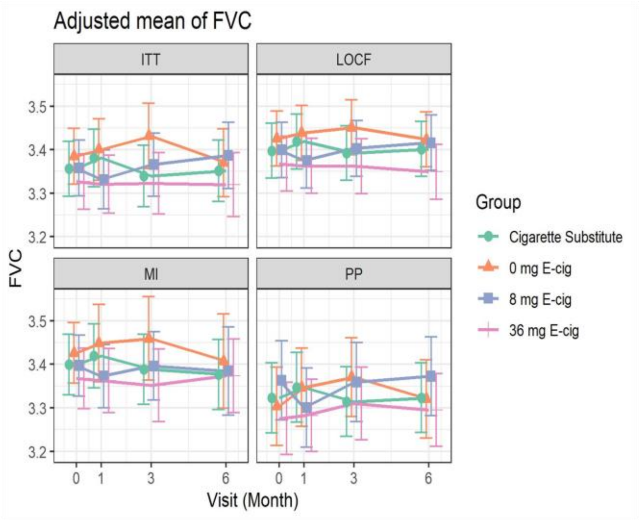

**C.** Unadjusted Mean

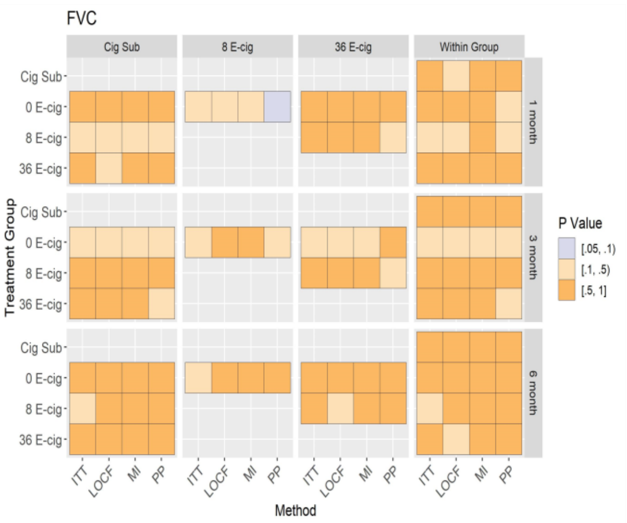

**D.** Adjusted Mean

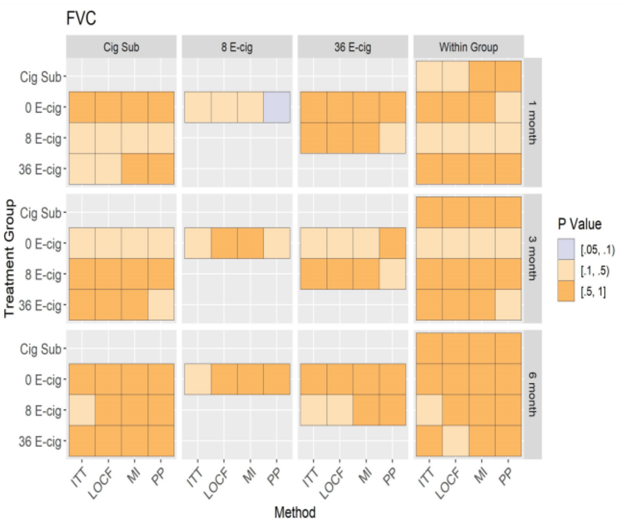

Figure A, B: Mean of FVC (95% CI) at baseline, 1, 3, and 6 months (unadjusted and adjusted models) across randomized groups. Figure C, D: Between and within group differences in FVC at baseline, 1, 3 and 6 months (unadjusted and adjusted models) among all randomized participants.

**Supplemental Figure 5: Unadjusted and adjusted analyses of FVC%**

**A**

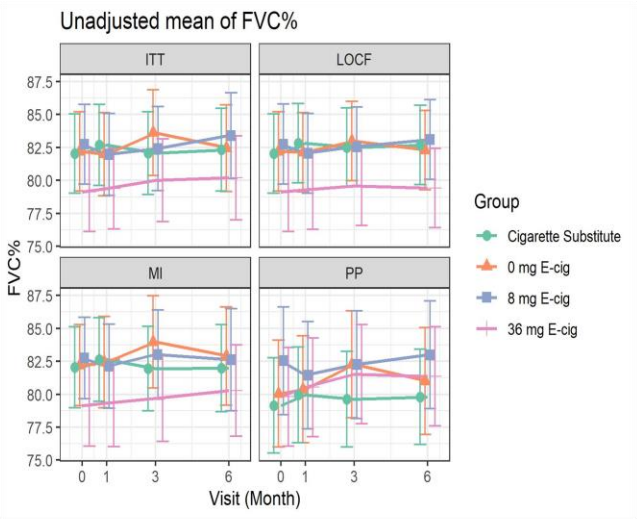

**B**

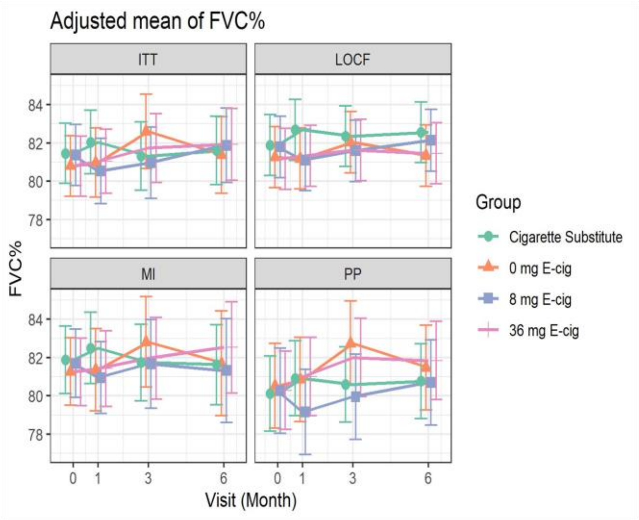

**C.** Unadjusted Mean

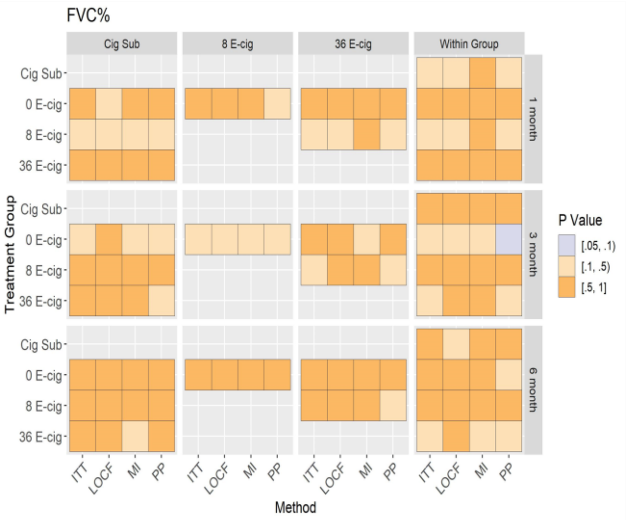

**D.** Adjusted Mean

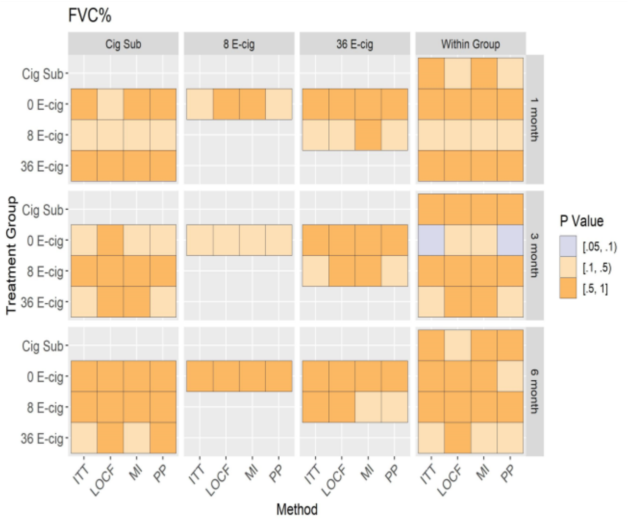

Figure A, B: Mean of FVC% (95% CI) at baseline, 1, 3, and 6 months (unadjusted and adjusted models) across randomized groups. Figure C, D: Between and within group differences in FVC% at baseline, 1, 3 and 6 months (unadjusted and adjusted models) among all randomized participants.

#### Supplemental Figure 6: Unadjusted and adjusted analyses of PEF

**A**

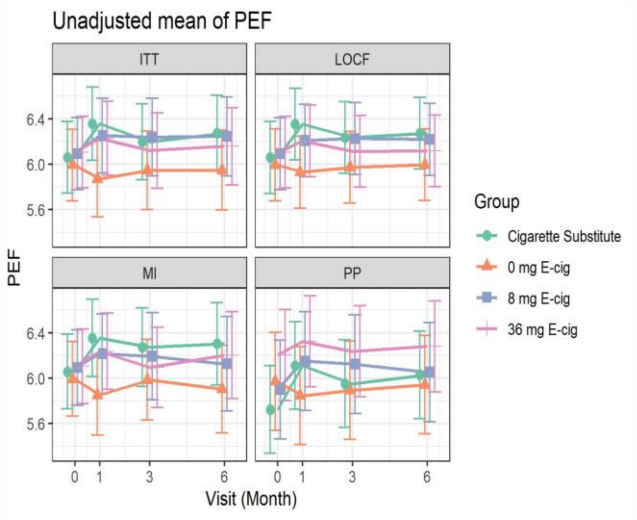

**B**

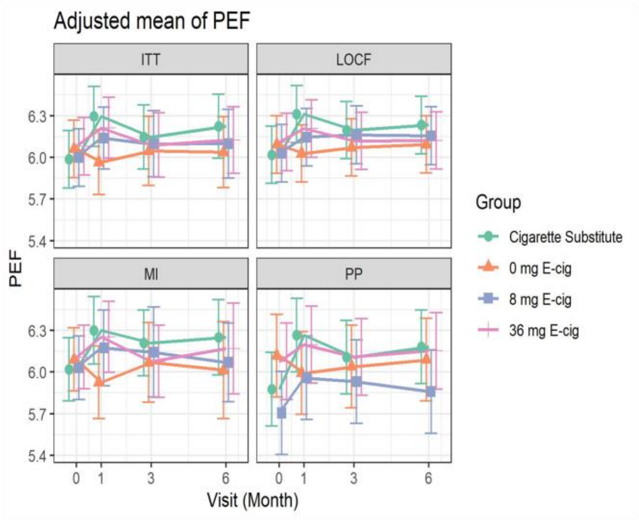

**C.** Unadjusted Mean

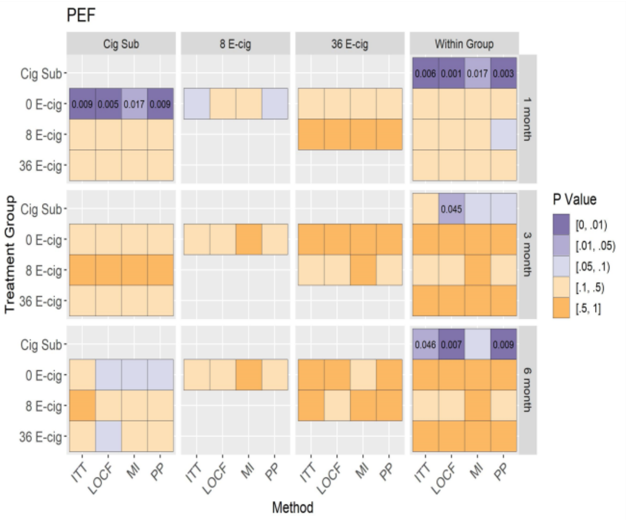

**D.** Adjusted Mean

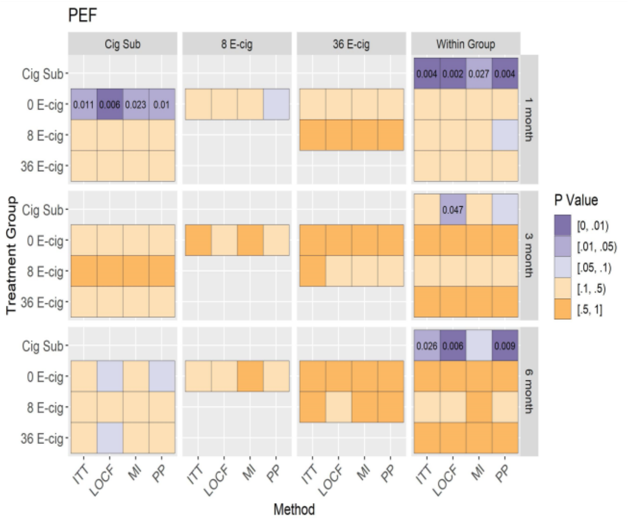

Figure A, B: Mean of PEF (95% CI) at baseline, 1, 3, and 6 months (unadjusted and adjusted models) across randomized groups. Figure C, D: Between and within group differences in PEF at baseline, 1, 3 and 6 months (unadjusted and adjusted models) among all randomized participants.

#### Supplemental Figure 7: Unadjusted and adjusted analyses of PEF%

**A**

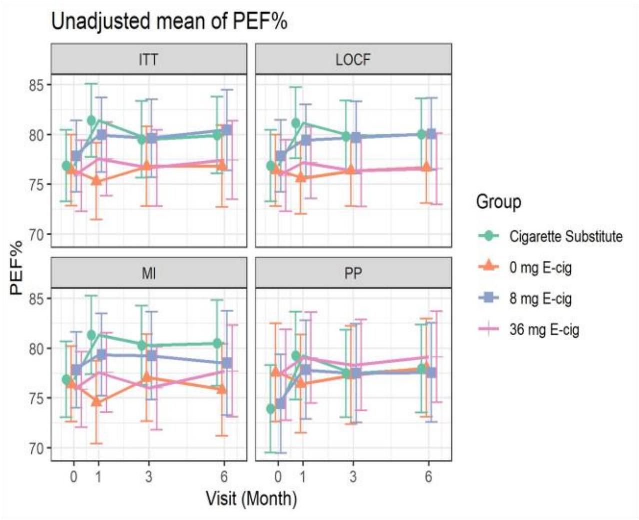

**B**

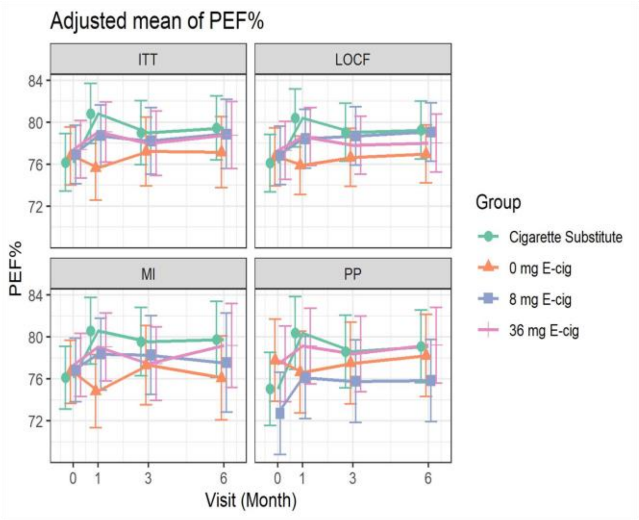

**C.** Unadjusted Mean

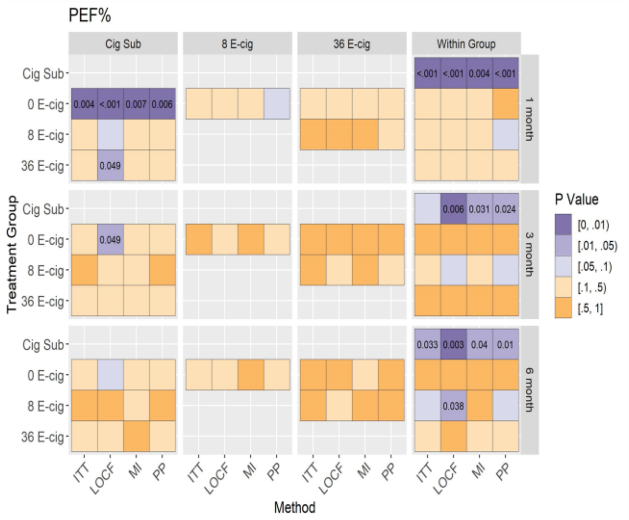

**D.** Adjusted Mean

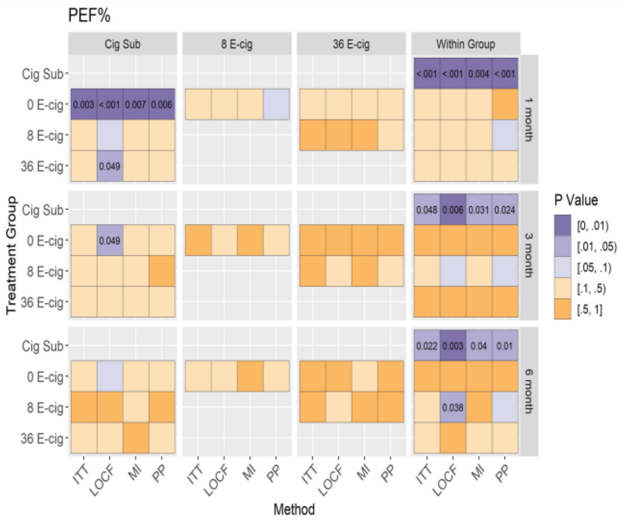

Figure A, B: Mean of PEF% (95% CI) at baseline, 1, 3, and 6 months (unadjusted and adjusted models) across randomized groups. Figure C, D: Between and within group differences in PEF% at baseline, 1, 3 and 6 months (unadjusted and adjusted models) among all randomized participants.

### Supplemental Figure 8: Unadjusted and adjusted analyses of FEV1/FVC

**A**

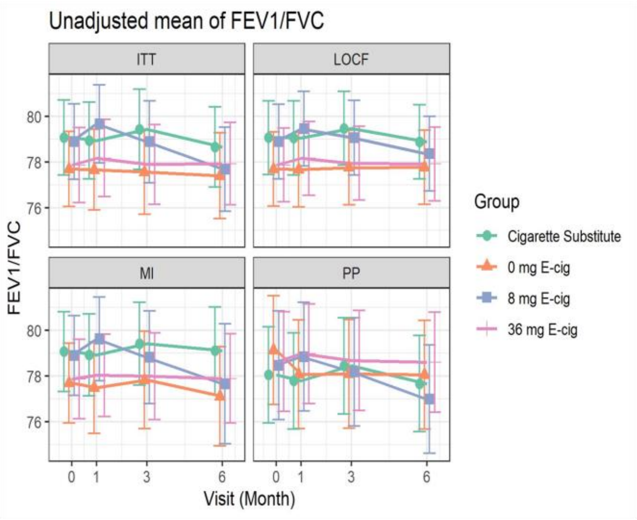

**B**

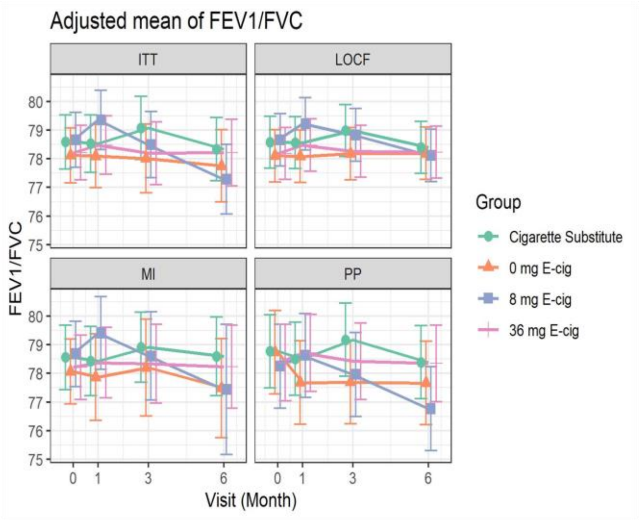

**C.** Unadjusted Mean

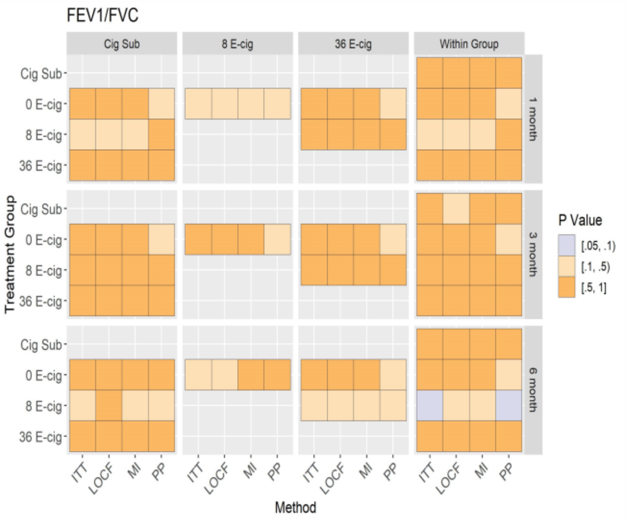

**D.** Adjusted Mean

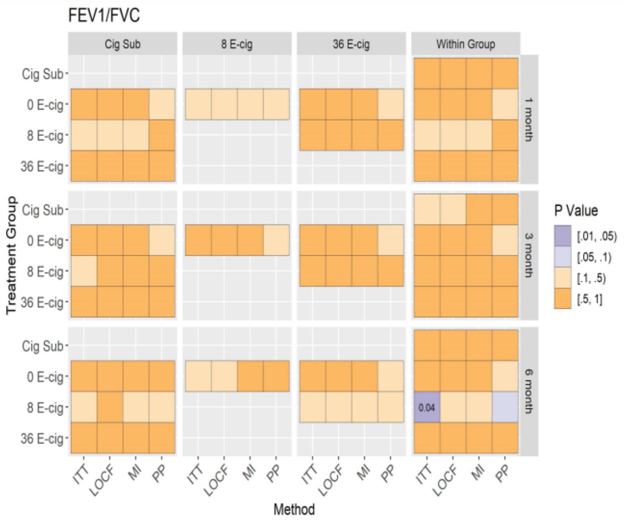

Figure A, B: Mean of FEV1/FVC (95% CI) at baseline, 1, 3, and 6 months (unadjusted and adjusted models) across randomized groups. Figure C, D: Between and within group differences in FEV1/FVC at baseline, 1, 3 and 6 months (unadjusted and adjusted models) among all randomized participants.

#### Supplemental Figure 9: Unadjusted and adjusted analyses of FEV1/FVC%

**A**

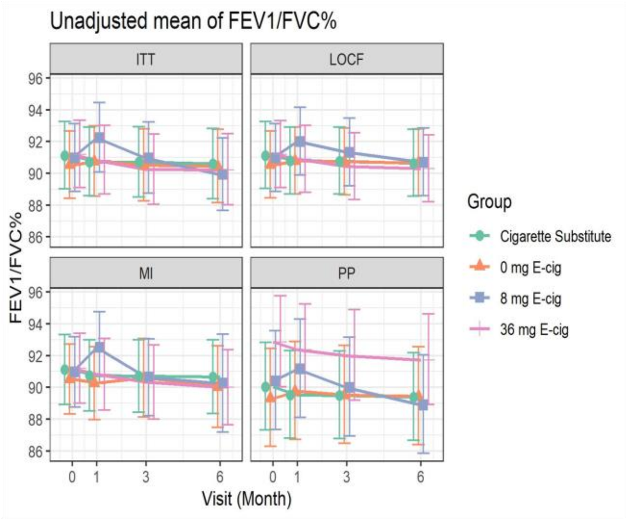

**B**

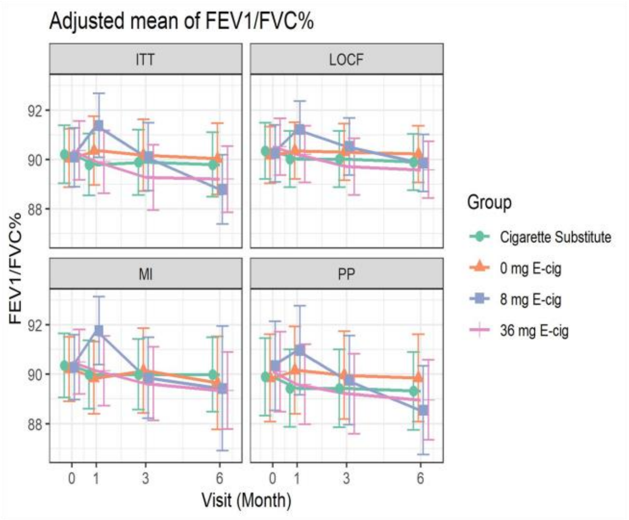

**C.** Unadjusted Mean

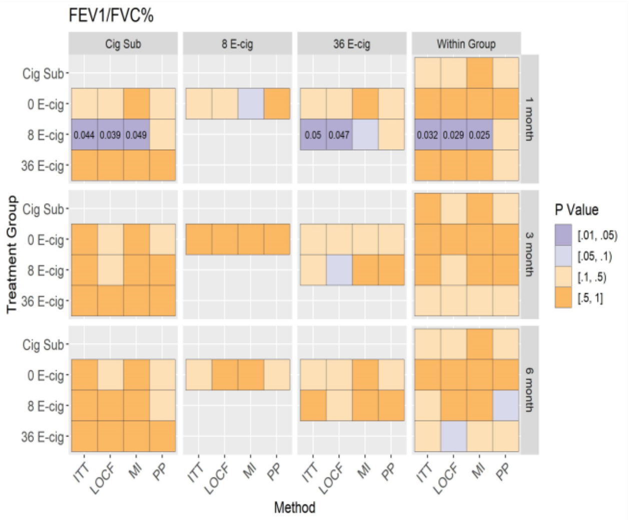

**D.** Adjusted Mean

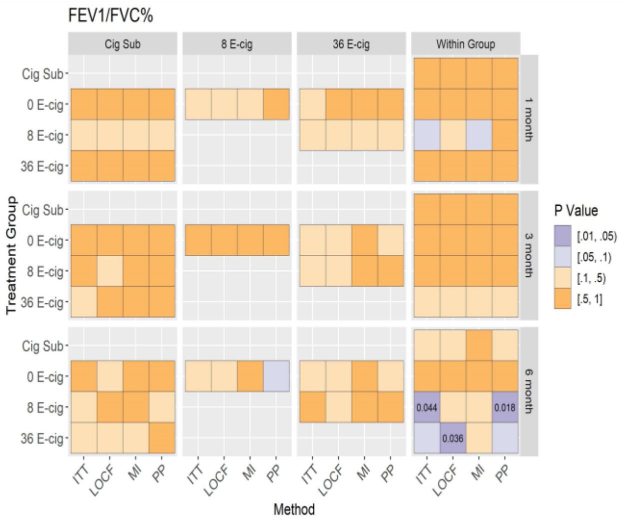

Figure A, B: Mean of FEV1/FVC% (95% CI) at baseline, 1, 3, and 6 months (unadjusted and adjusted models) across randomized groups. Figure C, D: Between group differences in FEV1/FVC% at baseline, 1, 3 and 6 months (unadjusted and adjusted models), and within group differences between baseline and follow-up visits among all randomized participants.

**Supplemental Figure 10: Unadjusted and adjusted analyses of FEF 25**

**A**

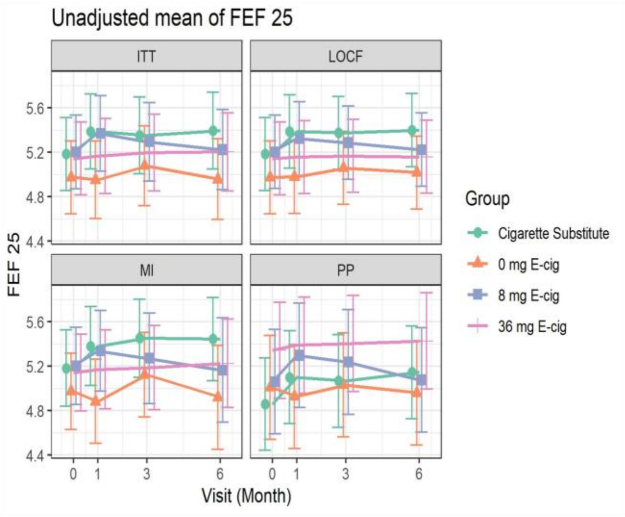

**B**

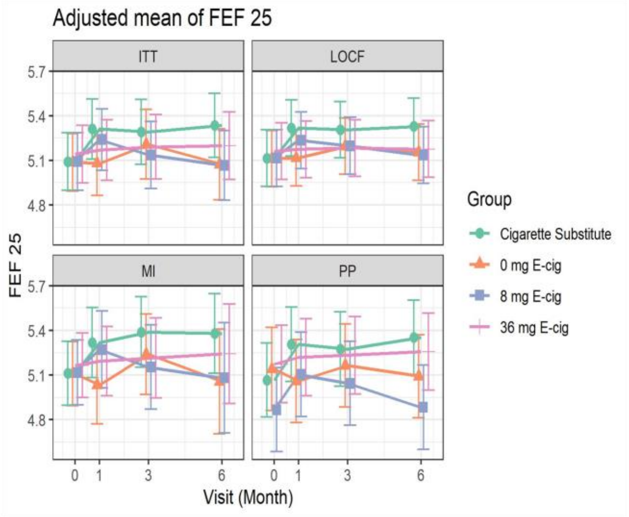

**C.** Unadjusted Mean

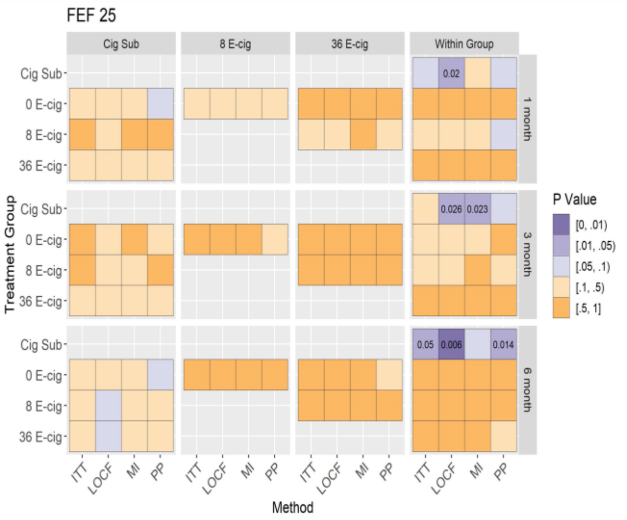

**D.** Adjusted Mean

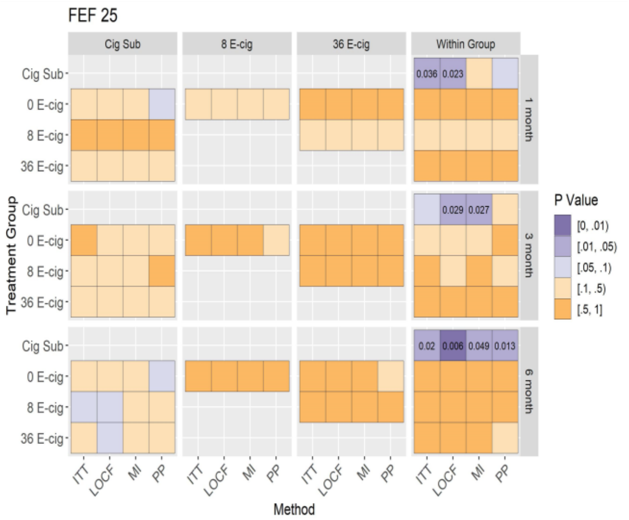

Figure A, B: Mean of FEF 25 (95% CI) at baseline, 1, 3, and 6 months (unadjusted and adjusted models) across randomized groups. Figure C, D: Between group differences in FEF 25 at baseline, 1, 3 and 6 months (unadjusted and adjusted models), and within group differences between baseline and follow-up visits among all randomized participants.

**Supplemental Figure 11: Unadjusted and adjusted analyses of FEF 75**

**A**

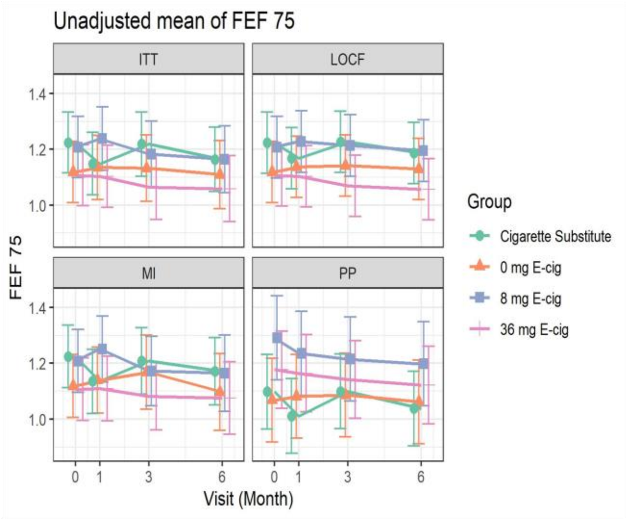

**B**

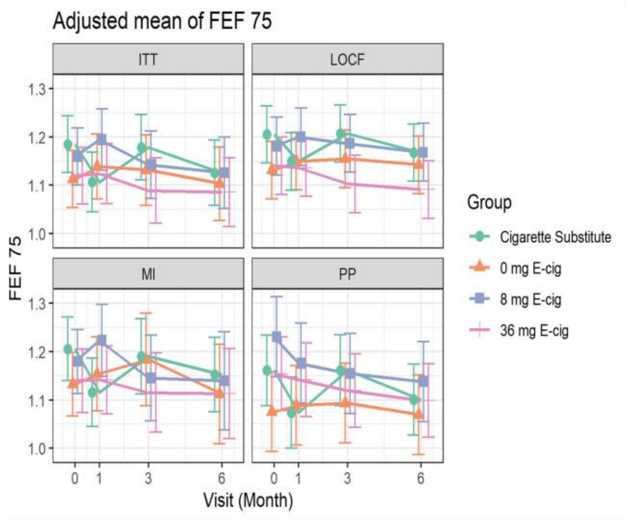

**C.** Unadjusted Mean

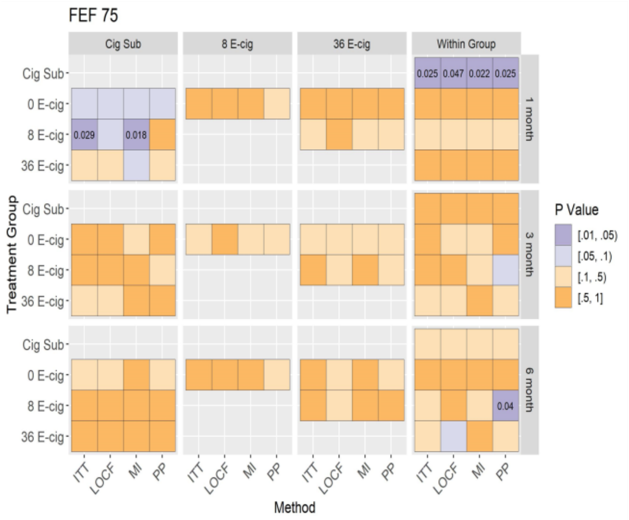

**D.** Adjusted Mean

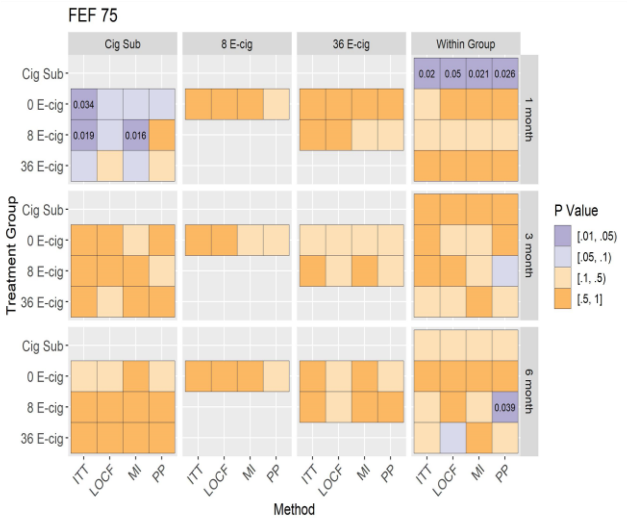

Figure A, B: Mean of FEF 75 (95% CI) at baseline, 1, 3, and 6 months (unadjusted and adjusted models) across randomized groups. Figure C, D: Between group differences in FEF 75 at baseline, 1, 3 and 6 months (unadjusted and adjusted models), and within group differences between baseline and follow-up visits among all randomized participants.

**Supplemental Figure 12: Unadjusted and adjusted analyses of FEF 25-75**

**A**

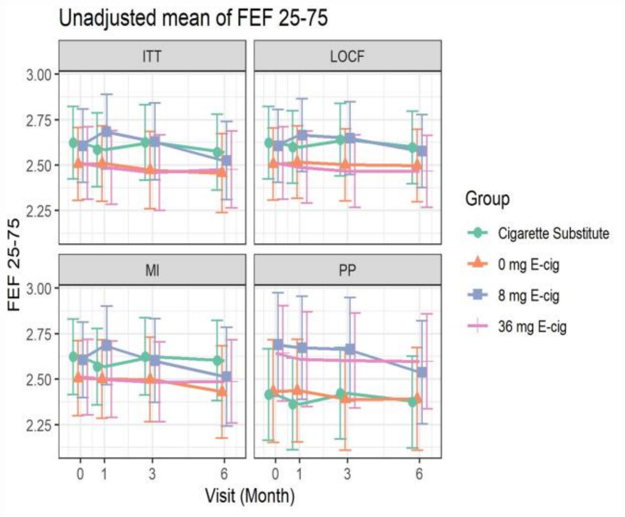

**B**

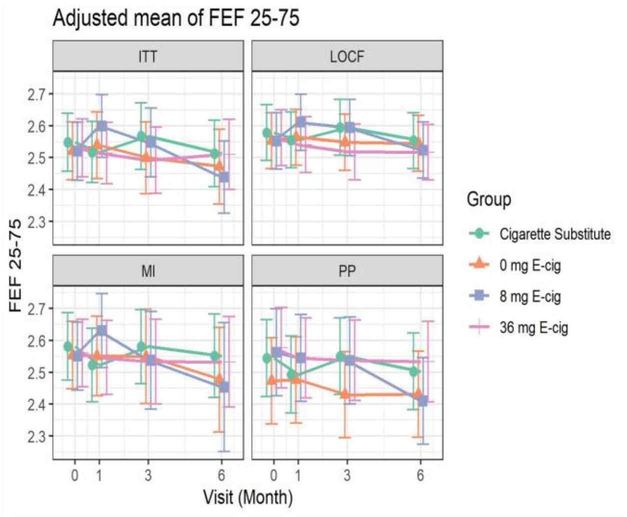

**C.** Unadjusted Mean

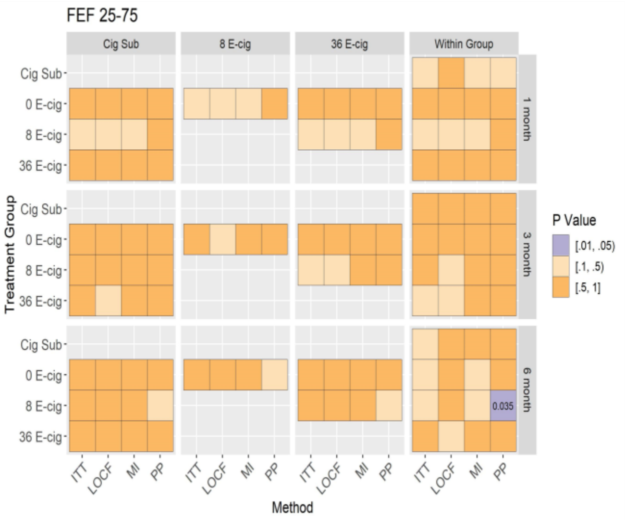

**D.** Adjusted Mean

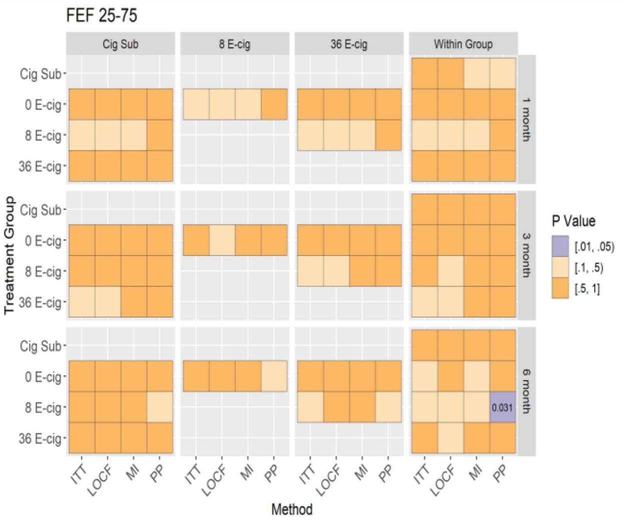

Figure A, B: Mean of FEF 25-75 (95% CI) at baseline, 1, 3, and 6 months (unadjusted and adjusted models) across randomized groups. Figure C, D: Between group differences in FEF 25-75 at baseline, 1, 3 and 6 months (unadjusted and adjusted models), and within group differences between baseline and follow-up visits among all randomized participants.

**Supplemental Figure 13: Unadjusted and adjusted analyses of FEF 25-75%**

**A**

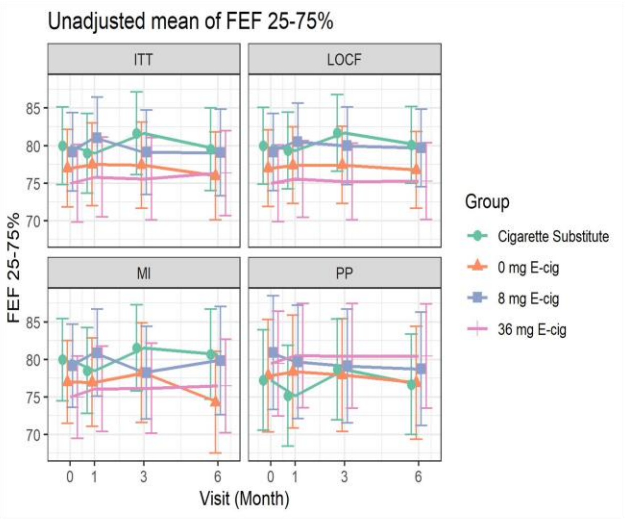

**B**

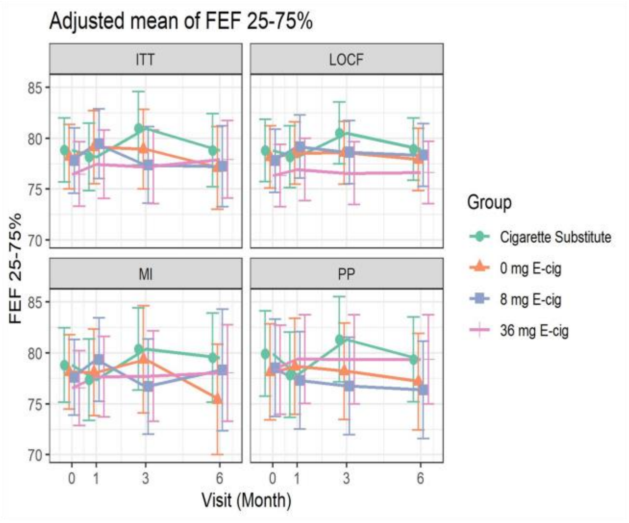

**C.** Unadjusted Mean

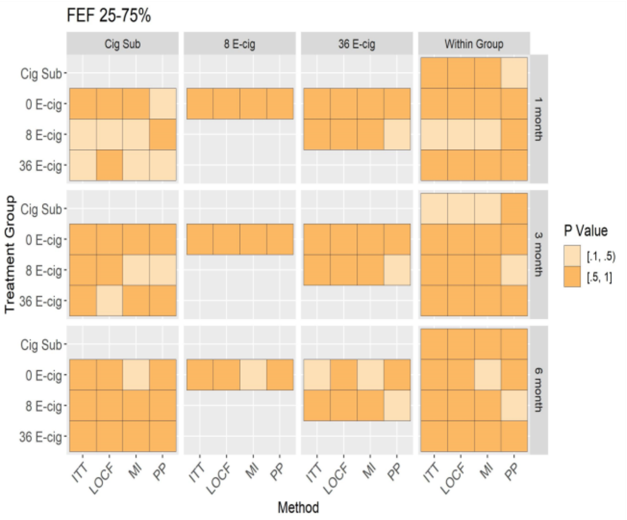

**D.** Adjusted Mean

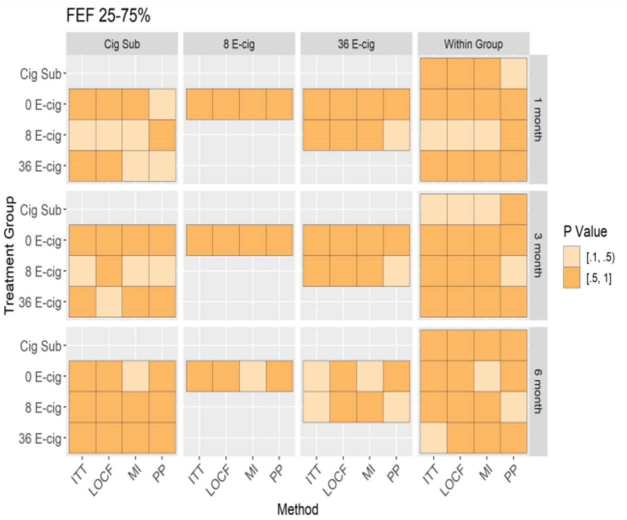

Figure A, B: Mean of FEF 25-75% (95% CI) at baseline, 1, 3, and 6 months (unadjusted and adjusted models) across randomized groups. Figure C, D: Between group differences in FEF 25-75% at baseline, 1, 3 and 6 months (unadjusted and adjusted models), and within group differences between baseline and follow-up visits among all randomized participants.

**Supplemental Figure 14: Unadjusted and adjusted analyses of Forced expiratory time in seconds**

**A**

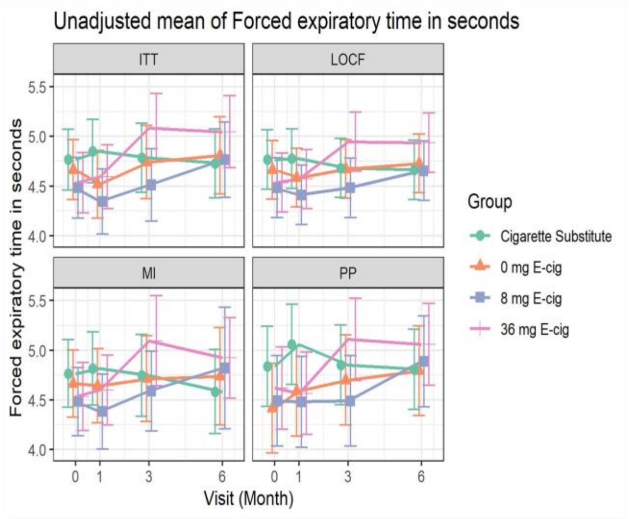

**B**

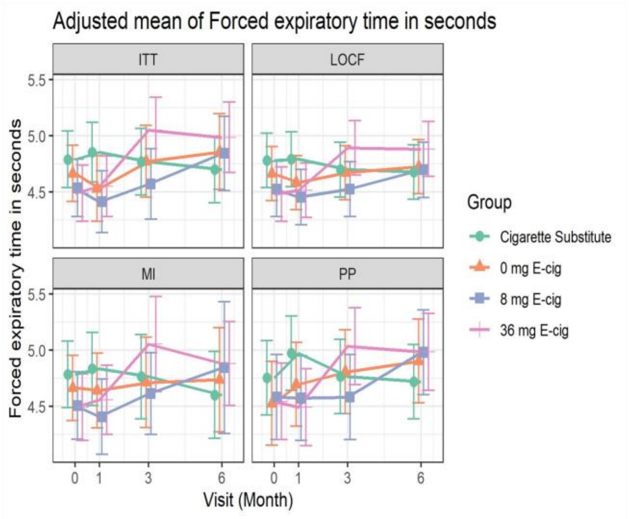

**C.** Unadjusted Mean

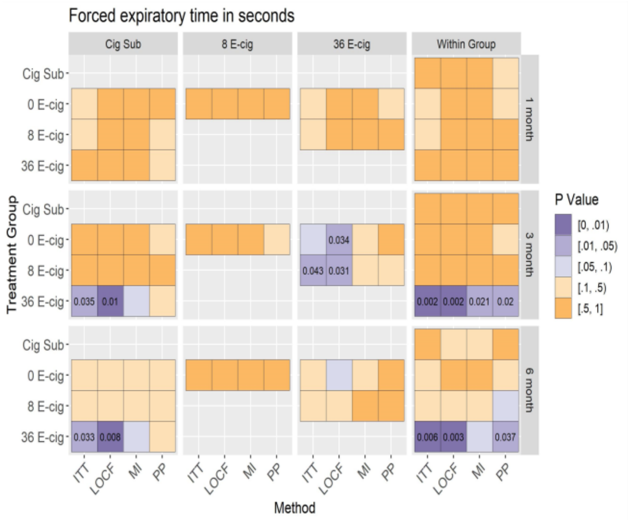

**D.** Adjusted Mean

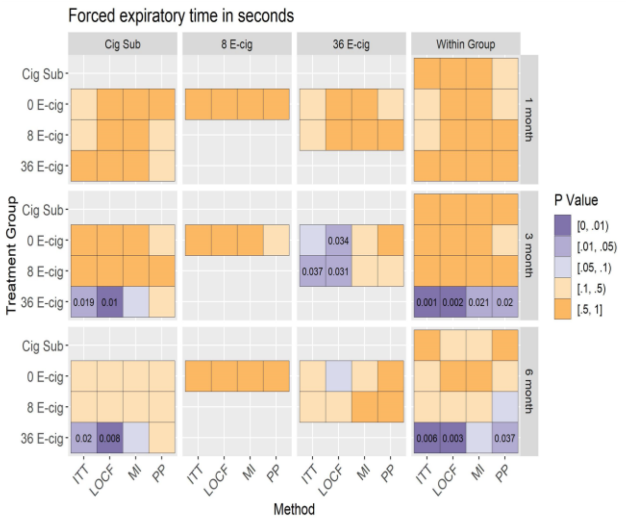

Figure A, B: Mean of Forced expiratory time in seconds (95% CI) at baseline, 1, 3, and 6 months (unadjusted and adjusted models) across randomized groups. Figure C, D: Between group differences in Forced expiratory time in seconds at baseline, 1, 3 and 6 months (unadjusted and adjusted models), and within group differences between baseline and follow-up visits among all randomized participants.

**Supplemental Figure 15: Unadjusted and adjusted analyses of COPD symptoms**

**A**

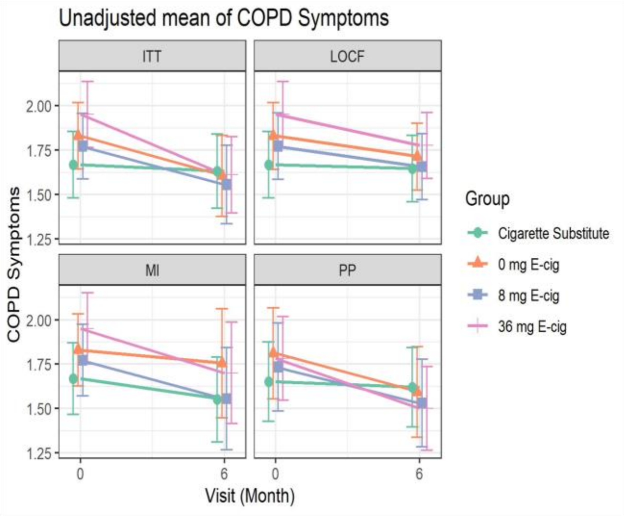

**B**

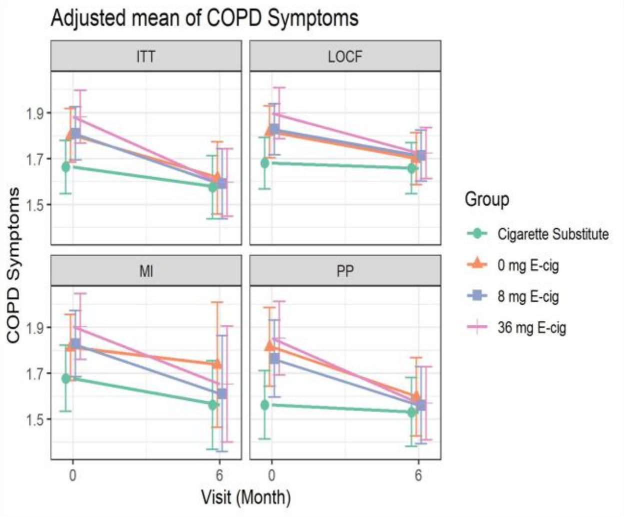

**C.** Unadjusted Mean

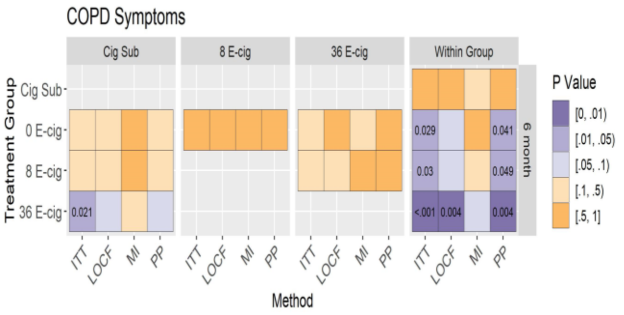

**D.** Adjusted Mean

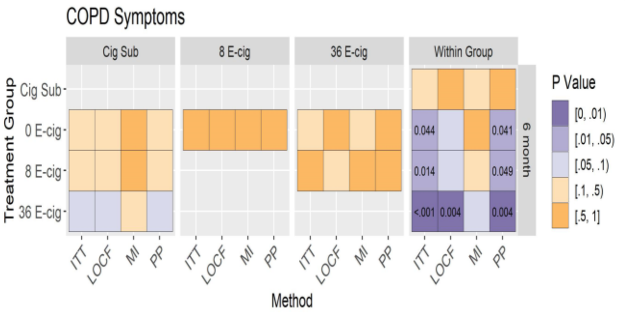

Figure A, B: Mean of COPD symptoms (95% CI) at baseline and 6 months (unadjusted and adjusted models) across randomized groups. Figure C, D: Between and within group differences in COPD symptoms at 6 months (unadjusted and adjusted models) among all randomized participants.

**Supplemental Figure 16: Unadjusted and adjusted analyses of COPD functional state**

**A**

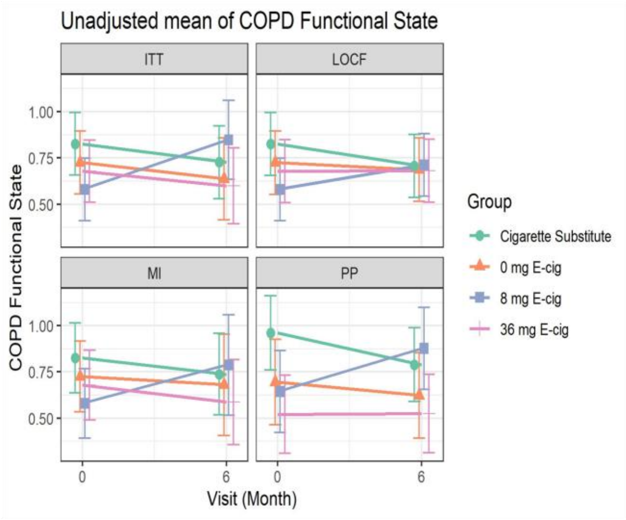

**B**

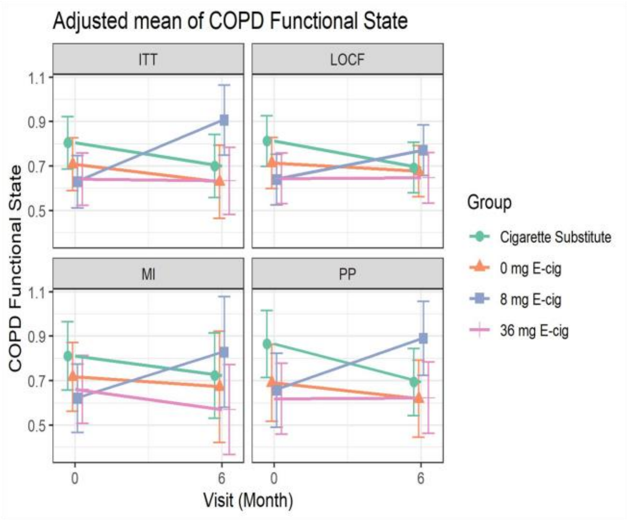

**C.** Unadjusted Mean

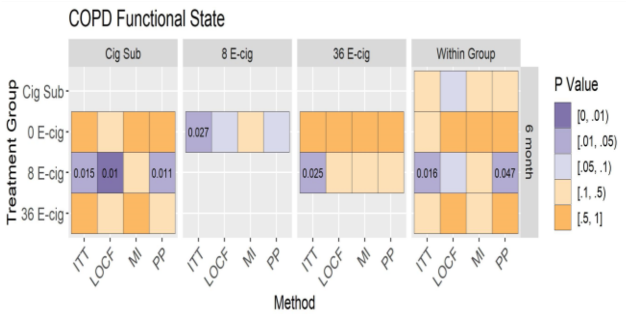

**D.** Adjusted Mean

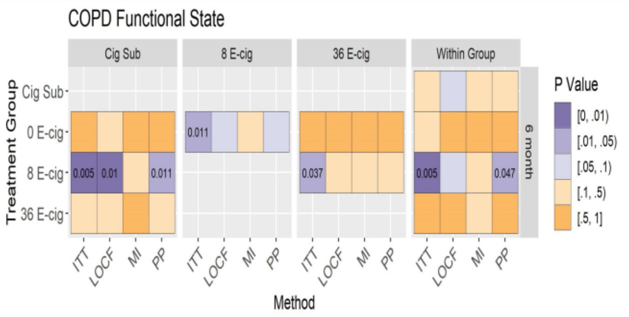

Figure A, B: Mean of COPD functional state (95% CI) at baseline and 6 months (unadjusted and adjusted models) across randomized groups. Figure C, D: Between and within group differences in COPD functional state at 6 months (unadjusted and adjusted models) among all randomized participants.

### Supplemental Figure 17: Unadjusted and adjusted analyses of COPD mental state

**A**

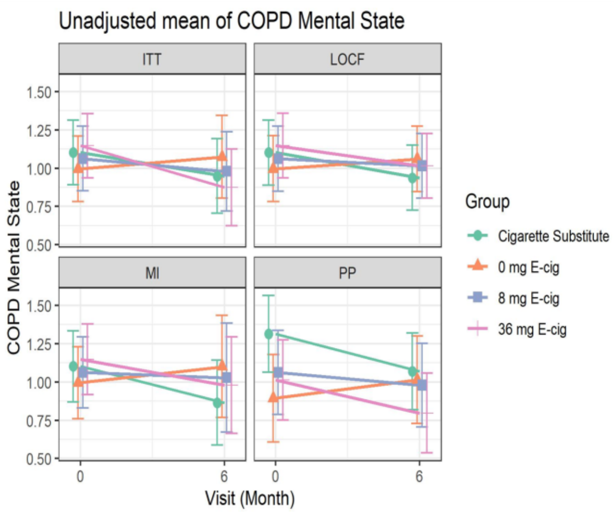

**B**

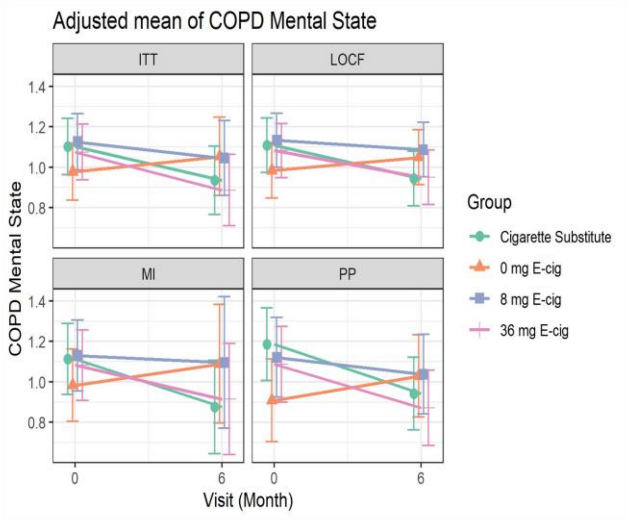

**C.** Unadjusted Mean

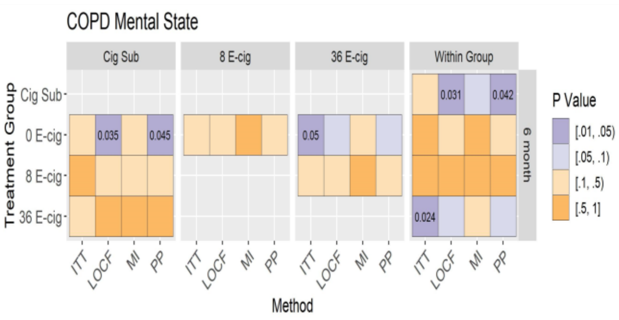

**D.** Adjusted Mean

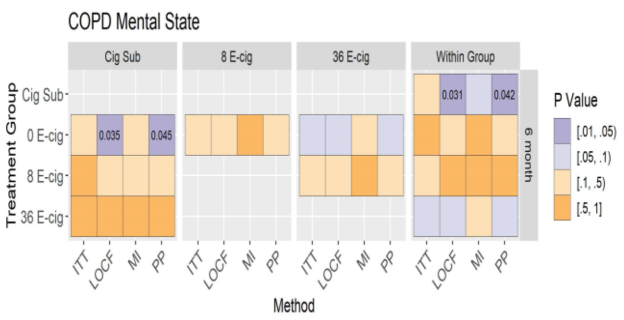

Figure A, B: Mean of COPD mental state (95% CI) at baseline and 6 months (unadjusted and adjusted models) across randomized groups. Figure C, D: Between and within group differences in COPD mental state at 6 months (unadjusted and adjusted models) among all randomized participants.

#### Supplemental Figure 18: Unadjusted and adjusted analyses of total COPD score

**A**

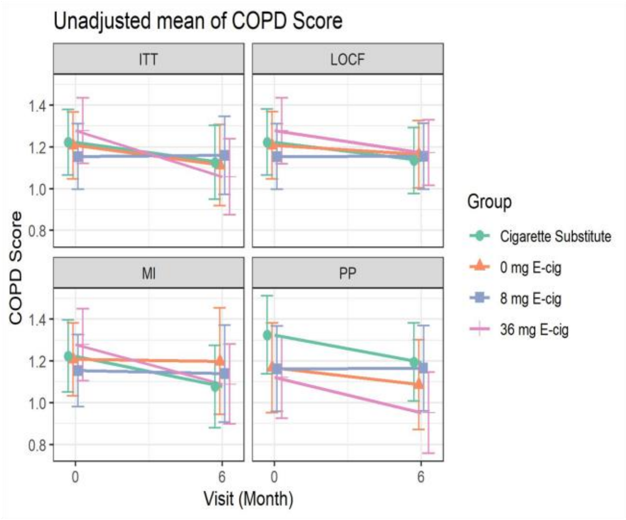

**B**

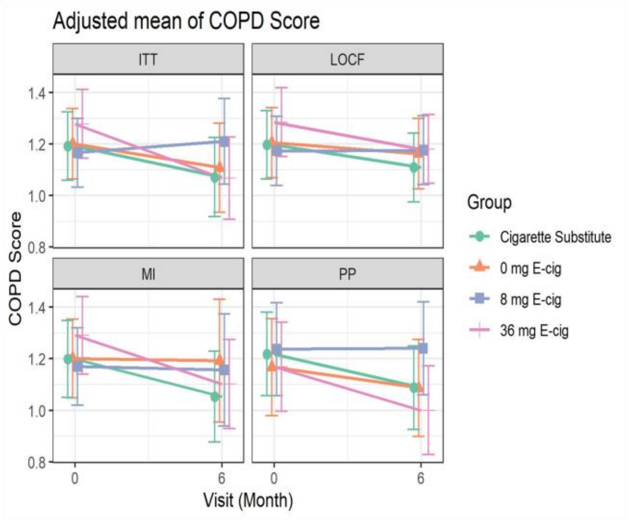

**C.** Unadjusted Mean

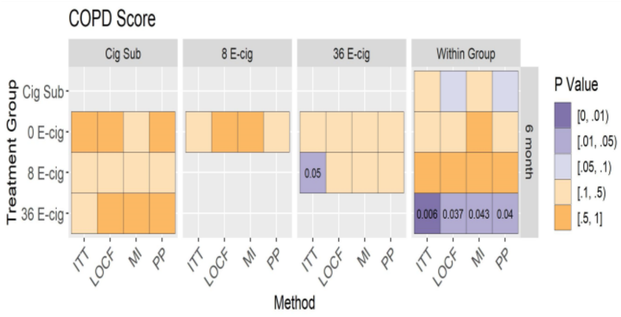

**D.** Adjusted Mean

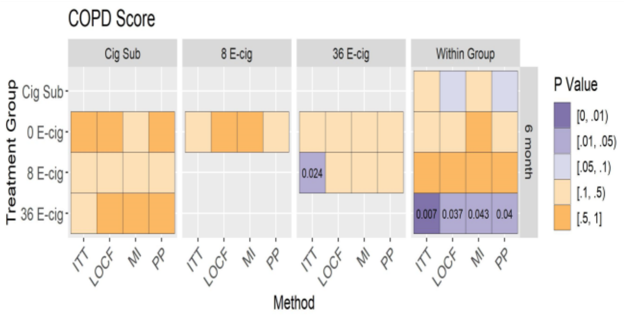

Figure A, B: Mean of total COPD score (95% CI) at baseline and 6 months (unadjusted and adjusted models) across randomized groups. Figure C, D: Between and within group differences in total COPD score at 6 months (unadjusted and adjusted models) among all randomized participants.

#### Supplemental Figure 19: Unadjusted and adjusted analyses of sodium

**A**

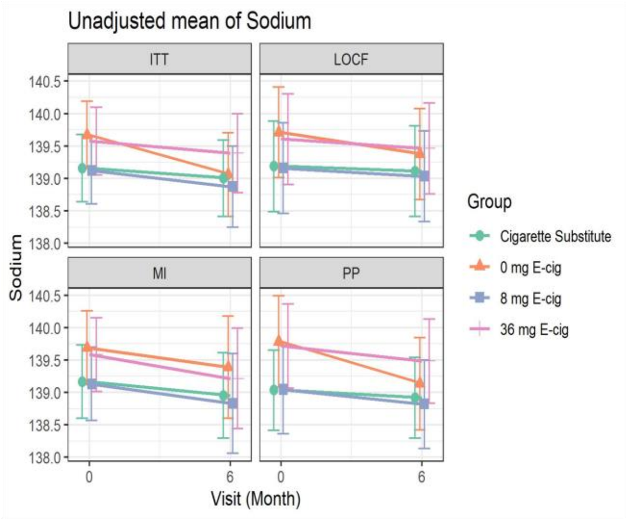

**B**

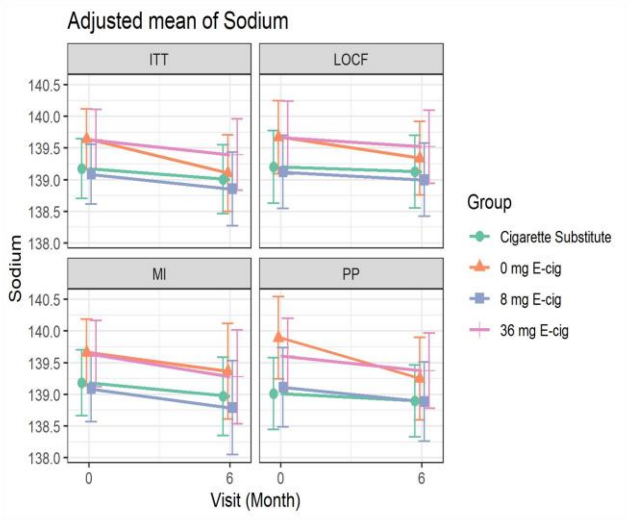

**C.** Unadjusted Mean

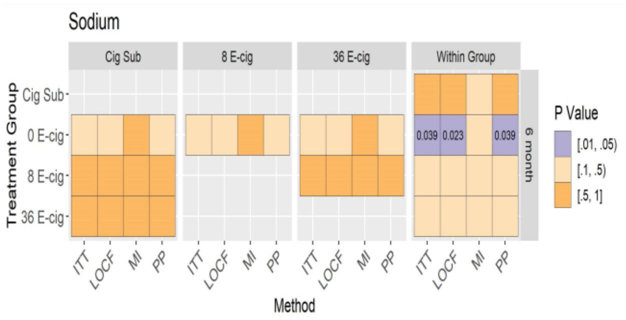

**D.** Adjusted Mean

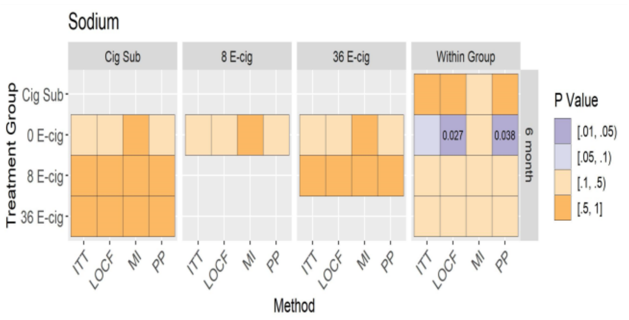

Figure A, B: Mean of sodium (95% CI) at baseline and 6 months (unadjusted and adjusted models) across randomized groups. Figure C, D: Between and within group differences in sodium at 6 months (unadjusted and adjusted models) among all randomized participants.

#### Supplemental Figure 20: Unadjusted and adjusted analyses of potassium

**A**

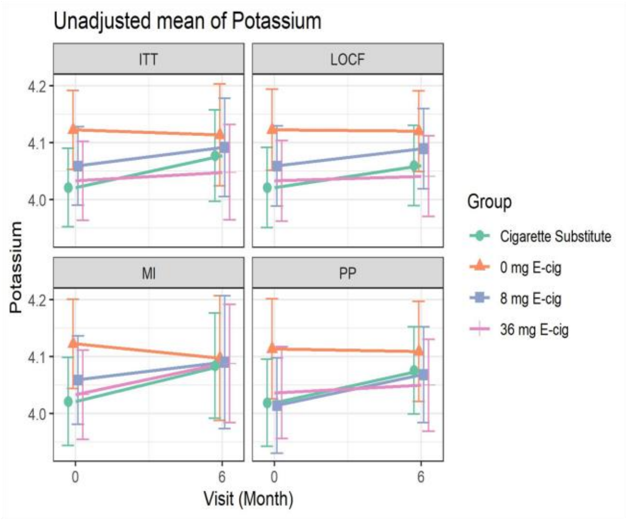

**B**

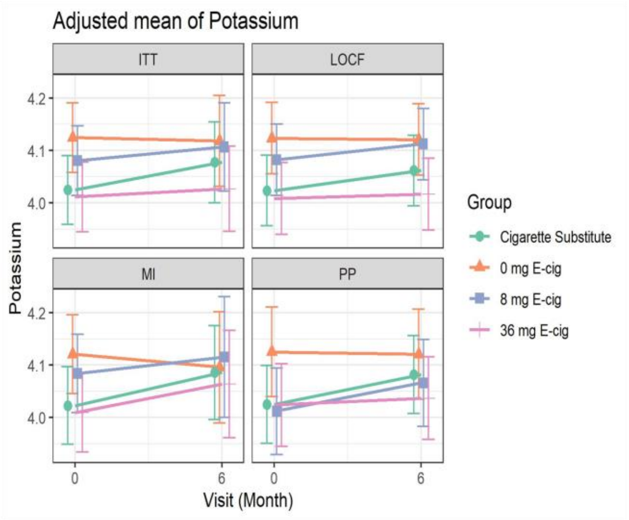

**C.** Unadjusted Mean

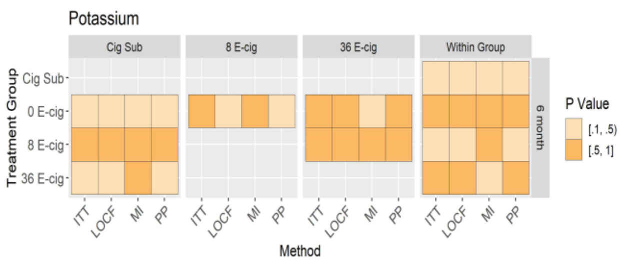

**D.** Adjusted Mean

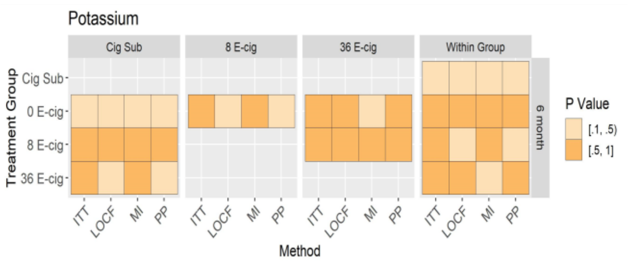

Figure A, B: Mean of potassium (95% CI) at baseline and 6 months (unadjusted and adjusted models) across randomized groups. Figure C, D: Between and within group differences in potassium at 6 months (unadjusted and adjusted models) among all randomized participants.

#### Supplemental Figure 21: Unadjusted and adjusted analyses of chloride

**A**

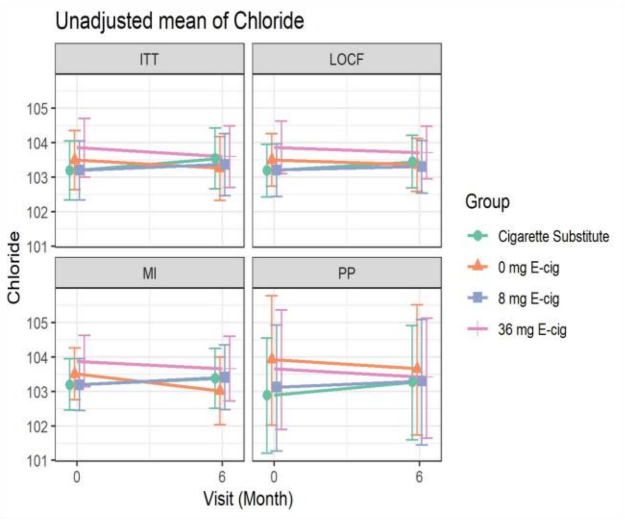

**B**

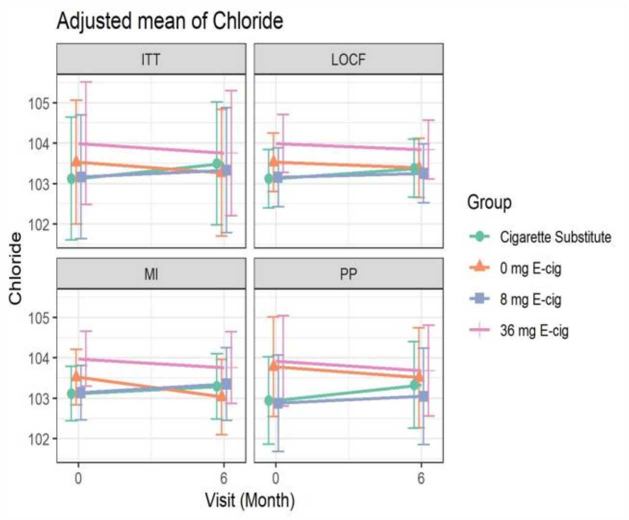

**C.** Unadjusted Mean

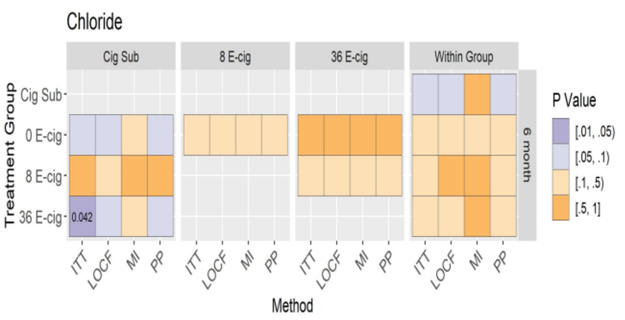

**D.** Adjusted Mean

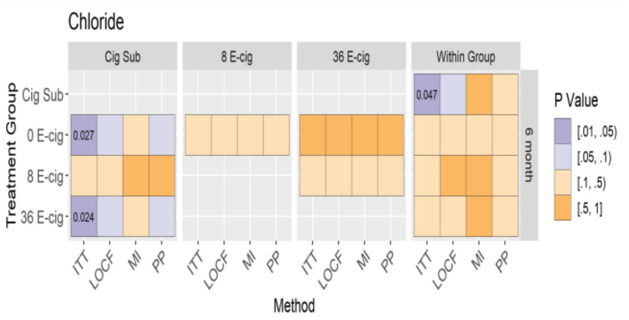

Figure A, B: Mean of chloride (95% CI) at baseline and 6 months (unadjusted and adjusted models) across randomized groups. Figure C, D: Between and within group differences in chloride at 6 months (unadjusted and adjusted models) among all randomized participants.

#### Supplemental Figure 22: Unadjusted and adjusted analyses of carbon dioxide

**A**

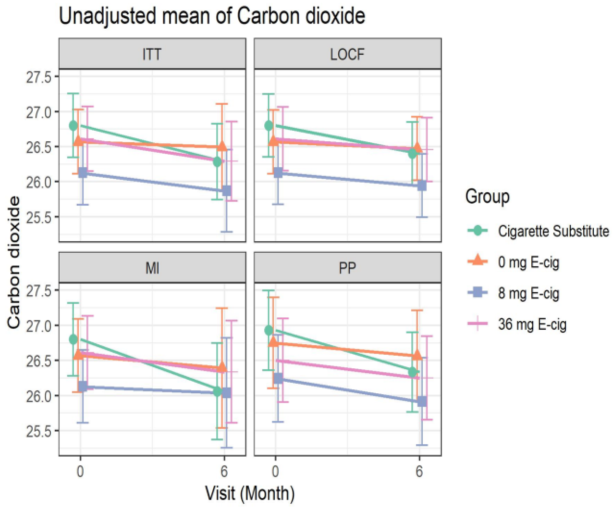

**B**

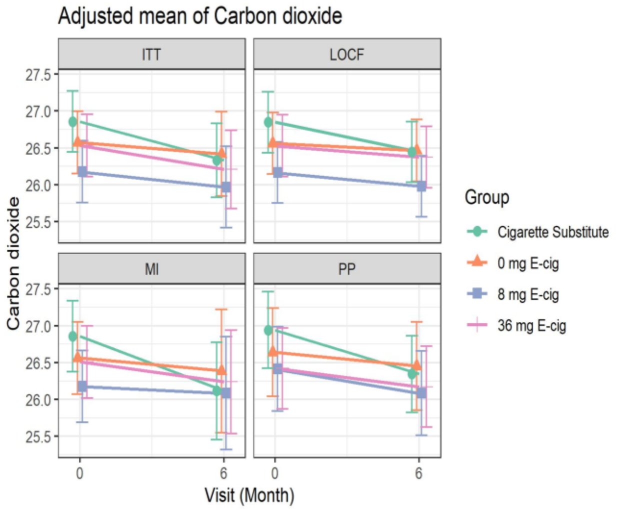

**C.** Unadjusted Mean

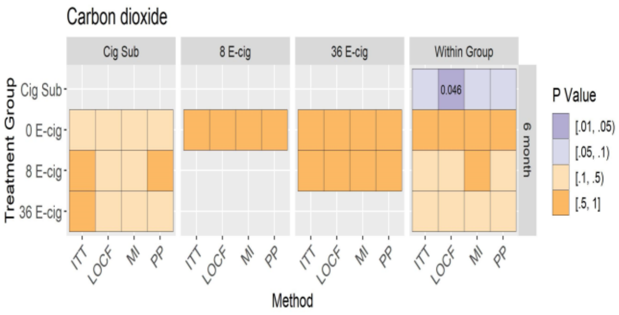

**D.** Adjusted Mean

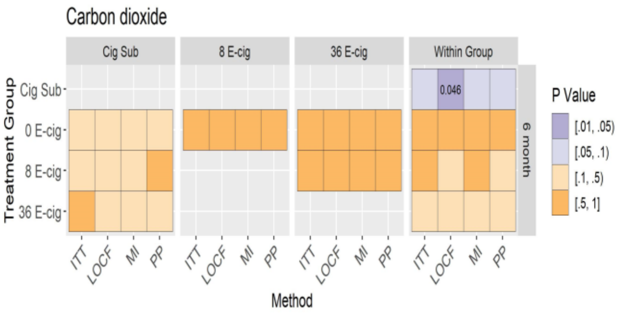

Figure A, B: Mean of carbon dioxide (95% CI) at baseline and 6 months (unadjusted and adjusted models) across randomized groups. Figure C, D: Between and within group differences in carbon dioxide at 6 months (unadjusted and adjusted models) among all randomized participants.

**Supplemental Figure 23: Unadjusted and adjusted analyses of anion gap**

**A**

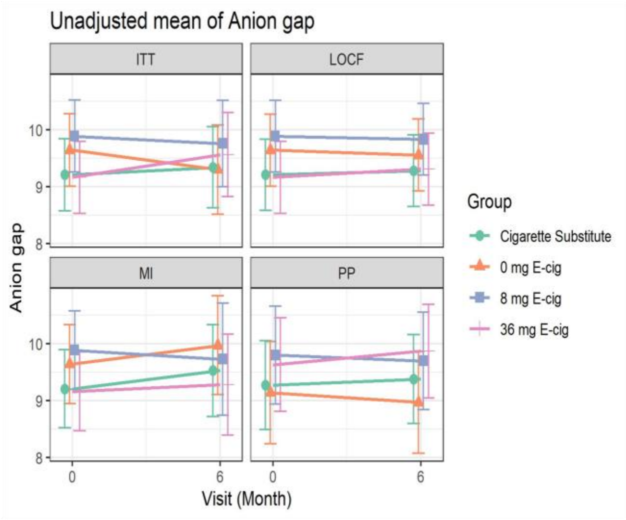

**B**

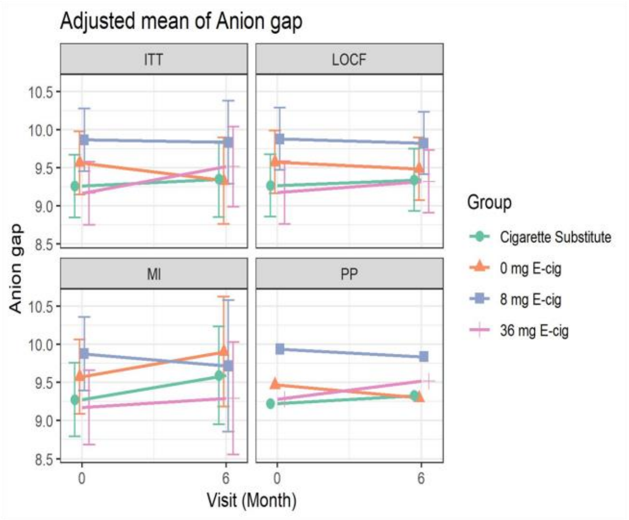

**C.** Unadjusted Mean

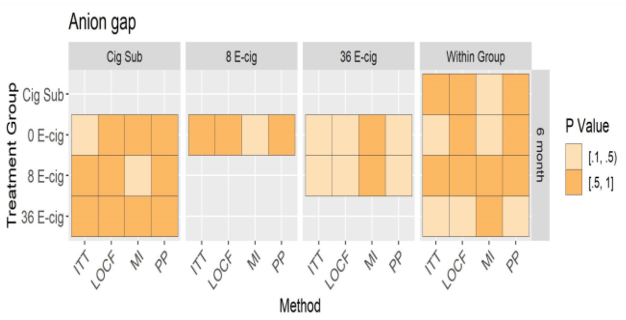

**D.** Adjusted Mean

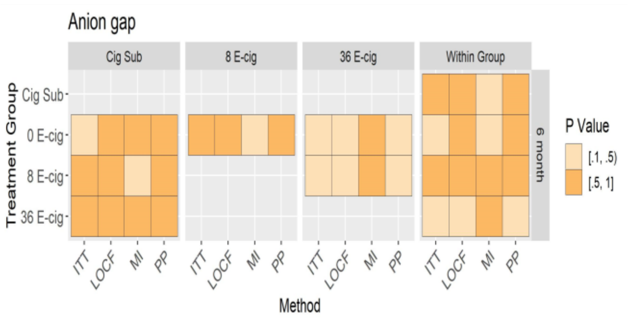

Figure A, B: Mean of anion gap (95% CI) at baseline and 6 months (unadjusted and adjusted models) across randomized groups. Figure C, D: Between and within group differences in anion gap at 6 months (unadjusted and adjusted models) among all randomized participants.

### Supplemental Figure 24: Unadjusted and adjusted analyses of glucose

**A**

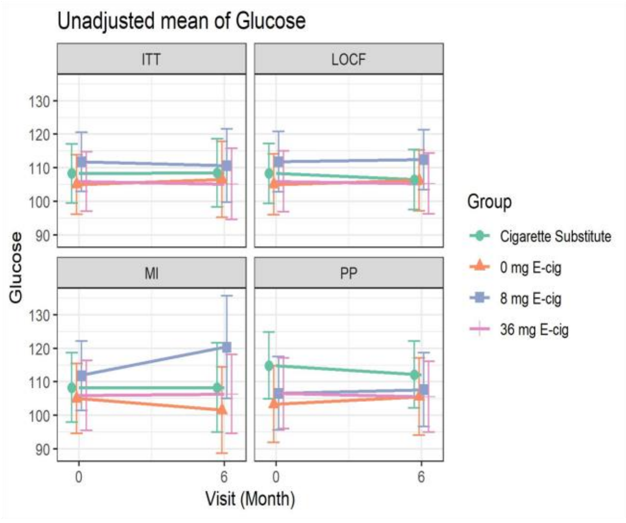

**B**

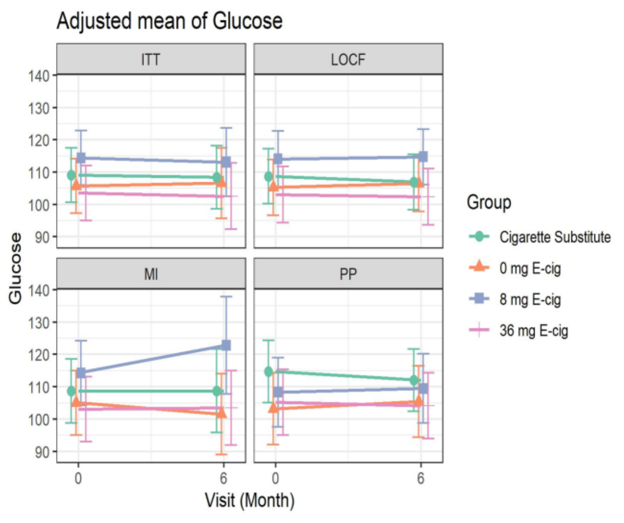

**C.** Unadjusted Mean

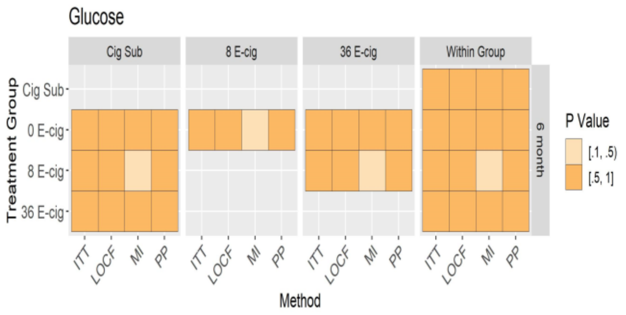

**D.** Adjusted Mean

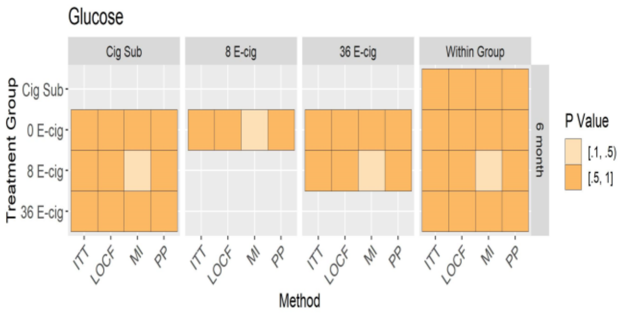

Figure A, B: Mean of glucose (95% CI) at baseline and 6 months (unadjusted and adjusted models) across randomized groups. Figure C, D: Between and within group differences in glucose at 6 months (unadjusted and adjusted models) among all randomized participants.

**Supplemental Figure 25: Unadjusted and adjusted analyses of BUN**

**A**

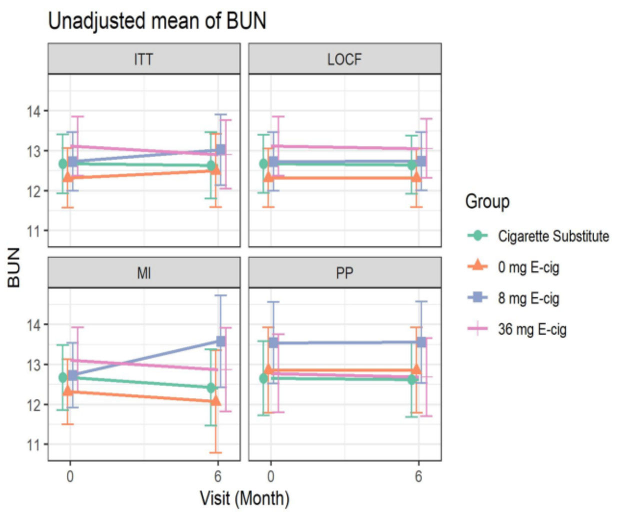

**B**

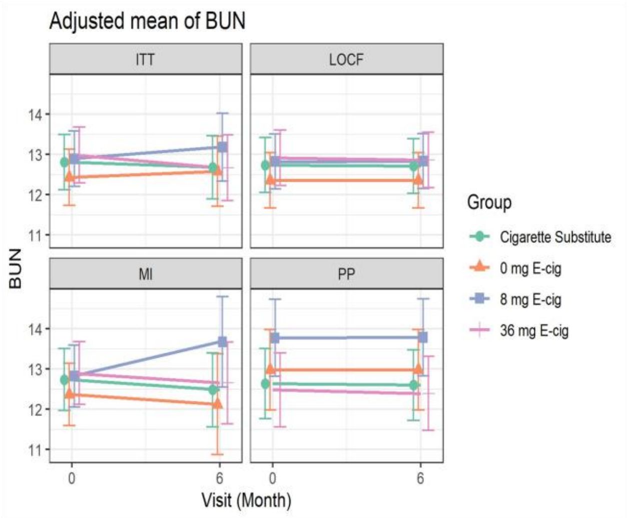

**C.** Unadjusted Mean

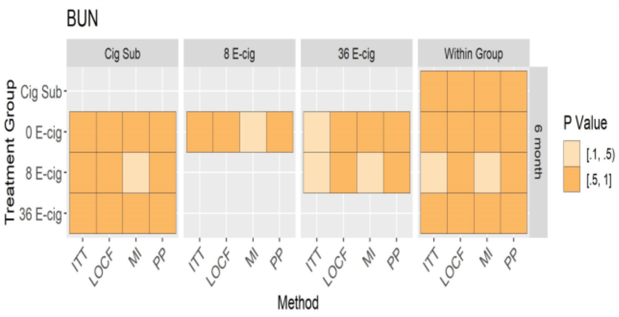

**D.** Adjusted Mean

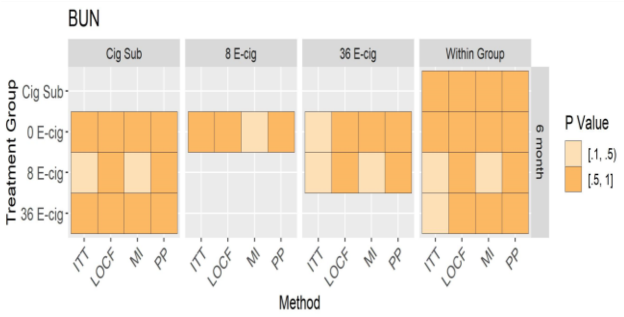

Figure A, B: Mean of BUN (95% CI) at baseline and 6 months (unadjusted and adjusted models) across randomized groups. Figure C, D: Between and within group differences in BUN at 6 months (unadjusted and adjusted models) among all randomized participants.

**Supplemental Figure 26: Unadjusted and adjusted analyses of AST**

**A**

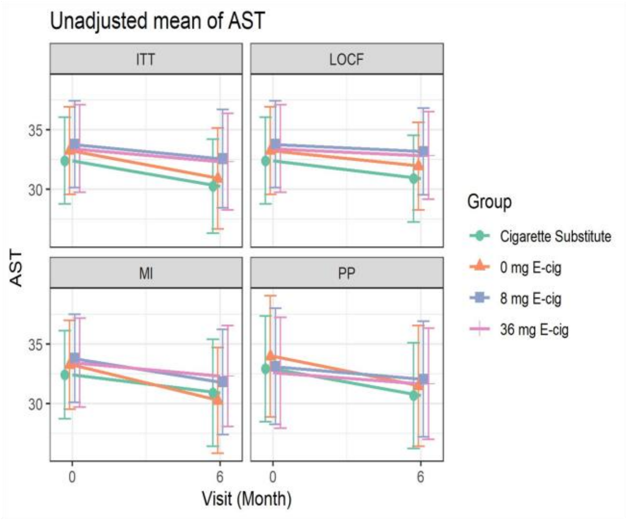

**B**

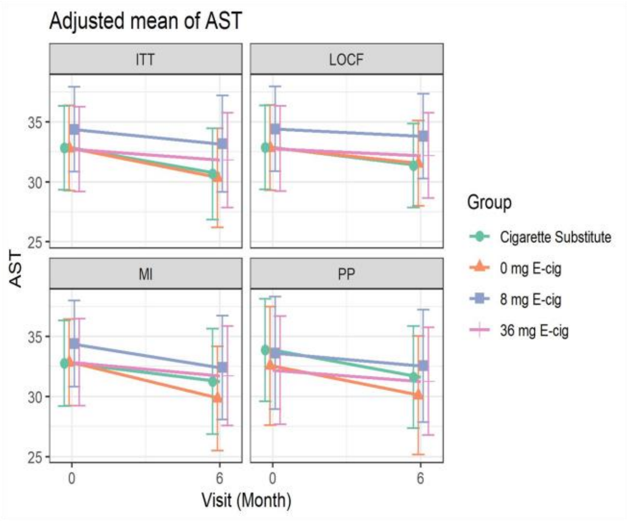

**C.** Unadjusted Mean

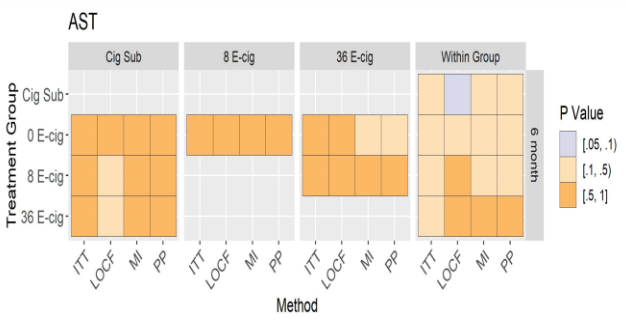

**D.** Adjusted Mean

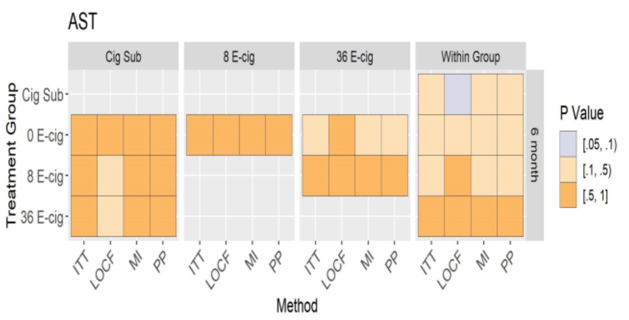

Figure A, B: Mean of AST (95% CI) at baseline and 6 months (unadjusted and adjusted models) across randomized groups. Figure C, D: Between and within group differences in AST at 6 months (unadjusted and adjusted models) among all randomized participants.

**Supplemental Figure 27: Unadjusted and adjusted analyses of ALT**

**A**

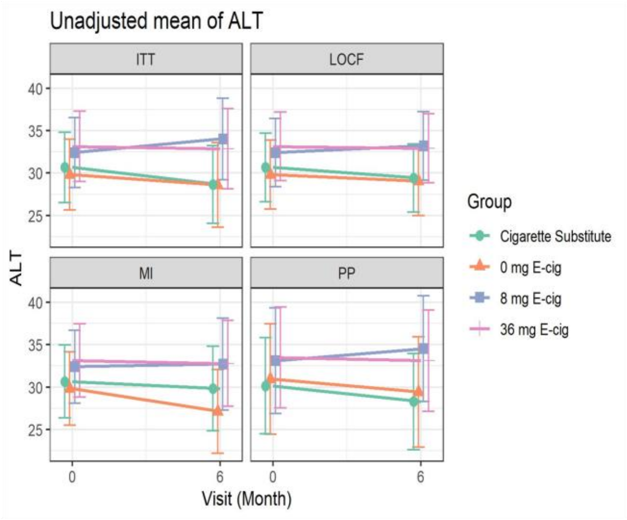

**B**

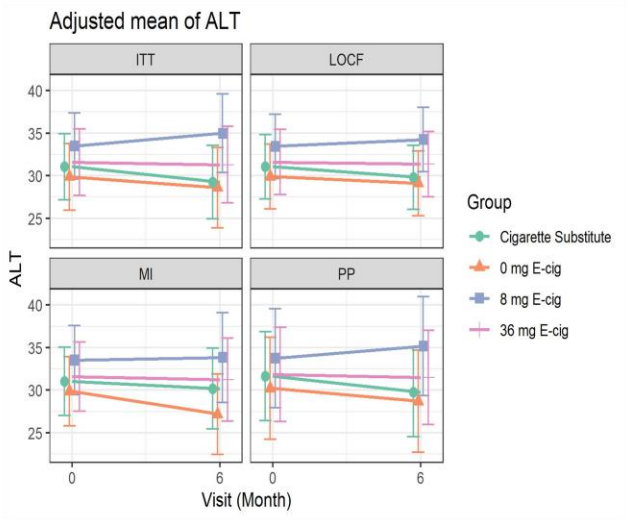

**C.** Unadjusted Mean

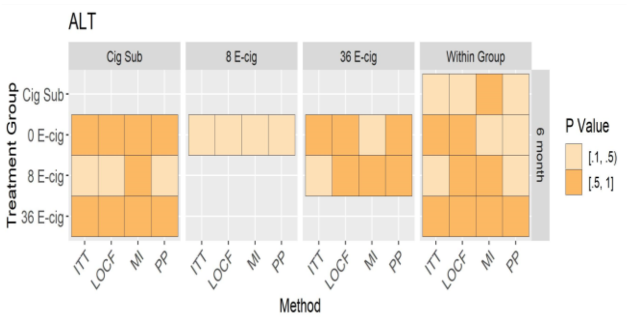

**D.** Adjusted Mean

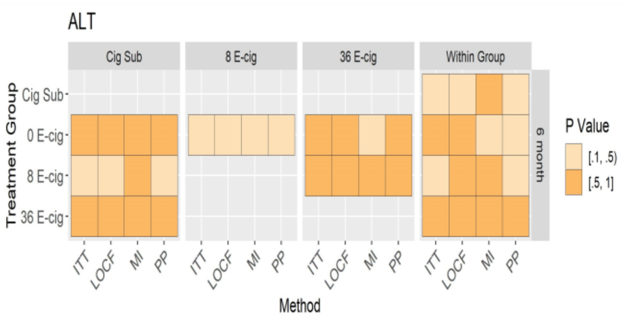

Figure A, B: Mean of ALT (95% CI) at baseline and 6 months (unadjusted and adjusted models) across randomized groups. Figure C, D: Between and within group differences in ALT at 6 months (unadjusted and adjusted models) among all randomized participants.

#### Supplemental Figure 28: Unadjusted and adjusted analyses of alkaline phosphatase

**A**

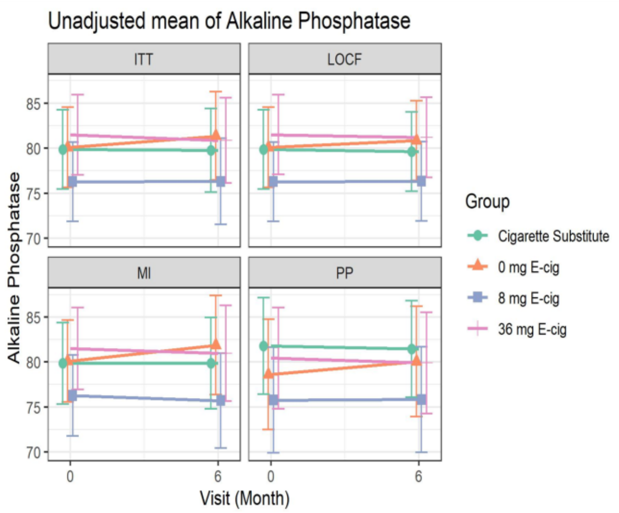

**B**

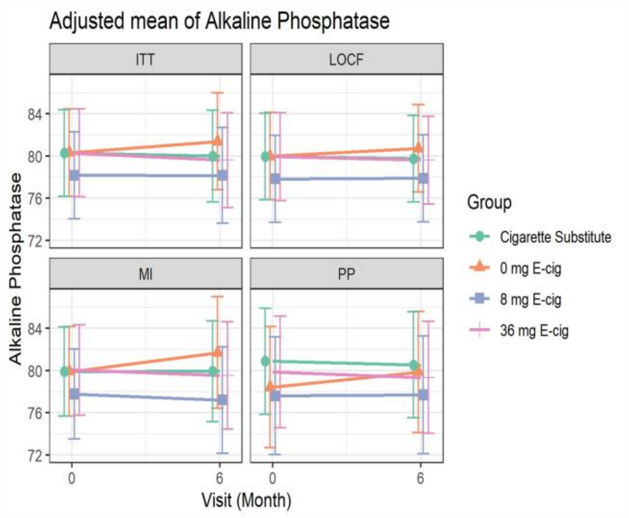

**C.** Unadjusted Mean

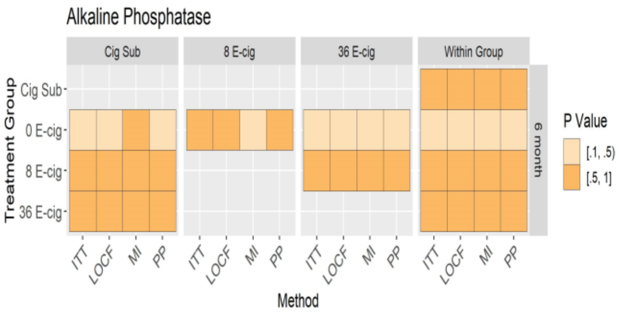

**D.** Adjusted Mean

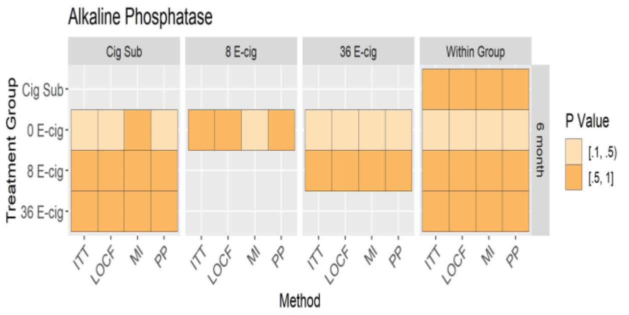

Figure A, B: Mean of alkaline phosphatase (95% CI) at baseline and 6 months (unadjusted and adjusted models) across randomized groups. Figure C, D: Between and within group differences in alkaline phosphatase at 6 months (unadjusted and adjusted models) among all randomized participants.

#### Supplemental Figure 29: Unadjusted and adjusted analyses of bilirubin

**A**

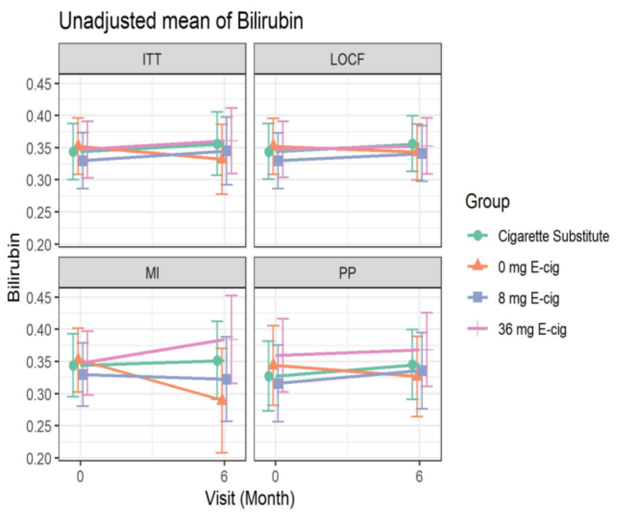

**B**

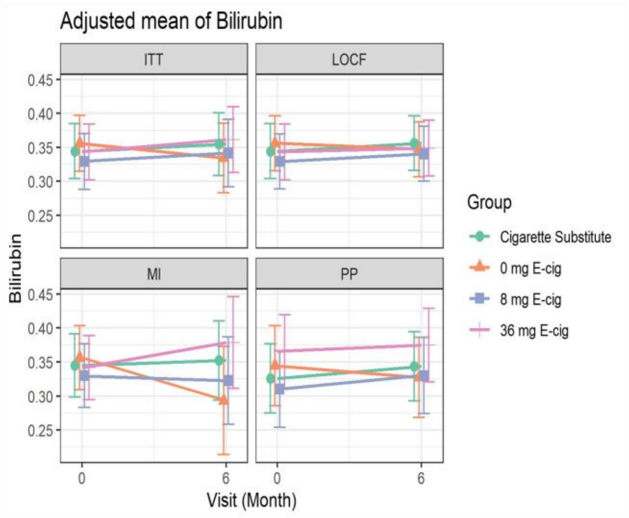

**C.** Unadjusted Mean

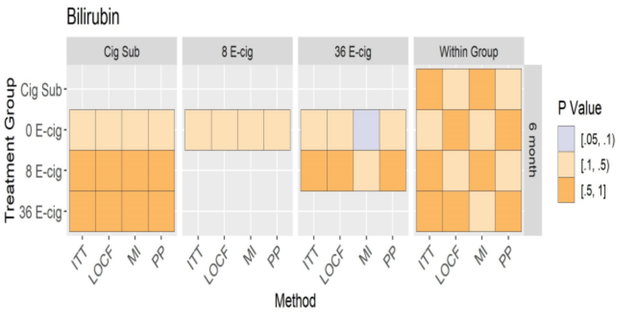

**D.** Adjusted Mean

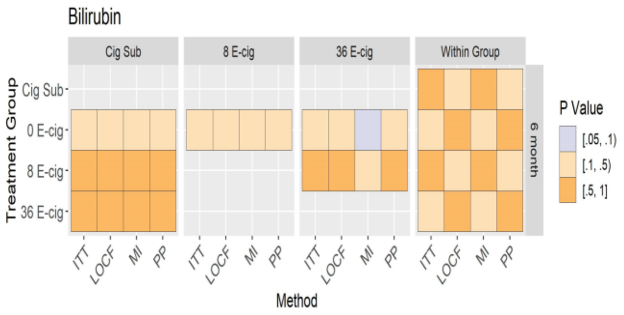

Figure A, B: Mean of bilirubin (95% CI) at baseline and 6 months (unadjusted and adjusted models) across randomized groups. Figure C, D: Between and within group differences in bilirubin at 6 months (unadjusted and adjusted models) among all randomized participants.

**Supplemental Figure 30: Unadjusted and adjusted analyses of protein**

**A**

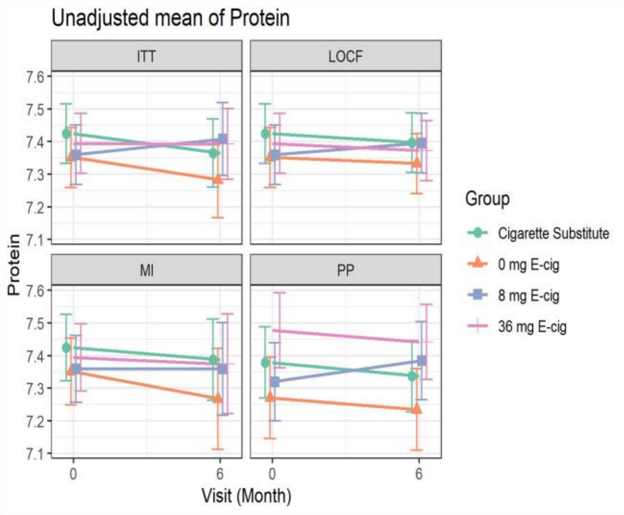

**B**

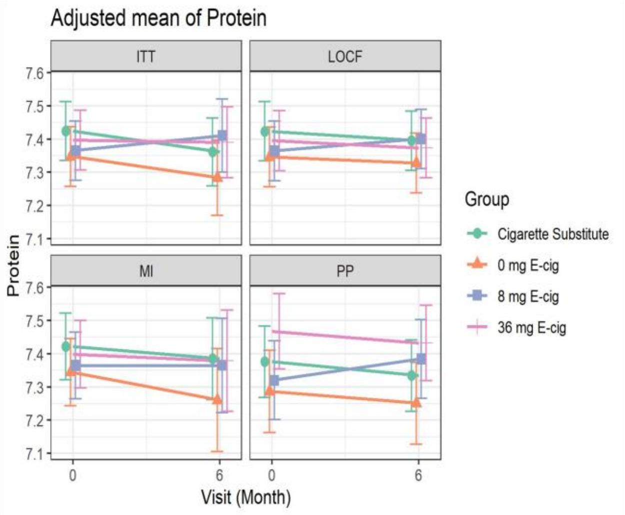

**C.** Unadjusted Mean

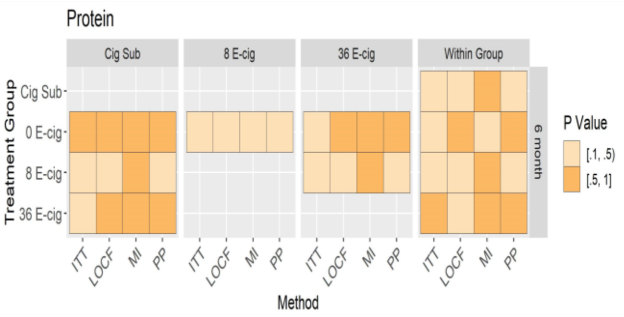

**D.** Adjusted Mean

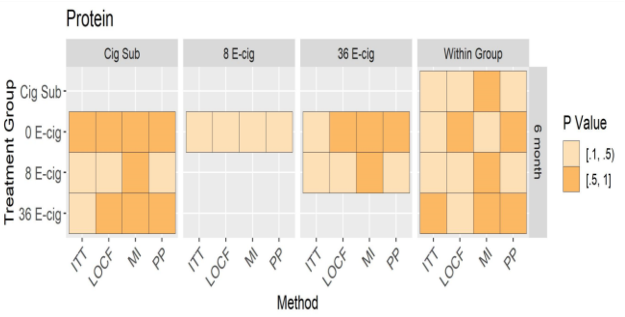

Figure A, B: Mean of protein (95% CI) at baseline and 6 months (unadjusted and adjusted models) across randomized groups. Figure C, D: Between and within group differences in protein at 6 months (unadjusted and adjusted models) among all randomized participants.

#### Supplemental Figure 31: Unadjusted and adjusted analyses of albumin

**A**

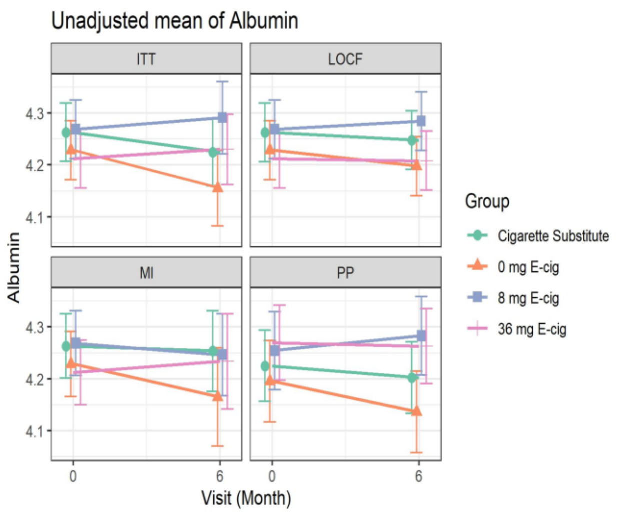

**B**

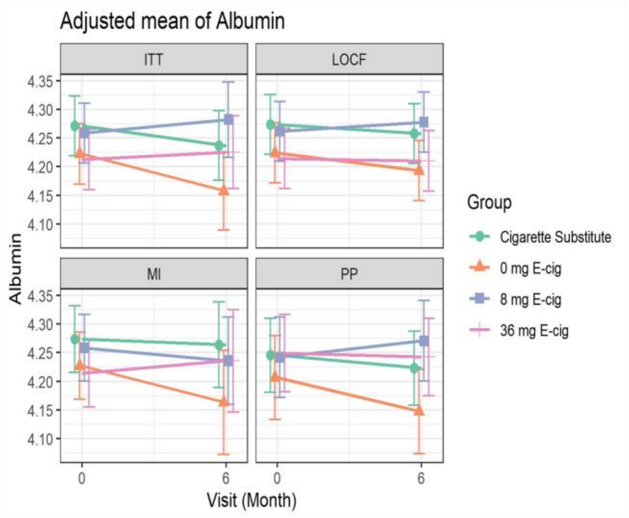

**C.** Unadjusted Mean

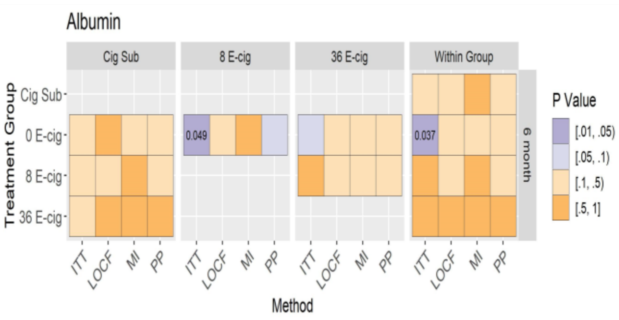

**D.** Adjusted Mean

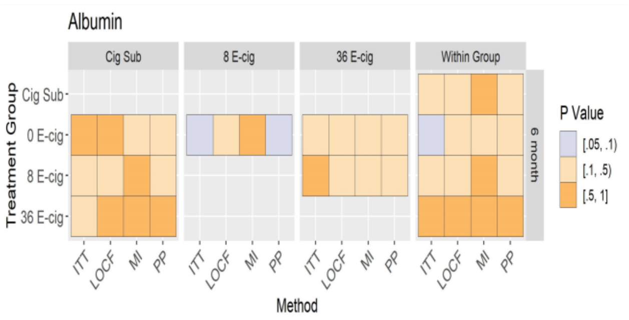

Figure A, B: Mean of albumin (95% CI) at baseline and 6 months (unadjusted and adjusted models) across randomized groups. Figure C, D: Between and within group differences in albumin at 6 months (unadjusted and adjusted models) among all randomized participants.

#### Supplemental Figure 32: Unadjusted and adjusted analyses of calcium

**A**

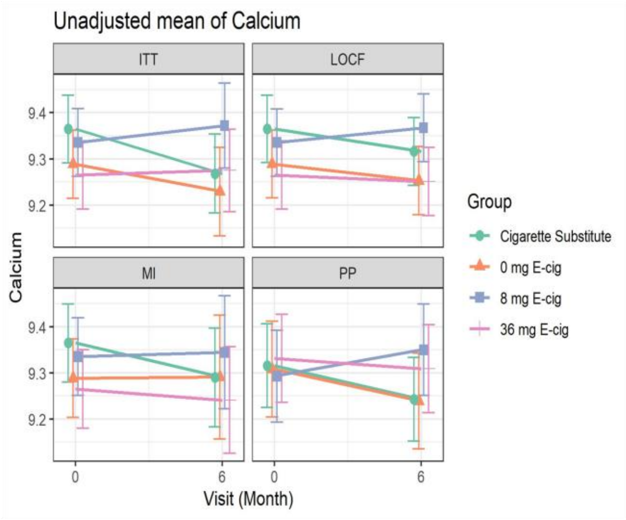

**B**

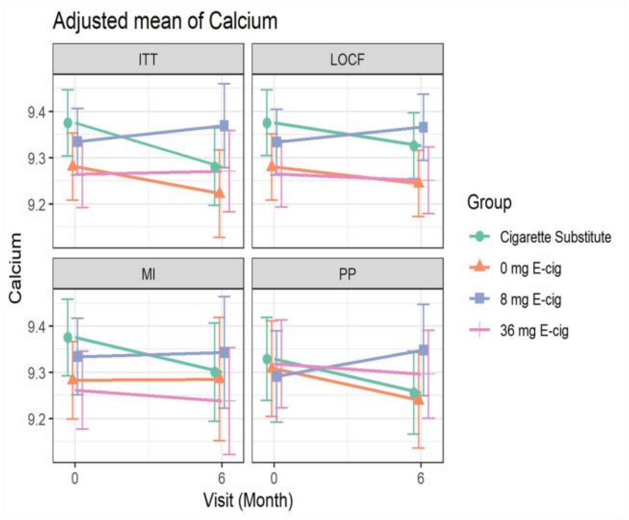

**C.** Unadjusted Mean

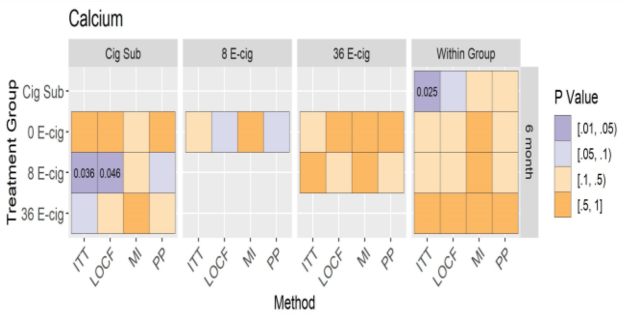

**D.** Adjusted Mean

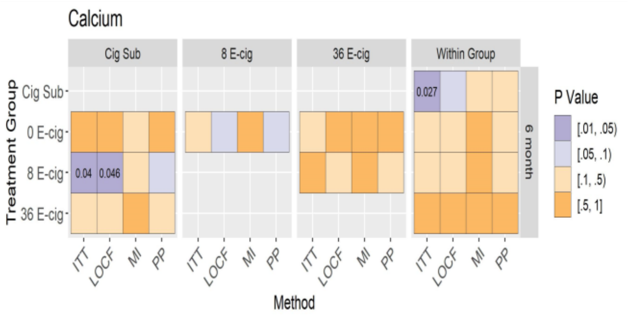

Figure A, B: Mean of calcium (95% CI) at baseline and 6 months (unadjusted and adjusted models) across randomized groups. Figure C, D: Between and within group differences in calcium at 6 months (unadjusted and adjusted models) among all randomized participants.

**Supplemental Figure 33: Unadjusted and adjusted analyses of C-reactive protein**

**A**

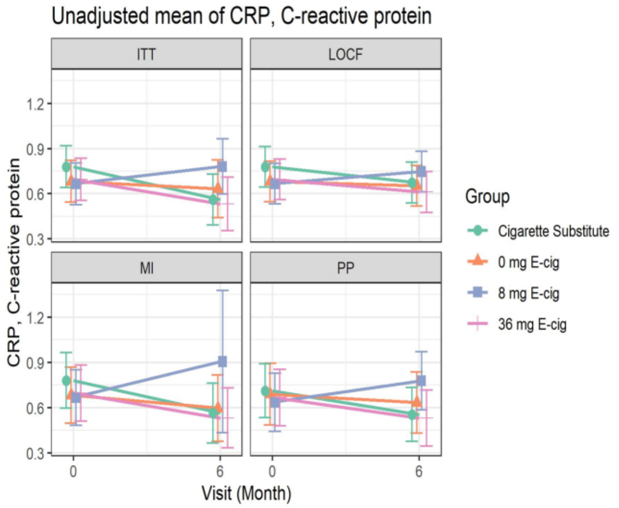

**B**

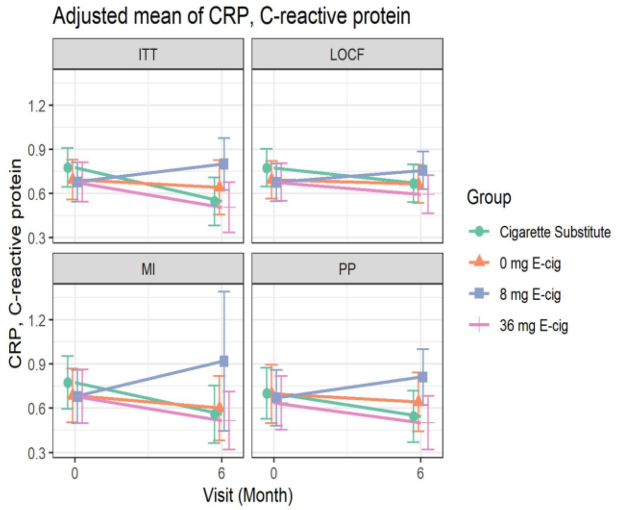

**C.** Unadjusted Mean

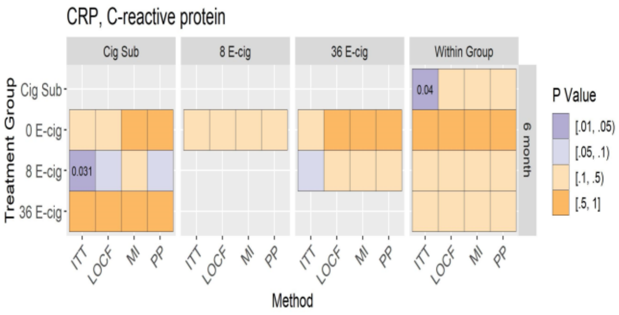

**D.** Adjusted Mean

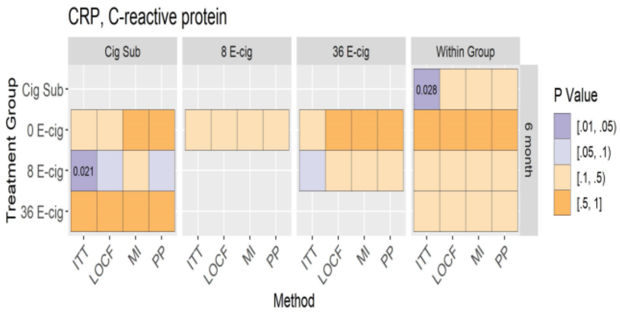

Figure A, B: Mean of C-reactive protein (95% CI) at baseline and 6 months (unadjusted and adjusted models) across randomized groups. Figure C, D: Between and within group differences in C-reactive protein at 6 months (unadjusted and adjusted models) among all randomized participants.

#### Supplemental Figure 34: Unadjusted and adjusted analyses of Creatinine

A

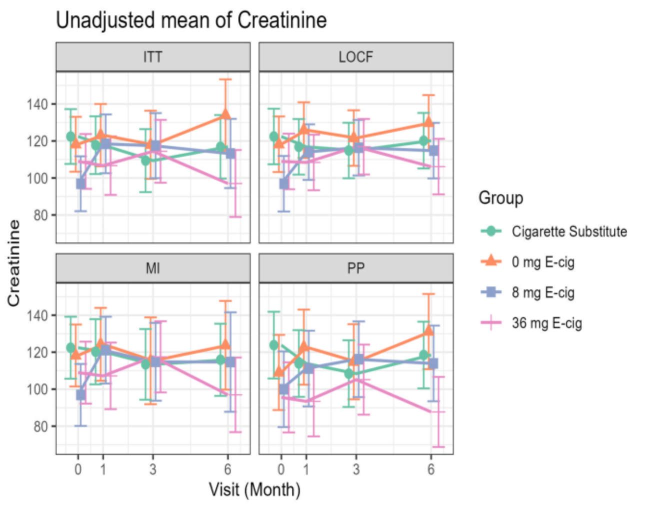

B

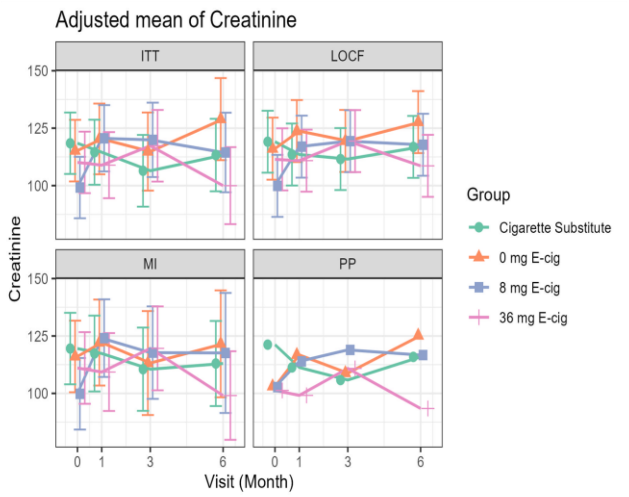

**C.** Unadjusted Mean

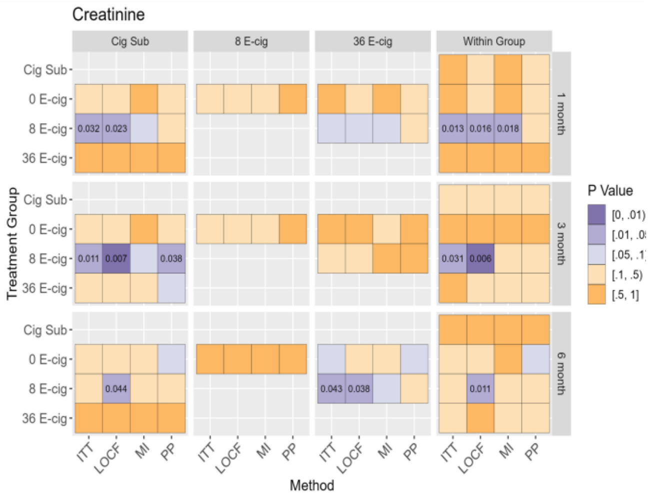

**D.** Adjusted Mean

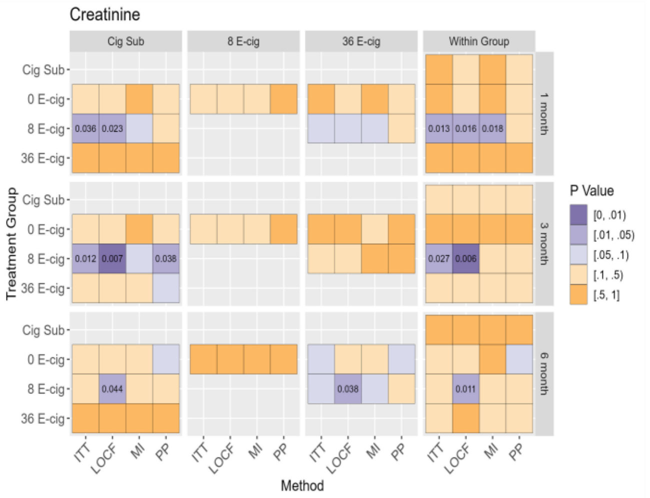

Figure A, B: Mean of urinary Creatinine (95% CI) at baseline, 1, 3 and 6 months (unadjusted and adjusted models) across randomized groups. Figure C, D: Between and within group differences in C-reactive protein at baseline, 1, 3, and 6 months (unadjusted and adjusted models) among all randomized participants.

**Supplemental Figure 35: Unadjusted and adjusted analyses of white blood cells**

**A**

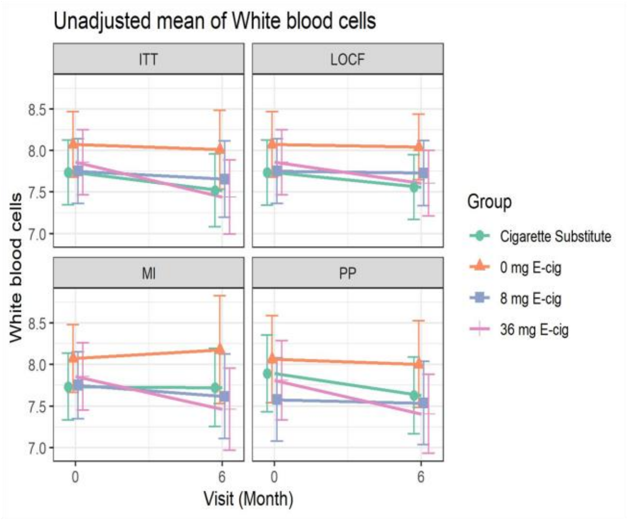

**B**

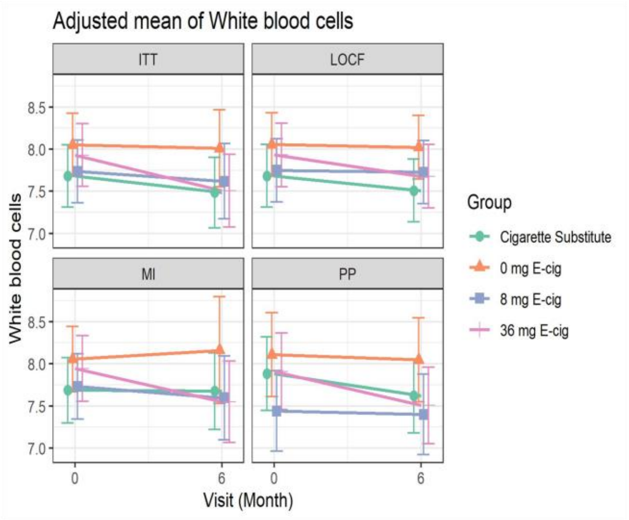

**C.** Unadjusted Mean

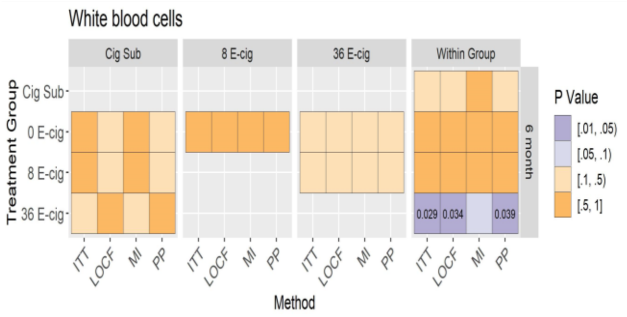

**D.** Adjusted Mean

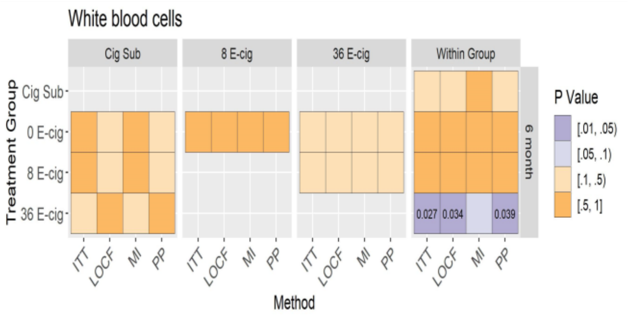

Figure A, B: Mean of white blood cells (95% CI) at baseline and 6 months (unadjusted and adjusted models) across randomized groups. Figure C, D: Between and within group differences in white blood cells at 6 months (unadjusted and adjusted models) among all randomized participants.

#### Supplemental Figure 36: Unadjusted and adjusted analyses of red blood cells

**A**

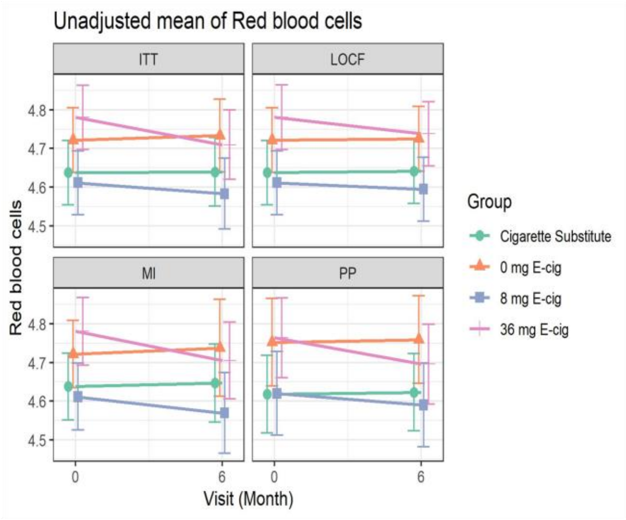

**B**

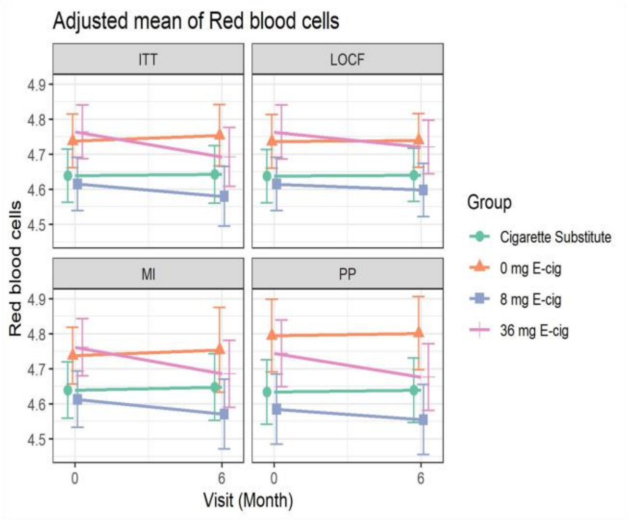

**C.** Unadjusted Mean

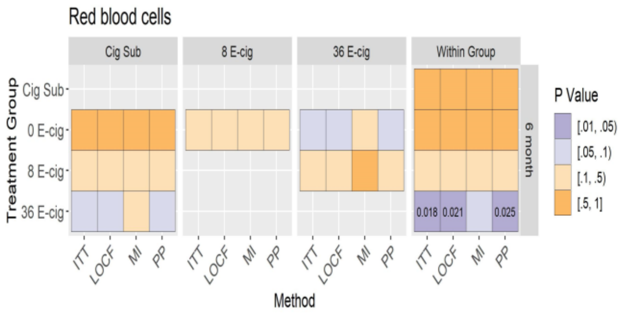

**D.** Adjusted Mean

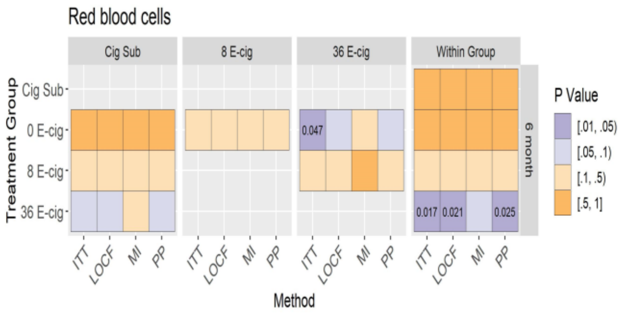

Figure A, B: Mean of red blood cells (95% CI) at baseline and 6 months (unadjusted and adjusted models) across randomized groups. Figure C, D: Between and within group differences in red blood cells at 6 months (unadjusted and adjusted models) among all randomized participants.

### Supplemental Figure 37: Unadjusted and adjusted analyses of hemoglobin

**A**

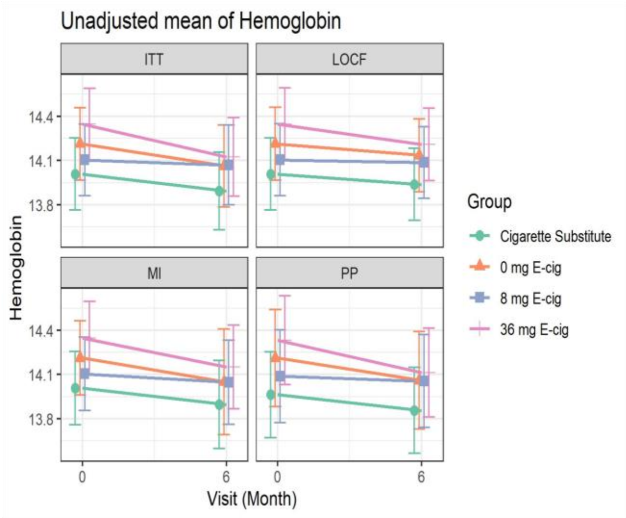

**B**

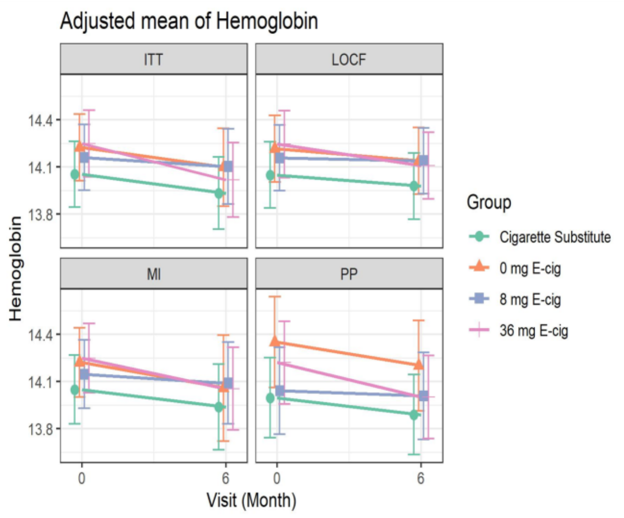

**C.** Unadjusted Mean

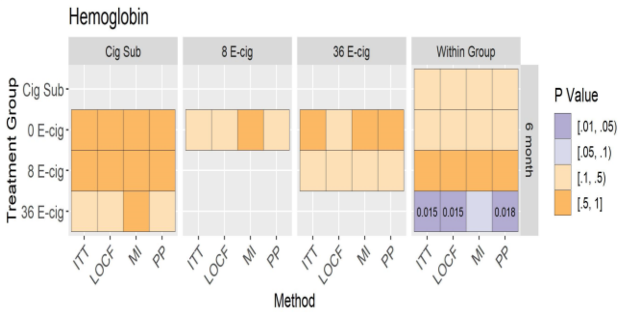

**D.** Adjusted Mean

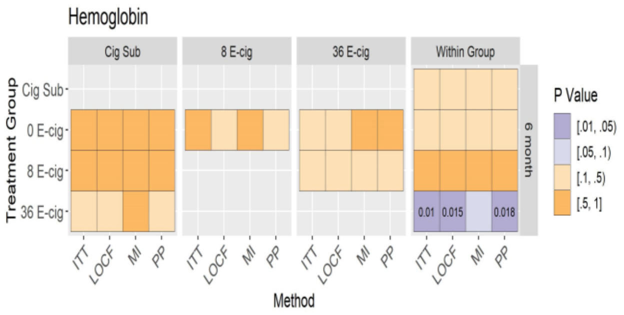

Figure A, B: Mean of hemoglobin (95% CI) at baseline and 6 months (unadjusted and adjusted models) across randomized groups. Figure C, D: Between and within group differences in hemoglobin at 6 months (unadjusted and adjusted models) among all randomized participants.

#### Supplemental Figure 38: Unadjusted and adjusted analyses of hematocrit

**A**

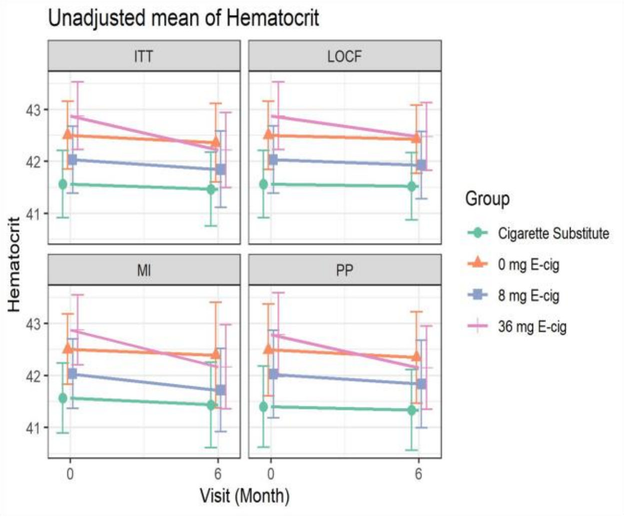

**B**

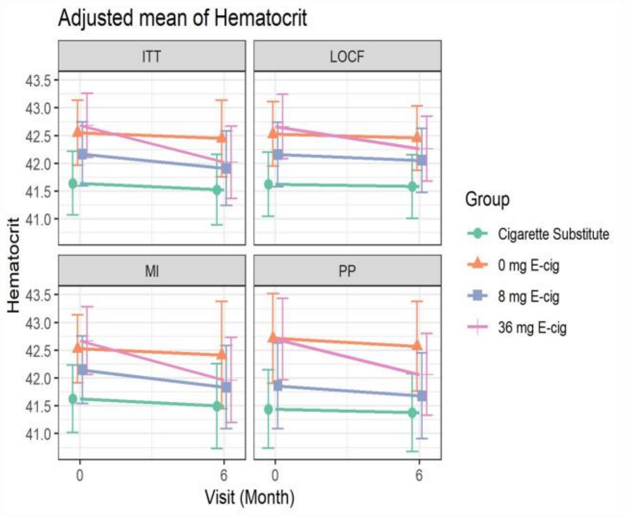

**C.** Unadjusted Mean

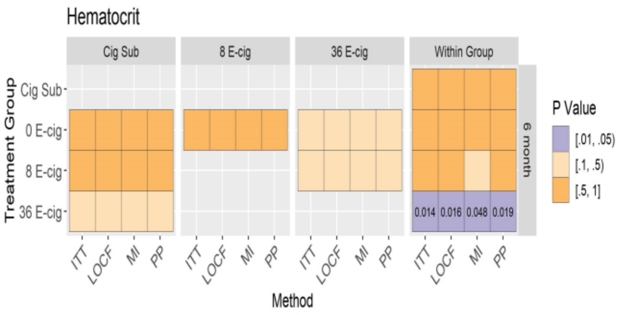

**D.** Adjusted Mean

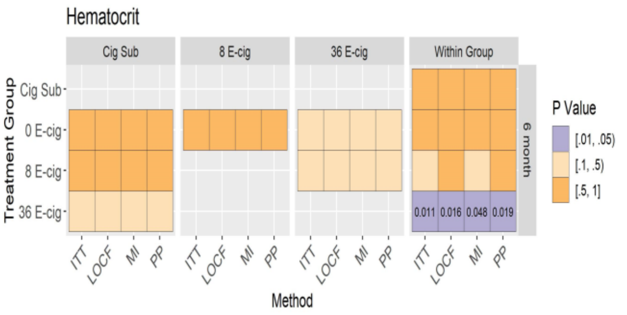

Figure A, B: Mean of hematocrit (95% CI) at baseline and 6 months (unadjusted and adjusted models) across randomized groups. Figure C, D: Between and within group differences in hematocrit at 6 months (unadjusted and adjusted models) among all randomized participants.

### Supplemental Figure 39: Unadjusted and adjusted analyses of MCV

**A**

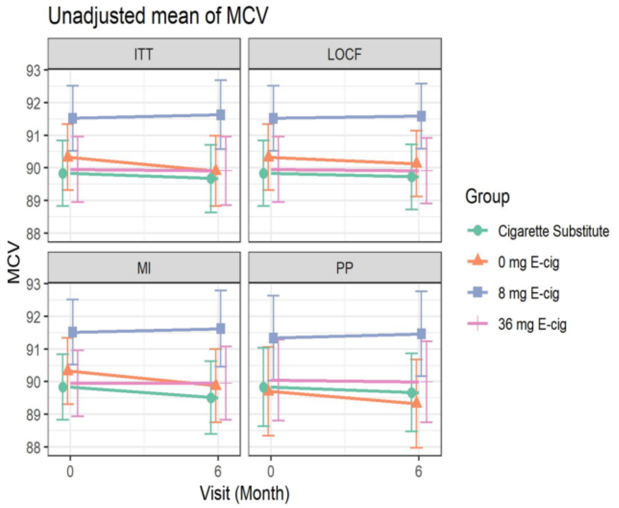

**B**

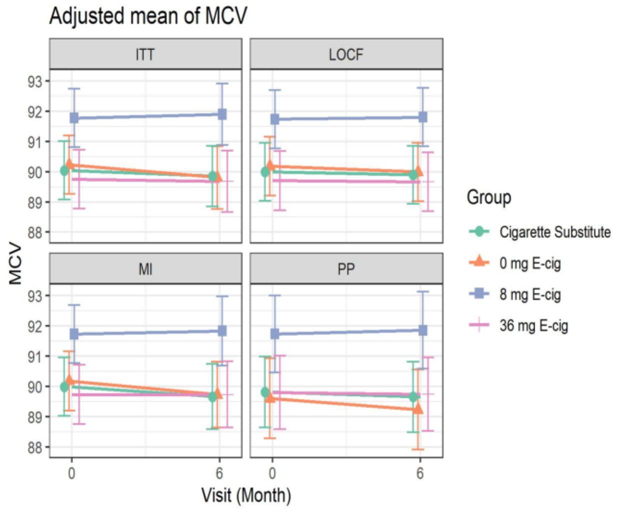

**C.** Unadjusted Mean

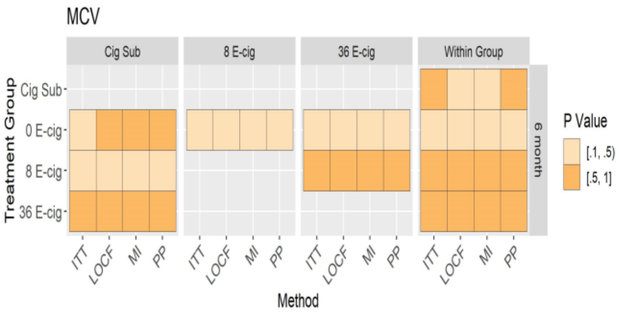

**D.** Adjusted Mean

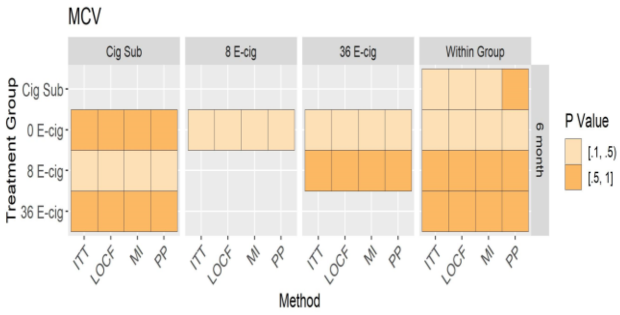

Figure A, B: Mean of MCV (95% CI) at baseline and 6 months (unadjusted and adjusted models) across randomized groups. Figure C, D: Between and within group differences in MCV at 6 months (unadjusted and adjusted models) among all randomized participants.

#### Supplemental Figure 40: Unadjusted and adjusted analyses of MCH

**A**

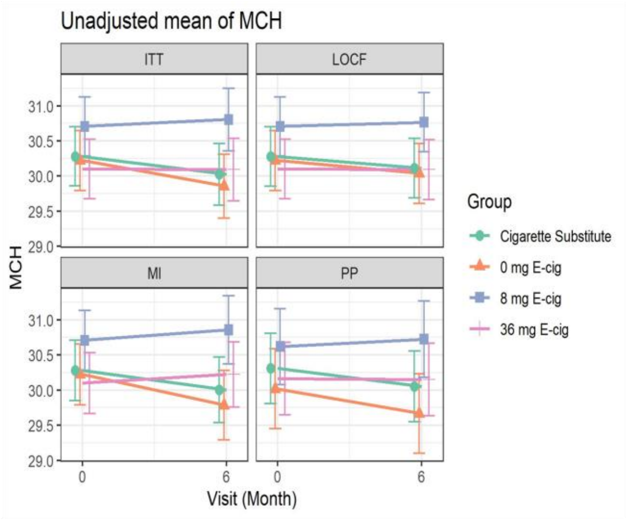

**B**

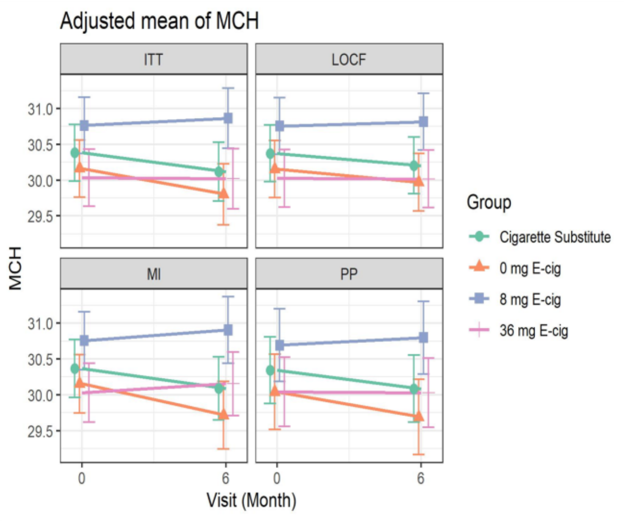

**C.** Unadjusted Mean

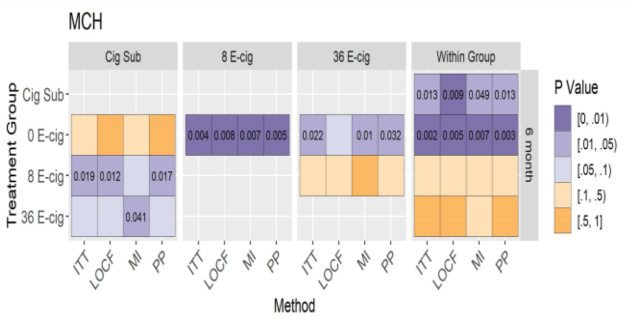

**D.** Adjusted Mean

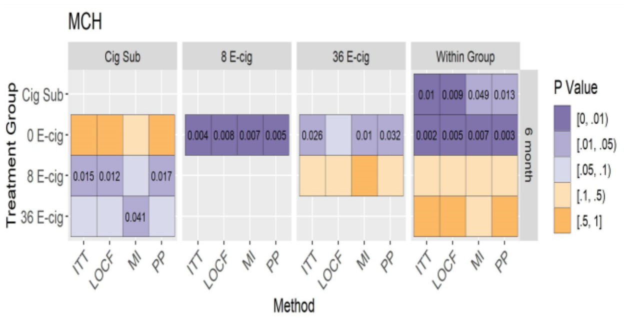

Figure A, B: Mean of MCH (95% CI) at baseline and 6 months (unadjusted and adjusted models) across randomized groups. Figure C, D: Between and within group differences in MCH at 6 months (unadjusted and adjusted models) among all randomized participants.

**Supplemental Figure 41: Unadjusted and adjusted analyses of MCHC**

**A**

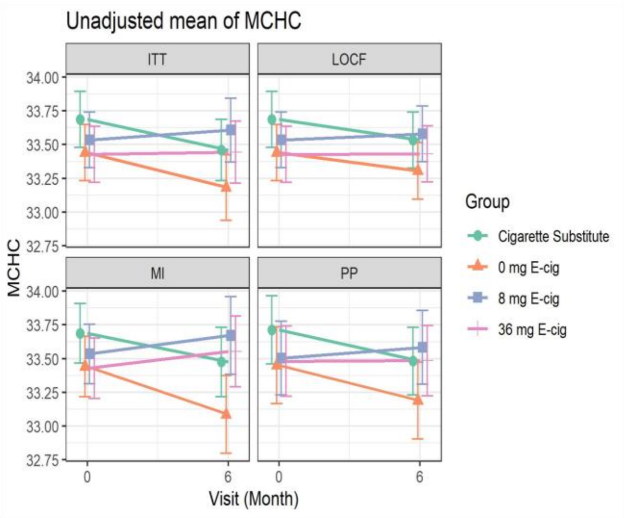

**B**

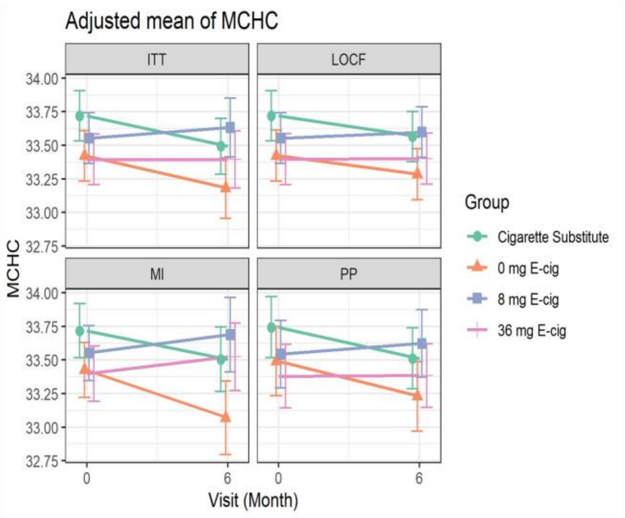

**C.** Unadjusted Mean

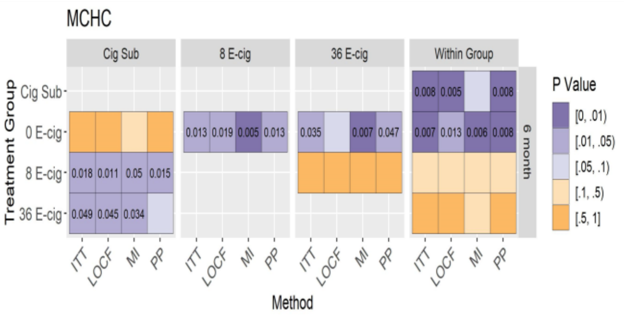

**D.** Adjusted Mean

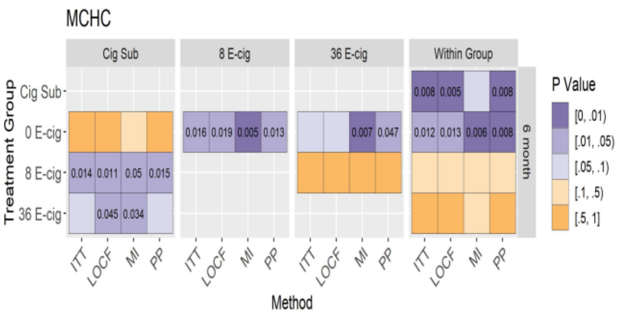

Figure A, B: Mean of MCHC (95% CI) at baseline and 6 months (unadjusted and adjusted models) across randomized groups. Figure C, D: Between and within group differences in MCHC at 6 months (unadjusted and adjusted models) among all randomized participants.

### Supplemental Figure 42: Unadjusted and adjusted analyses of RDW

**A**

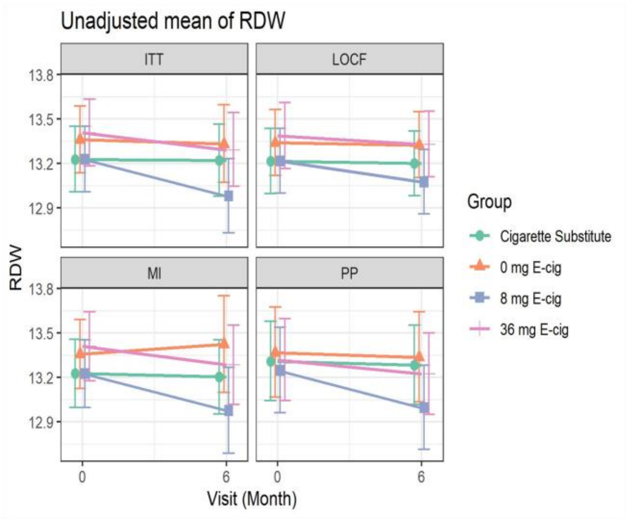

**B**

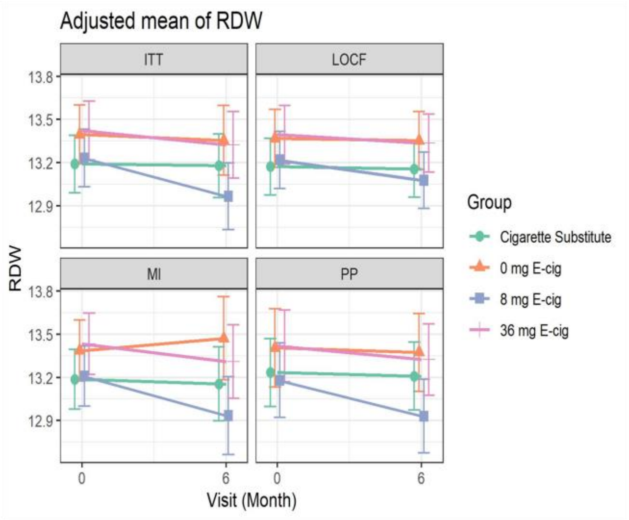

**C.** Unadjusted Mean

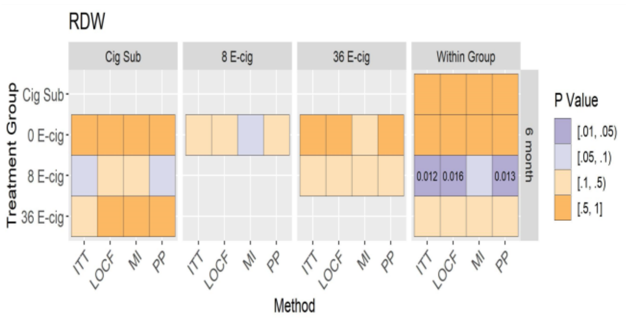

**D.** Adjusted Mean

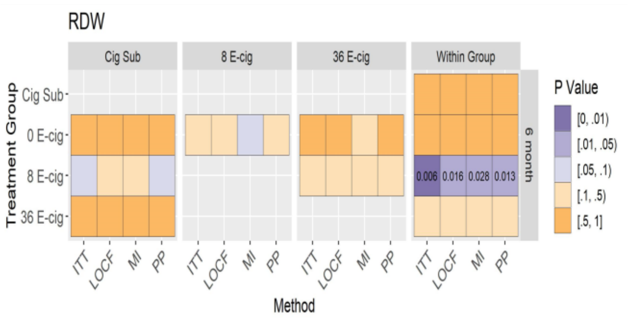

Figure A, B: Mean of RDW (95% CI) at baseline and 6 months (unadjusted and adjusted models) across randomized groups. Figure C, D: Between and within group differences in RDW at 6 months (unadjusted and adjusted models) among all randomized participants.

**Supplemental Figure 43: Unadjusted and adjusted analyses of platelets**

**A**

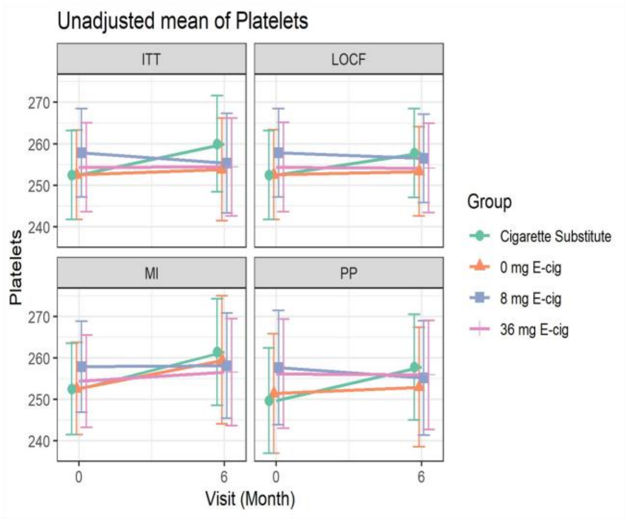

**B**

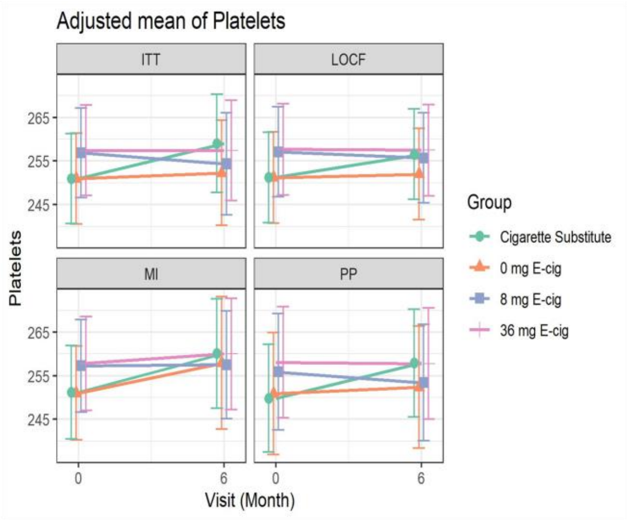

**C.** Unadjusted Mean

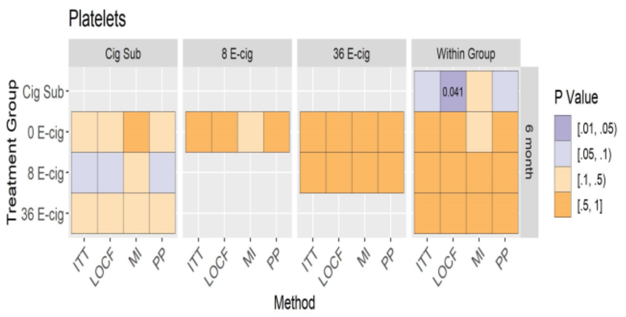

**D.** Adjusted Mean

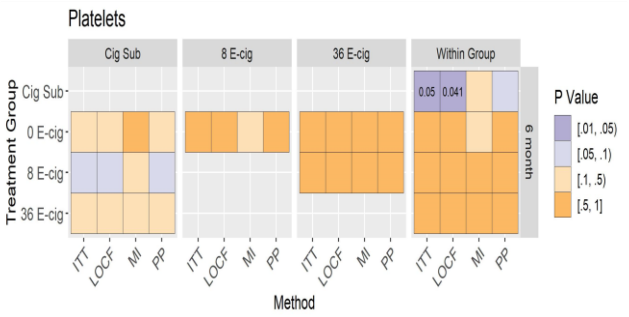

Figure A, B: Mean of platelets (95% CI) at baseline and 6 months (unadjusted and adjusted models) across randomized groups. Figure C, D: Between and within group differences in platelets at 6 months (unadjusted and adjusted models) among all randomized participants.

**Supplemental Figure 44: Unadjusted and adjusted analyses of MPV**

**A**

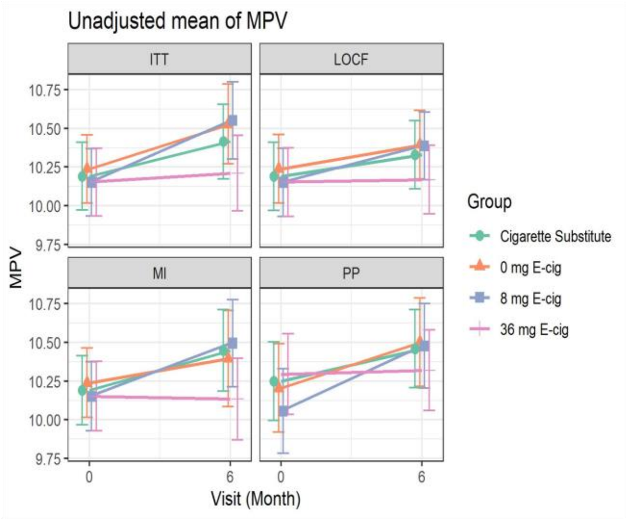

**B**

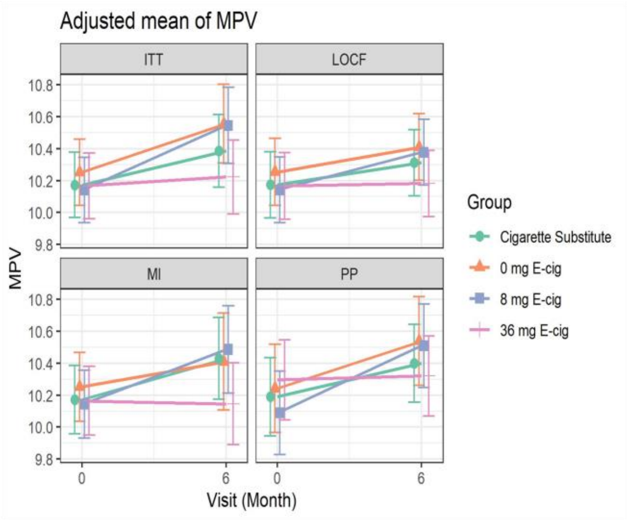

**C.** Unadjusted Mean

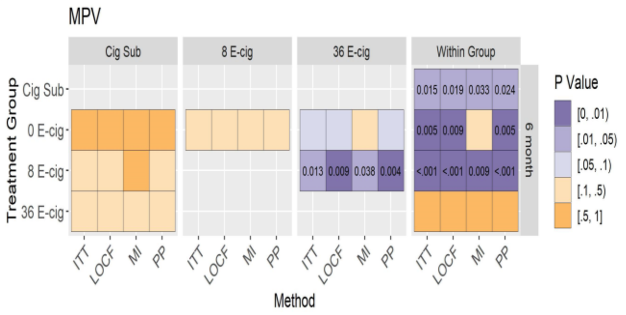

**D.** Adjusted Mean

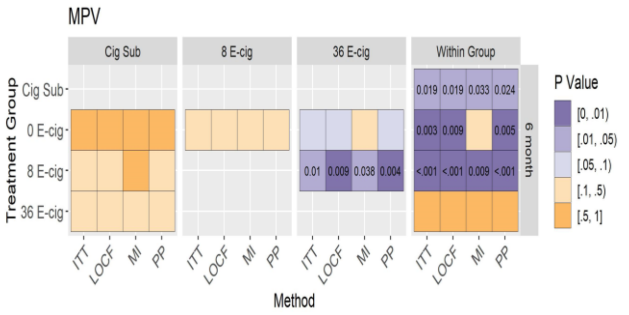

Figure A, B: Mean of MPV (95% CI) at baseline and 6 months (unadjusted and adjusted models) across randomized groups. Figure C, D: Between and within group differences in MPV at 6 months (unadjusted and adjusted models) among all randomized participants.

**Supplemental Figure 45: Unadjusted and adjusted analyses of cholesterol**

**A**

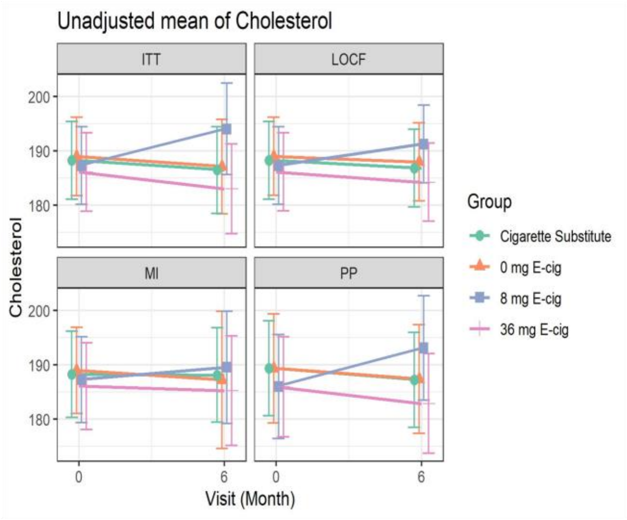

**B**

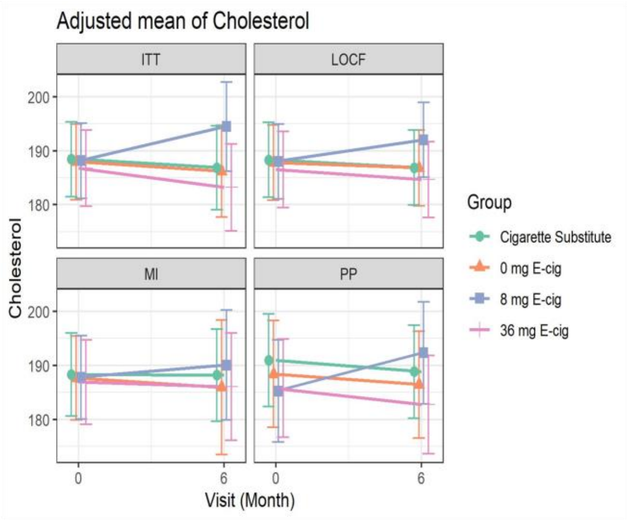

**C.** Unadjusted Mean

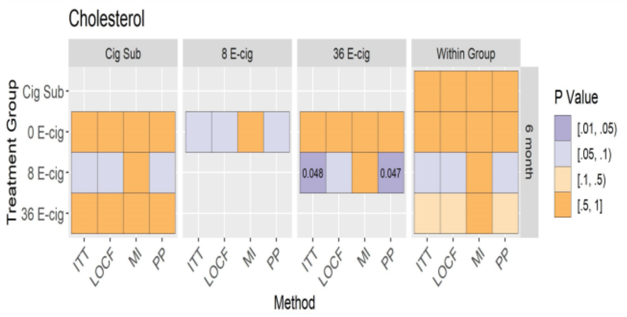

**D.** Adjusted Mean

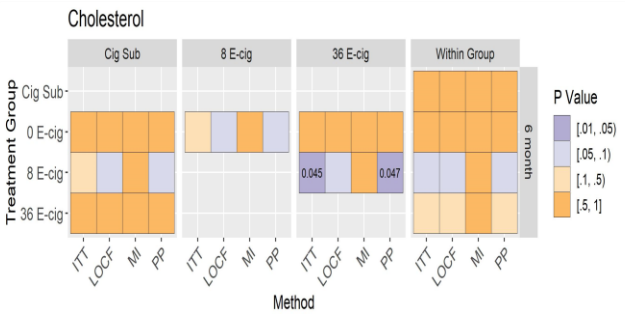

Figure A, B: Mean of cholesterol (95% CI) at baseline and 6 months (unadjusted and adjusted models) across randomized groups. Figure C, D: Between and within group differences in cholesterol at 6 months (unadjusted and adjusted models) among all randomized participants.

#### Supplemental Figure 46: Unadjusted and adjusted analyses of HDL

**A**

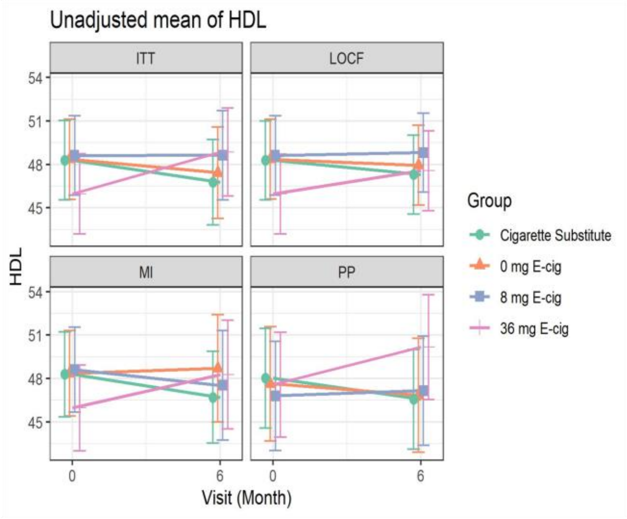

**B**

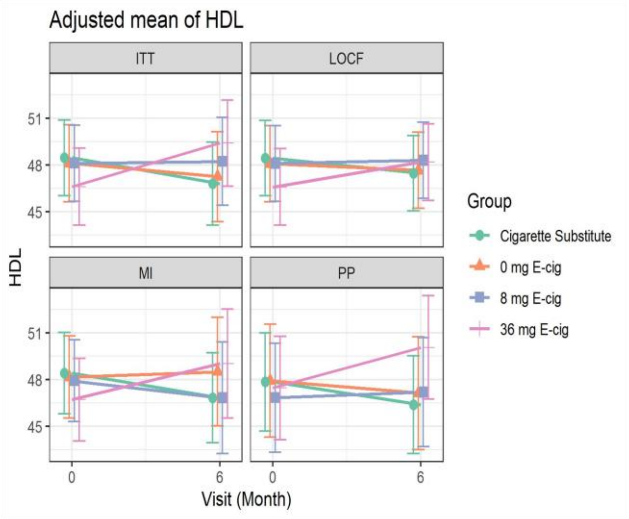

**C.** Unadjusted Mean

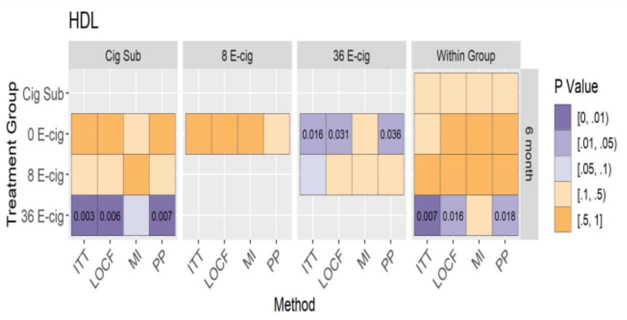

**D.** Adjusted Mean

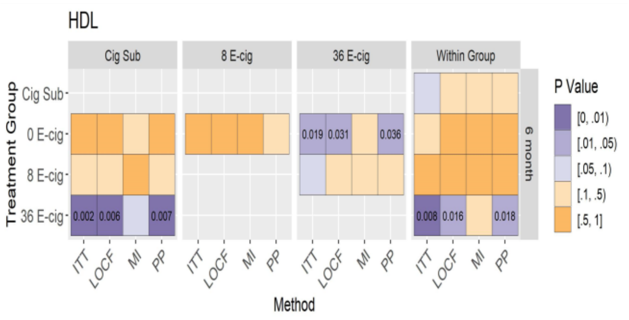

Figure A, B: Mean of HDL (95% CI) at baseline and 6 months (unadjusted and adjusted models) across randomized groups. Figure C, D: Between and within group differences in HDL at 6 months (unadjusted and adjusted models) among all randomized participants.

### Supplemental Figure 47: Unadjusted and adjusted analyses of Non-HDL

**A**

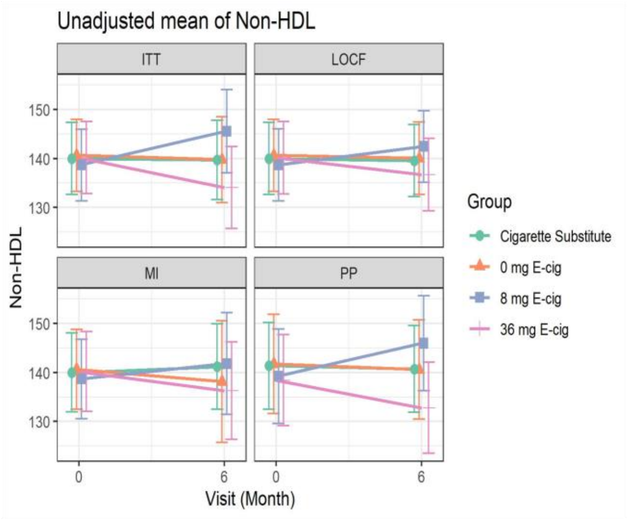

**B**

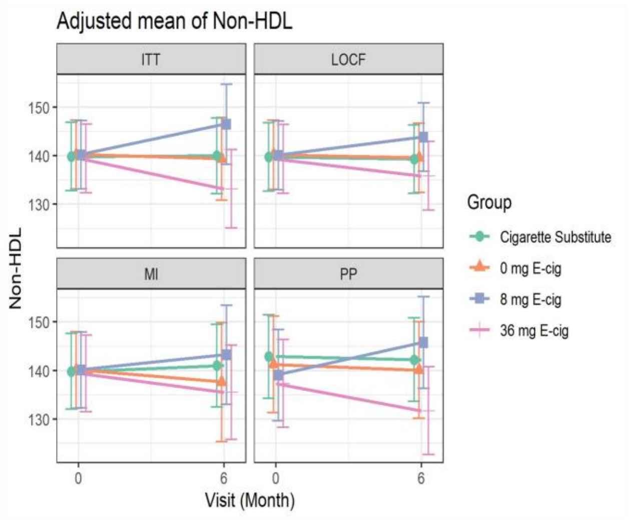

**C.** Unadjusted Mean

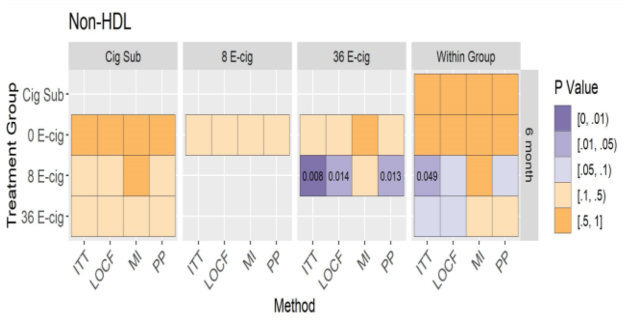

**D.** Adjusted Mean

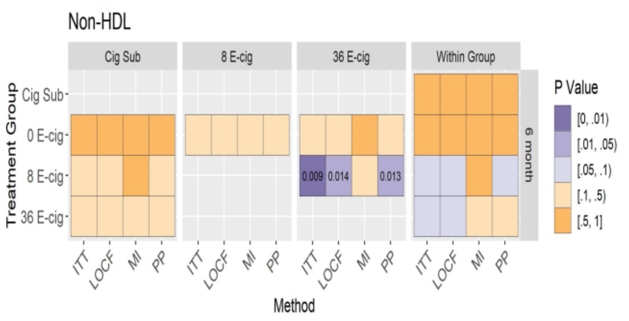

Figure A, B: Mean of non-HDL (95% CI) at baseline and 6 months (unadjusted and adjusted models) across randomized groups. Figure C, D: Between and within group differences in non-HDL at 6 months (unadjusted and adjusted models) among all randomized participants.

#### Supplemental Figure 48: Unadjusted and adjusted analyses of cholesterol/HDL ratio

**A**

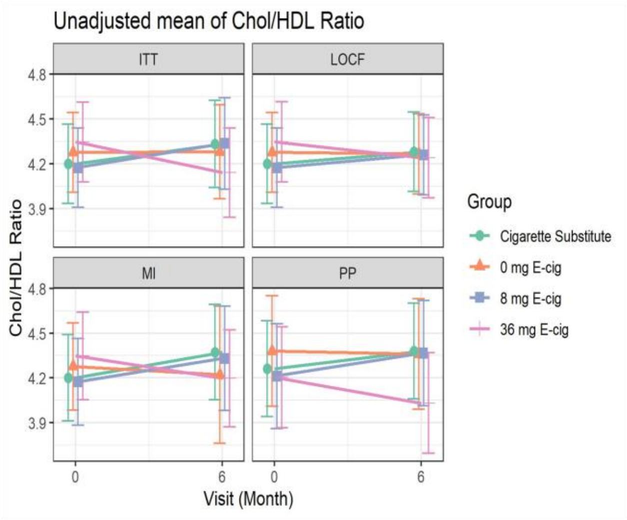

**B**

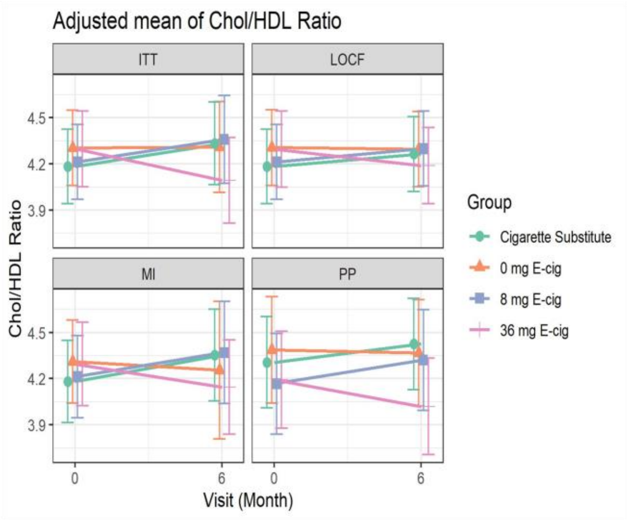

**C.** Unadjusted Mean

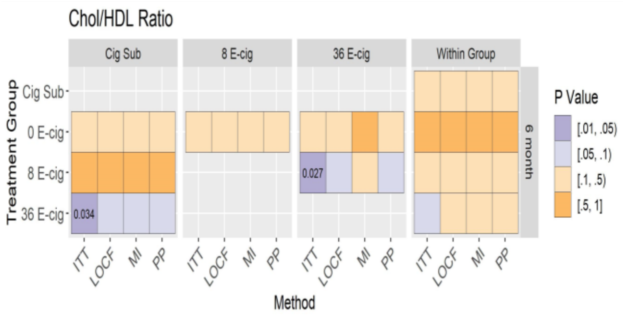

**D.** Adjusted Mean

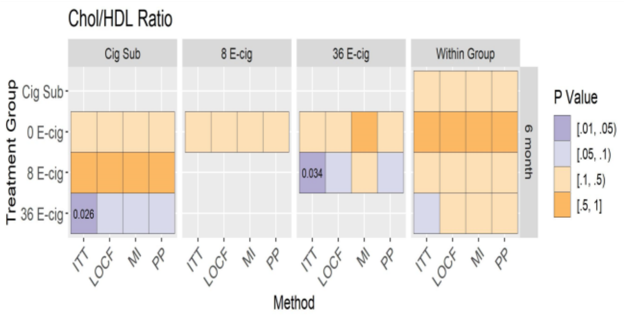

Figure A, B: Mean of cholesterol/HDL Ratio (95% CI) at baseline and 6 months (unadjusted and adjusted models) across randomized groups. Figure C, D: Between and within group differences in cholesterol/HDL Ratio at 6 months (unadjusted and adjusted models) among all randomized participants.

#### Supplemental Figure 49: Unadjusted and adjusted analyses of LDL

**A**

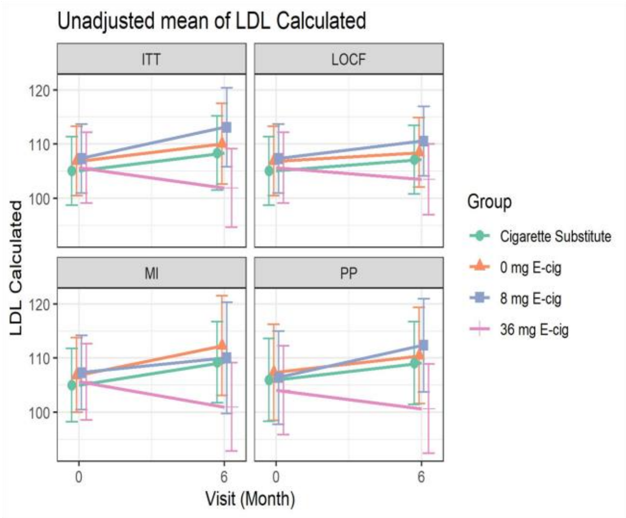

**B**

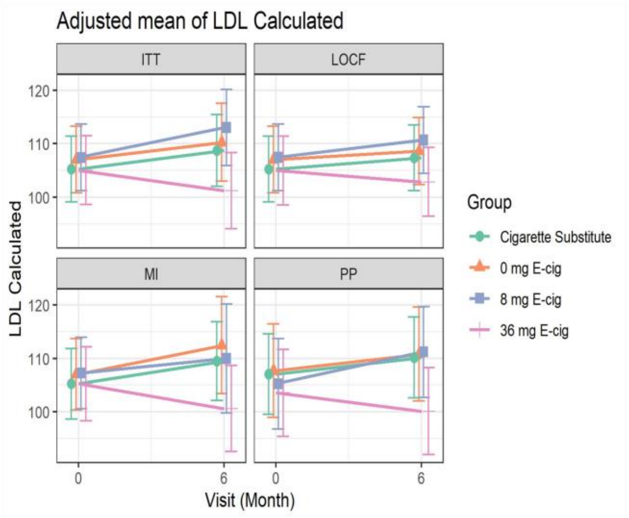

**C.** Unadjusted Mean

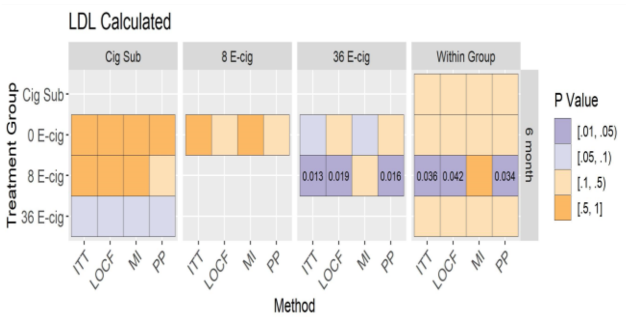

**D.** Adjusted Mean

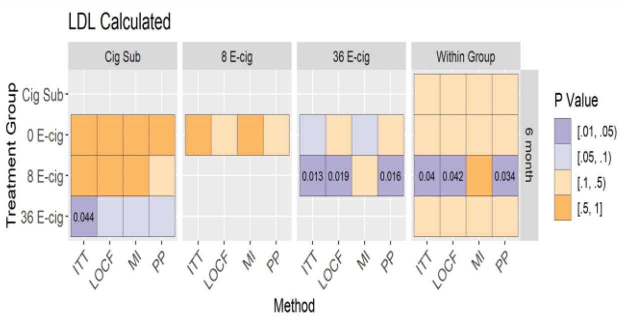

Figure A, B: Mean of LDL (95% CI) at baseline and 6 months (unadjusted and adjusted models) across randomized groups. Figure C, D: Between and within group differences in LDL at 6 months (unadjusted and adjusted models) among all randomized participants.

#### Supplemental Figure 50: Unadjusted and adjusted analyses of triglycerides

**A**

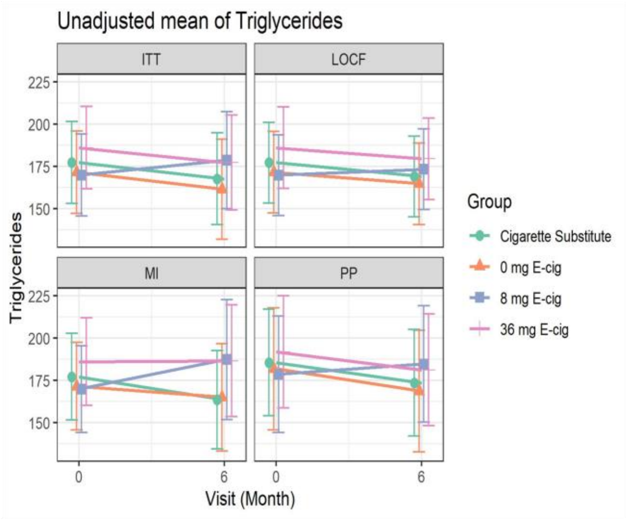

**B**

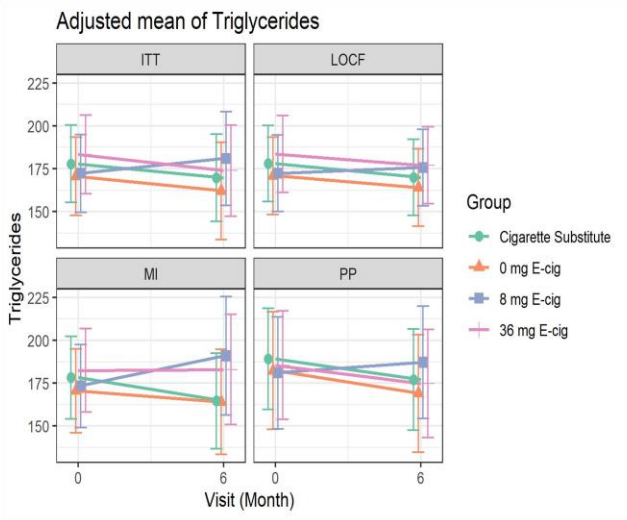

**C.** Unadjusted Mean

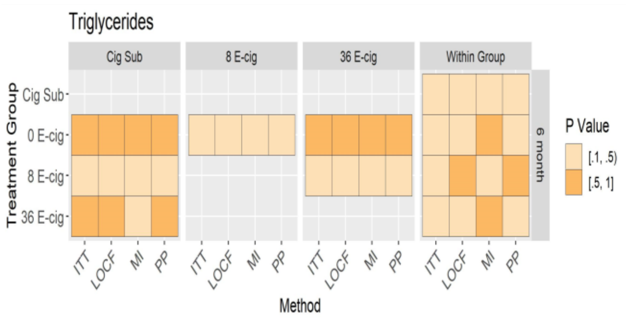

**D.** Adjusted Mean

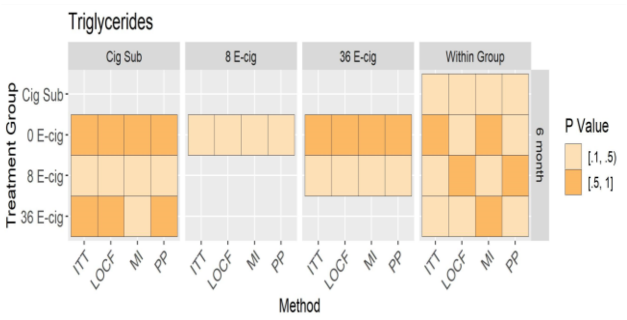

Figure A, B: Mean of triglycerides (95% CI) at baseline and 6 months (unadjusted and adjusted models) across randomized groups. Figure C, D: Between and within group differences in triglycerides at 6 months (unadjusted and adjusted models) among all randomized participants.

### Supplemental Figure 51: Unadjusted and adjusted analyses of weight

**A**

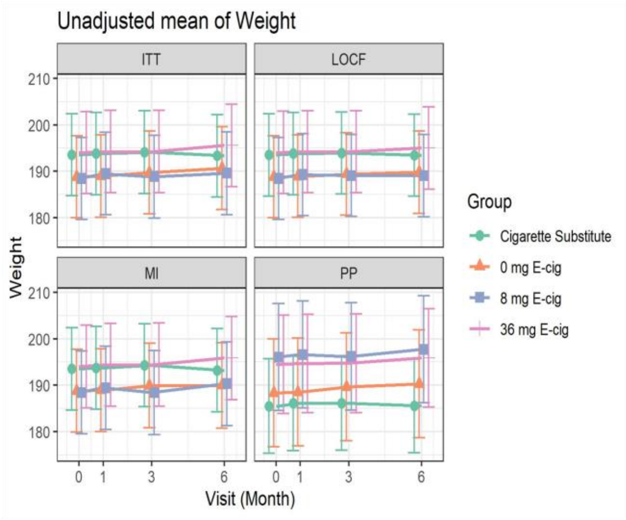

**B**

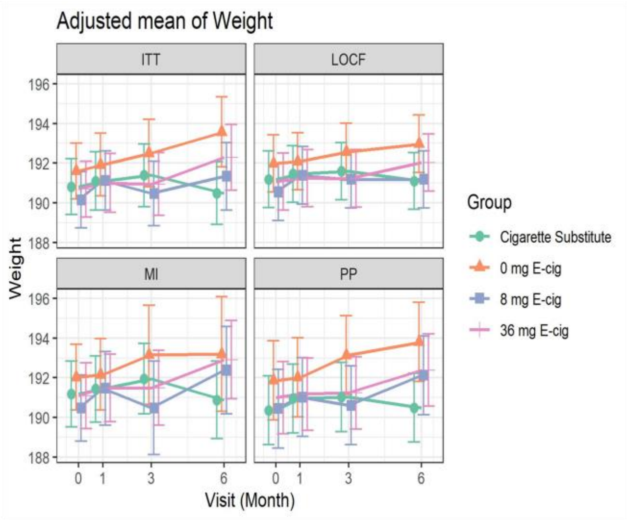

**C.** Unadjusted Mean

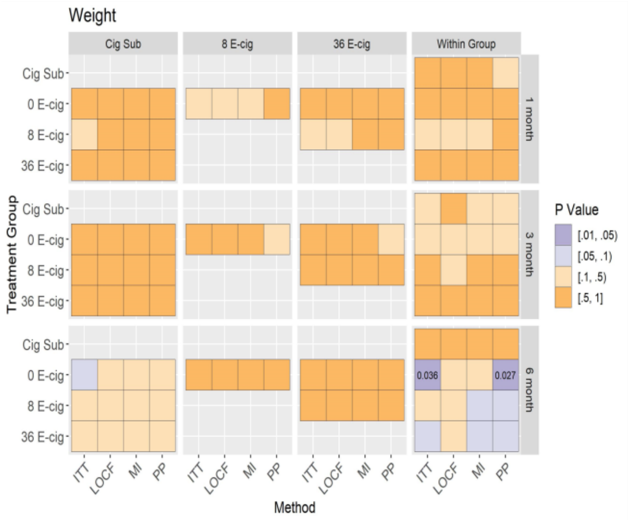

**D.** Adjusted Mean

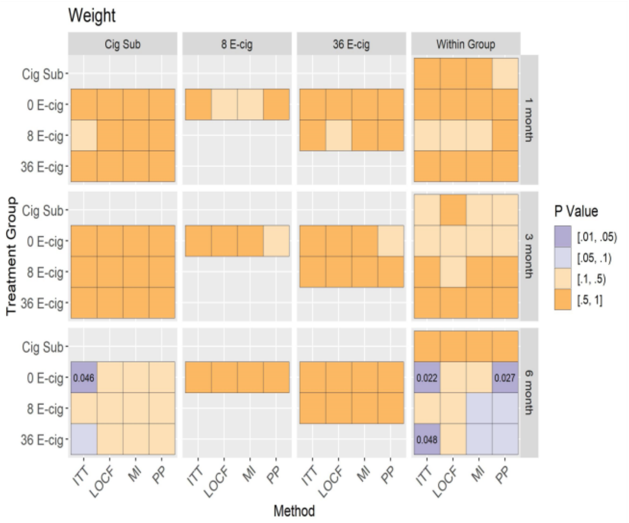

Figure A, B: Mean of weight (95% CI) at baseline, 1, 3, and 6 months (unadjusted and adjusted models) across randomized groups. Figure C, D: Between group differences in weight at baseline, 1, 3 and 6 months (unadjusted and adjusted models), and within group differences between baseline and follow-up visits among all randomized participants.

#### Supplemental Figure 52: Unadjusted and adjusted analyses of BMI

**A**

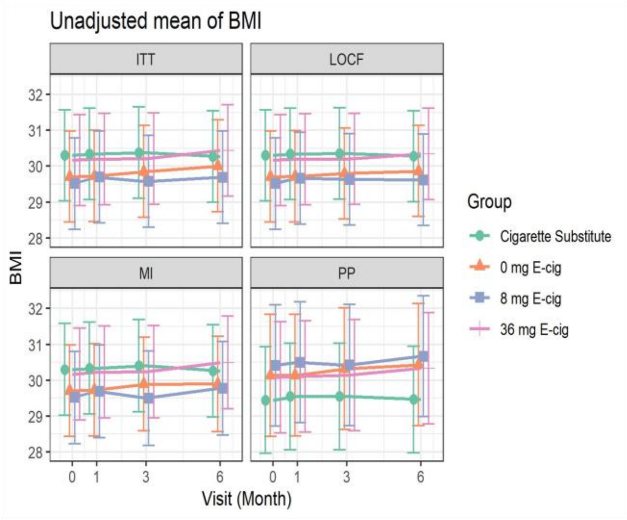

**B**

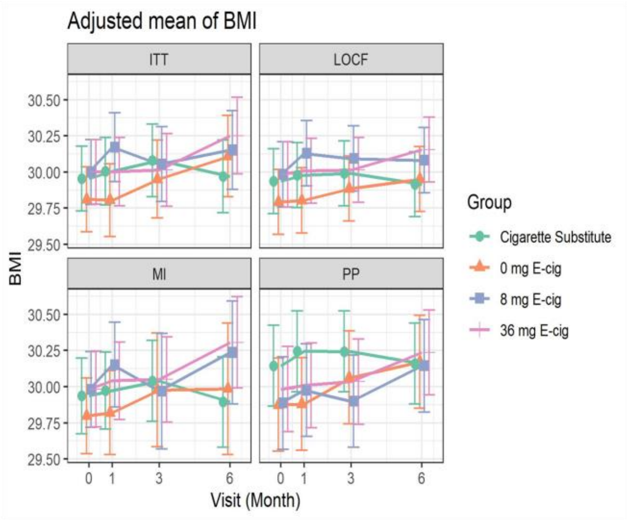

**C.** Unadjusted Mean

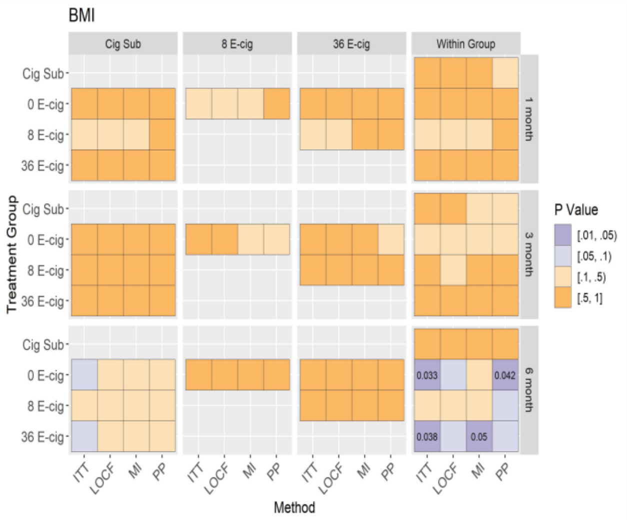

**D.** Adjusted Mean

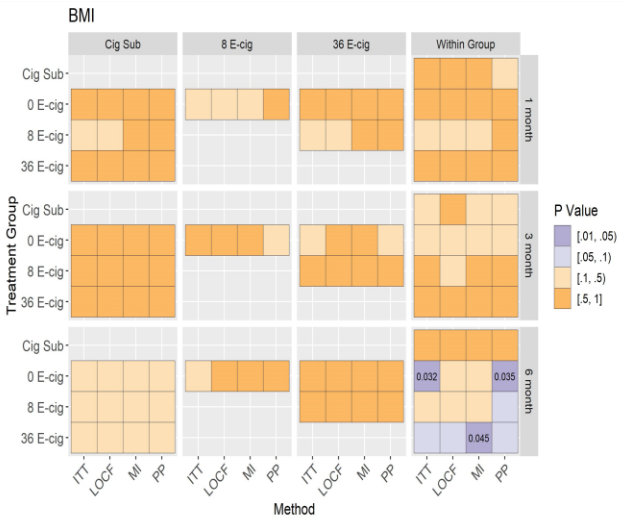

Figure A, B: Mean of BMI (95% CI) at baseline, 1, 3, and 6 months (unadjusted and adjusted models) across randomized groups. Figure C, D: Between group differences in BMI at baseline, 1, 3 and 6 months (unadjusted and adjusted models), and within group differences between baseline and follow-up visits among all randomized participants.

#### Supplemental Figure 53: Unadjusted and adjusted analyses of heart rate

**A**

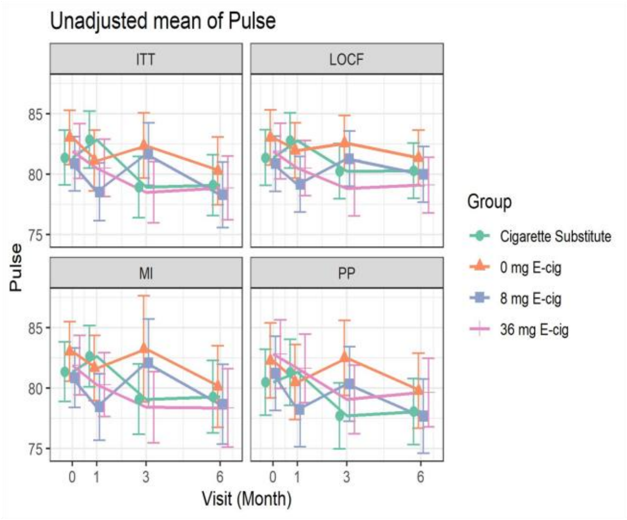

**B**

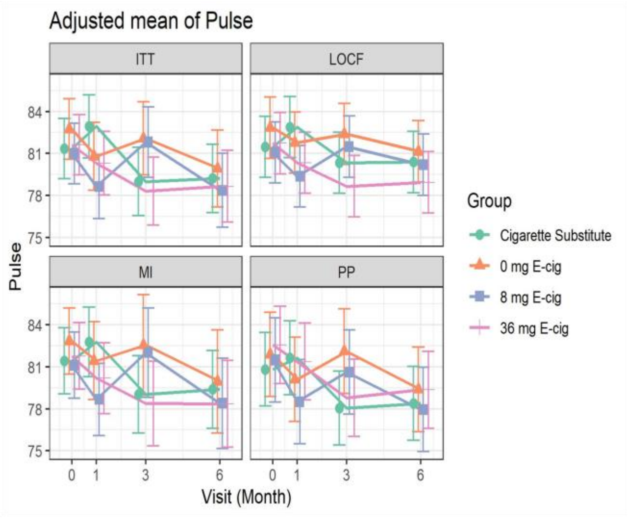

**C.** Unadjusted Mean

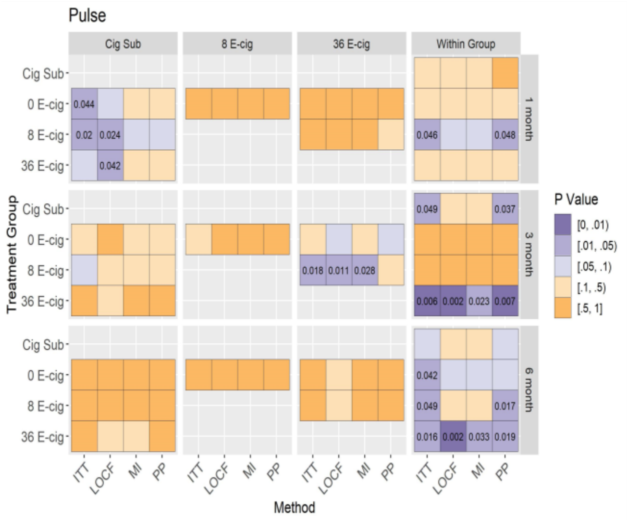

**D.** Adjusted Mean

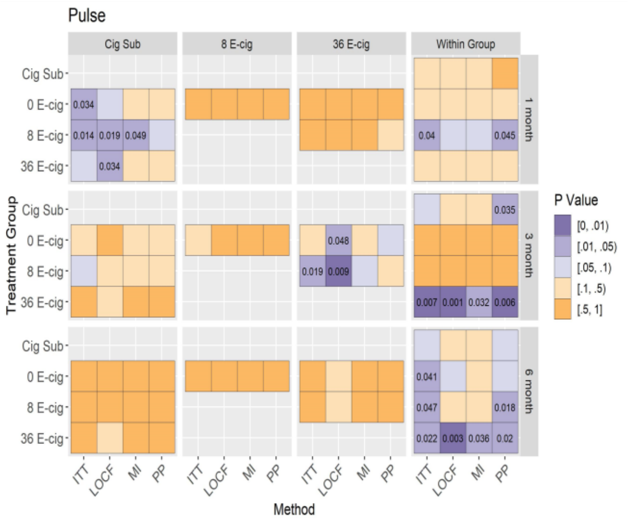

Figure A, B: Mean of heart rate (95% CI) at baseline, 1, 3, and 6 months (unadjusted and adjusted models) across randomized groups. Figure C, D: Between group differences in heart rate at baseline, 1, 3 and 6 months (unadjusted and adjusted models), and within group differences between baseline and follow-up visits among all randomized participants.

**Supplemental Figure 54: Unadjusted and adjusted analyses of systolic BP**

**A**

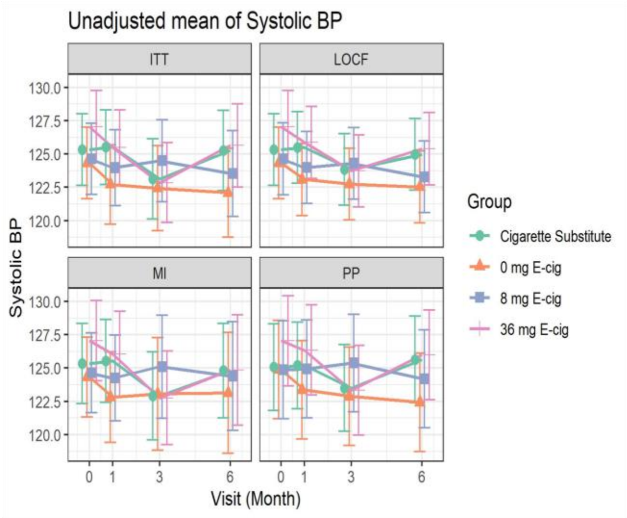

**B**

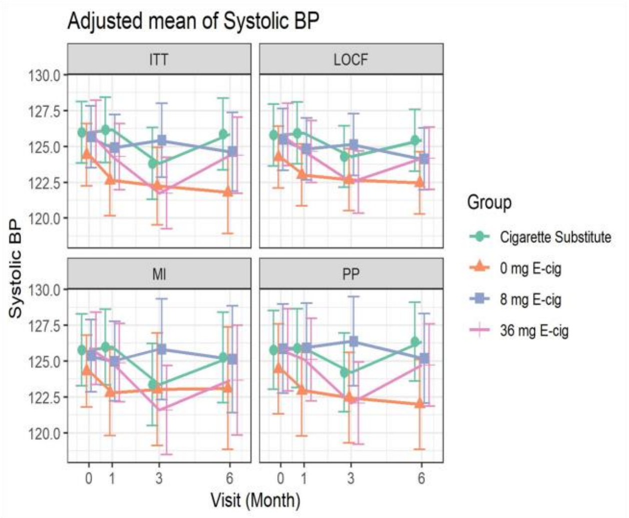

**C.** Unadjusted Mean

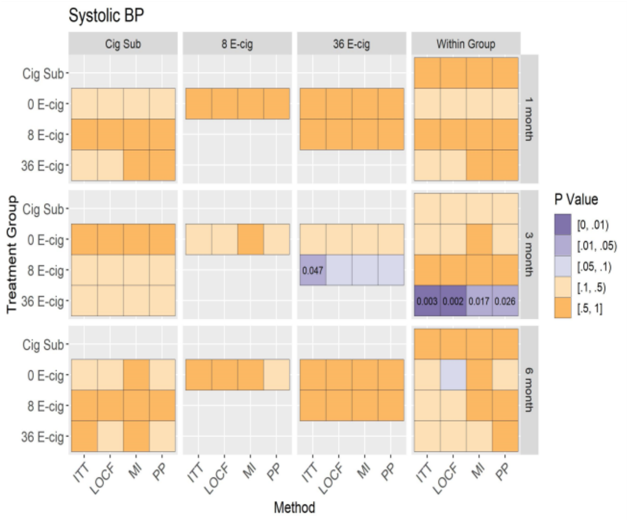

**D.** Adjusted Mean

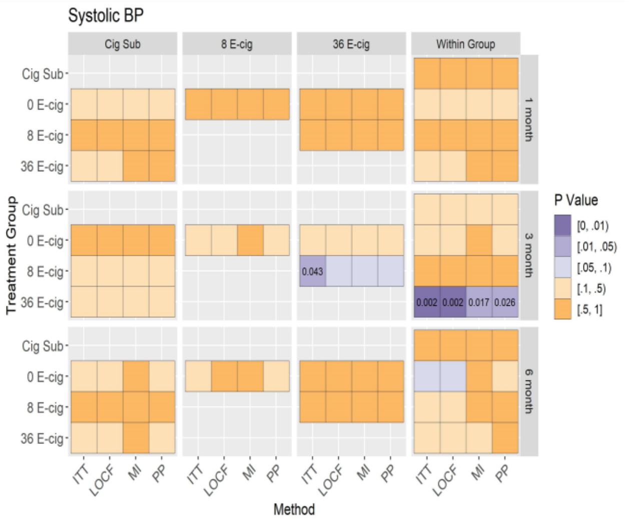

Figure A, B: Mean of systolic BP (95% CI) at baseline, 1, 3, and 6 months (unadjusted and adjusted models) across randomized groups. Figure C, D: Between group differences in systolic BP at baseline, 1, 3 and 6 months (unadjusted and adjusted models), and within group differences between baseline and follow-up visits among all randomized participants.

#### Supplemental Figure 55: Unadjusted and adjusted analyses of diastolic BP

**A**

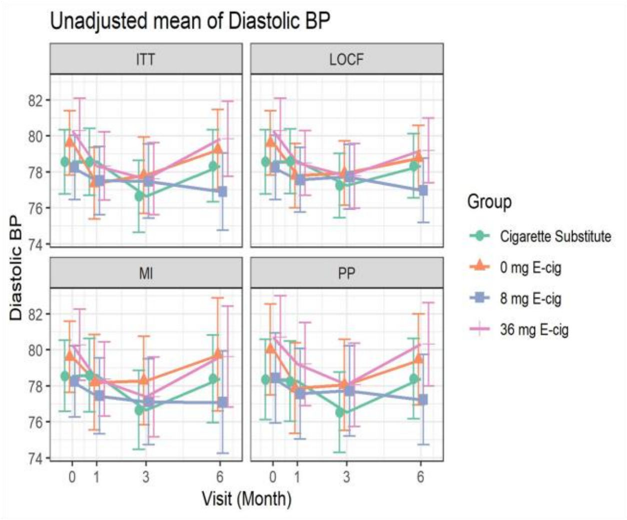

**B**

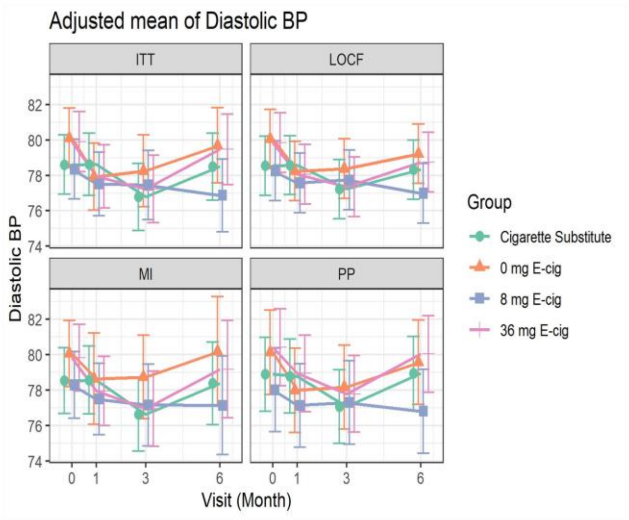

**C.** Unadjusted Mean

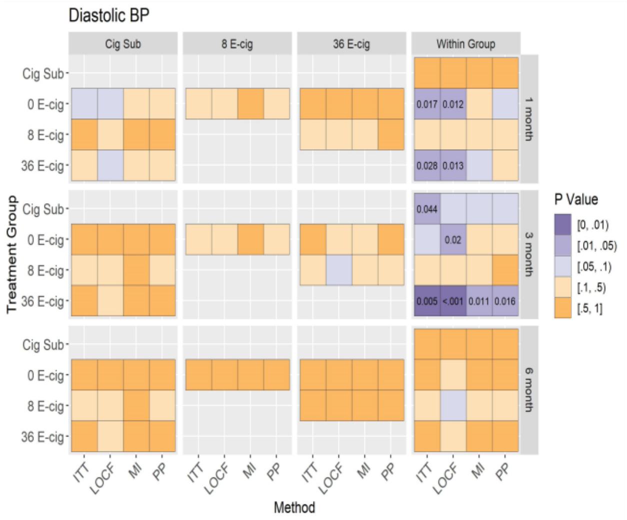

**D.** Adjusted Mean

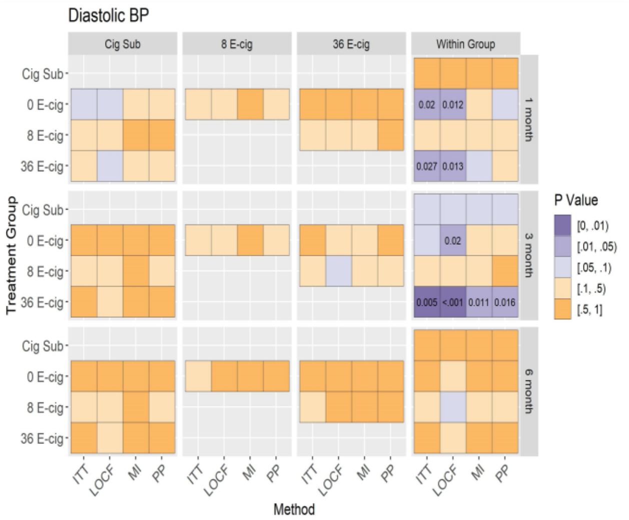

Figure A, B: Mean of diastolic BP (95% CI) at baseline, 1, 3, and 6 months (unadjusted and adjusted models) across randomized groups. Figure C, D: Between group differences in diastolic BP at baseline, 1, 3 and 6 months (unadjusted and adjusted models), and within group differences between baseline and follow-up visits among all randomized participants.

#### Supplemental Figure 56: Unadjusted and adjusted analyses of waist-to-hip ratio

**A**

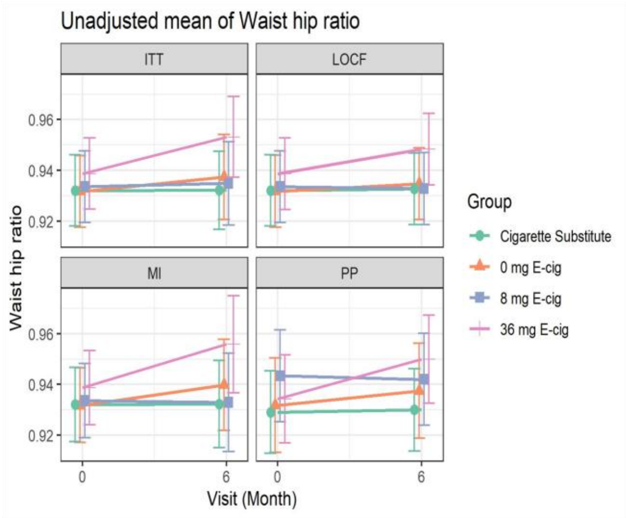

**B**

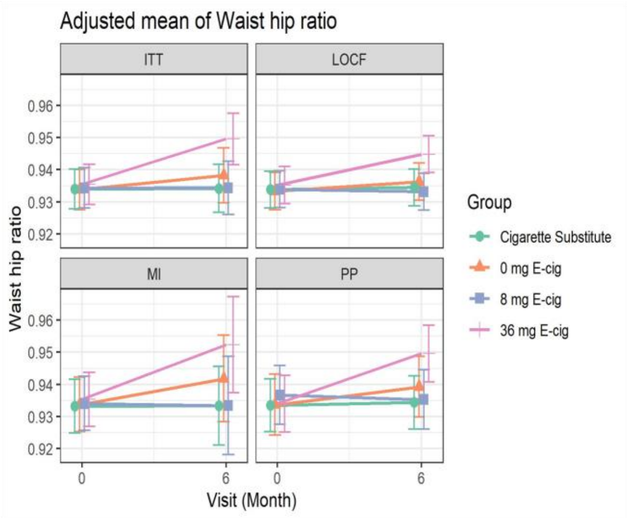

**C.** Unadjusted Mean

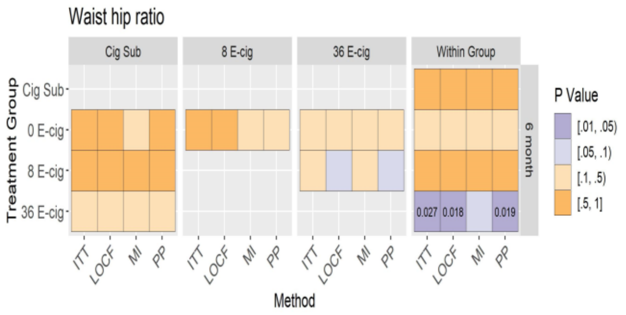

**D.** Adjusted Mean

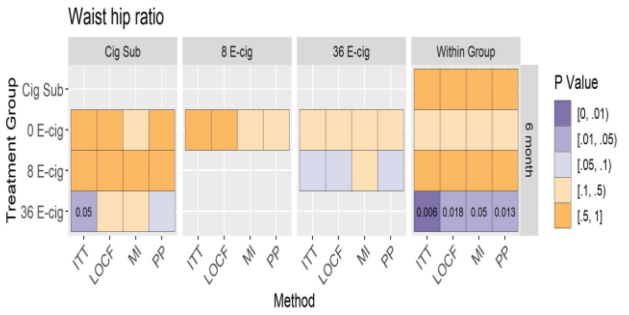

Figure A, B: Mean of waist-to-hip ratio (95% CI) at baseline and 6 months (unadjusted and adjusted models) across randomized groups. Figure C, D: Between and within group differences in waist-to-hip ratio at 6 months (unadjusted and adjusted models) among all randomized participants.

### Supplemental Figure 57: Unadjusted and adjusted analyses of INTERHEART risk score

**A**

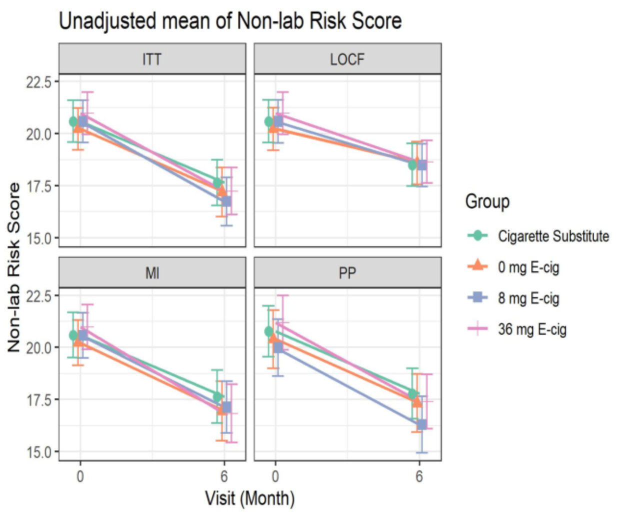

**B**

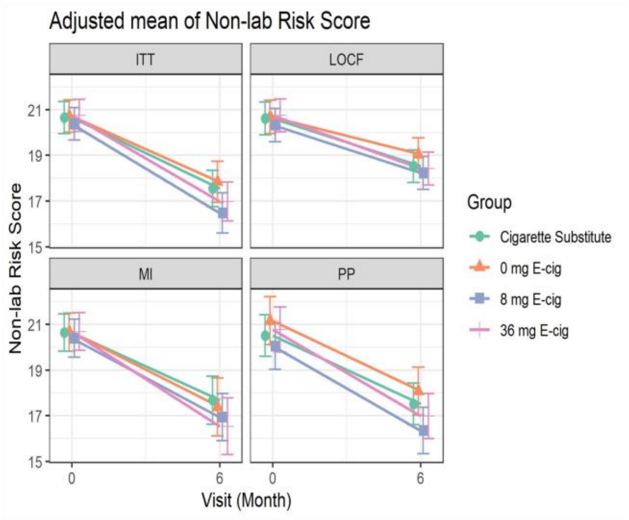

**C.** Unadjusted Mean

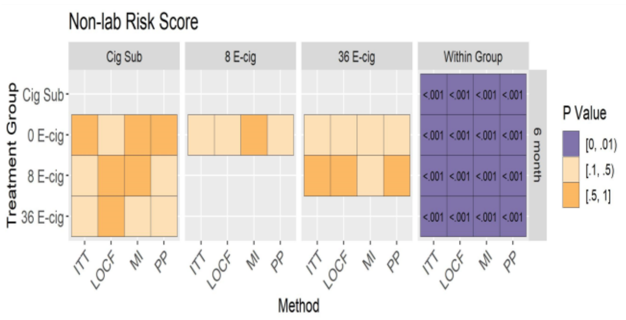

**D.** Adjusted Mean

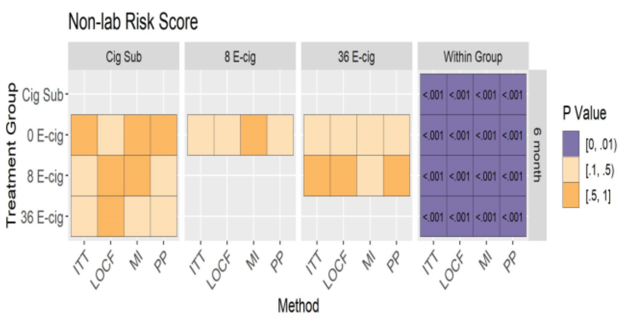

Figure A, B: Mean of INTERHEART risk score (95% CI) at baseline and 6 months (unadjusted and adjusted models) across randomized groups. Figure C, D: Between and within group differences in INTERHEART risk score at 6 months (unadjusted and adjusted models) among all randomized participants.
